# A systematic review of associations between risk factors during the first 1000 days of life and cardiometabolic outcomes later in childhood

**DOI:** 10.1101/2024.06.14.24308770

**Authors:** Marco Brandimonte-Hernández, Francisco Javier Ruiz-Ojeda, Sophia M Blaauwendraad, Arwen SJ Kamphuis, Eduard Flores-Ventura, Marieke Abrahamse-Berkeveld, Maria Carmen Collado, Janna A van Diepen, Patricia Iozzo, Karen Knipping, Carolien A van Loo-Bouwman, Romy Gaillard, Ángel Gil

## Abstract

**Importance:** Childhood obesity increases cardiometabolic risk during childhood among individuals aged 2-18 years. These cardiometabolic outcomes include glucose intolerance, dyslipidemia, hypertension, metabolic syndrome, and type 2 diabetes (T2D). In the current state of research, a comprehensive review identifying all early-life exposures and risk factors that could predict cardiometabolic risk in children is lacking.

**Objective:** To identify and evaluate the predictive early-life risk factors during the first 1,000 days of life, including preconception, pregnancy and birth, and early infancy periods for cardiometabolic risk outcomes in childhood.

**Evidence review:** The present systematic review of existing literature was conducted to revise and search selected electronic databases (Medline, EMBASE, WEB OF SCIENCE, SCOPUS, and Cochrane CENTRAL) for longitudinal studies published between the database’s inception and August ^17^, 2022. This systematic review protocol was registered to PROSPERO, CRD42022355152, and following the PRISMA guidelines. We selected articles that studied the risk factors in mothers, fathers and infants, from preconception to infancy, for childhood cardiometabolic outcomes between 2 and 18 years.

**Findings:** In 68 studies, we identified 229 associations between exposures and childhood cardiometabolic outcomes namely glucose intolerance, dyslipidemia, hypertension, metabolic syndrome, and T2D. The majority of associations (n=162) were positively associated with cardiometabolic risk factors. Pregnancy and birth risk factors were the categories with the most reported associations (86%). Among them, the most frequently assessed characteristics were birth anthropometrics (n=75), sociodemographics data (n=47), and pregnancy complications factors (n=34). However, we only identified few risk factors during preconception. In infancy period, breastfeeding (n=16) and infant anthropometrics (n=15) were consistently associated with cardiometabolic outcomes. In all periods, the most studied associations were identified for hypertension and metabolic syndrome.

**Conclusions and relevance:** Birth anthropometrics, sociodemographics, and pregnancy complication factors were the most frequently reported predictive factors associated with a higher risk for cardiometabolic outcomes in children, particularly hypertension and metabolic syndrome. These results of this study are useful for predicting the risk for childhood cardiometabolic outcomes and for the modifiable factors. They also may facilitate the design of approaches aimed at the alteration of several behaviours from birth to infancy, encompassing both the maternal and paternal influences, as well as the preconception to infancy transition period. Future studies evaluating early-life risk factors with scarce scientific evidence, such as paternal and preconception factors, are urgently needed.

**Key points:** *Question:* What is the existing evidence of early-life risk factors during the first 1000 days of life that are associated with cardiometabolic dysfunction between 2 and 18 years?

*Findings:* Birth anthropometrics, pregnancy complications, sociodemographics and lifestyle factors are the most frequently reported associated exposures with hypertension and metabolic syndrome in children.

*Meaning:* Identifying early-life risk factors and changing behavior patterns throughout preconception and infancy can contribute to prevent metabolic diseases later in childhood.

## INTRODUCTION

Obesity increases the risk and severity of several cardiometabolic conditions, such as glucose intolerance, dyslipidemia, hypertension, metabolic syndrome, and type 2 diabetes (T2D). More than 50% of children and adolescents with obesity are diagnosed with one cardiometabolic risk condition, and 25% are diagnosed with two and more cardiometabolic risk conditions ^1,2^. The global prevalence of T2D by 2030 is estimated to increase to 10.2% from 9.3% in 2019 ^3^. Furthermore, estimations from 2021 indicate that T2D or its complications is main cause of death for nearly 7 million people worldwide, leading to an economic cost of approximately US$ 966 billion in 2021, which could reach US$ 1.03 trillion by 2030 and US$ 1.05 trillion by 2045 ^4^. Early-life exposures or risk factors during the preconception period, pregnancy and the early years of life could lead to the development of cardiometabolic features during childhood ^5^. Children rarely suffer adverse cardiovascular events as those experienced by adults, but cardiovascular and metabolic disease risk factors may already be present and related to future adult cardiovascular diseases (CVD) development ^6,7^, which are responsible for about 31% of deaths worldwide ^8^. Several studies emphasize intrauterine conditions are an important factors of cardiometabolic risk. In fact, birth weight is a sentinel marker of fetal health, reflecting both intrauterine growth and length of gestation. Interestingly, both low birth weight (LBW) (2500 g) and high birth weight (HBW) (>4000 g) are linked to cardiometabolic risk factors later in childhood ^7^. Extrauterine growth retardation seems to be also associated with a greater risk of cardiometabolic risk later in life ^9,10^. Several studies have shown that maternal factors might influence cardiometabolic health in children^11^. Thus, maternal body mass index (BMI) before pregnancy, maternal weight gain during pregnancy, gestational diabetes, maternal malnutrition, and maternal smoking during pregnancy, are independently associated with neonatal, infant, and child adiposity, and might impact the development of cardiometabolic risk factors. In addition, pregnancy complications such as preeclampsia and gestational diabetes also predict cardiovascular morbidity and mortality, and the cardiometabolic risk is often transmitted to the next generation^12^. Children who were exposed to gestational diabetes in utero may also be more susceptible to developing metabolic syndrome, since they have higher systolic blood pressure, BMI, and blood glucose than those who were not exposed to gestational diabetes ^13^. Although there is less information about how paternal factors might imprint cardiometabolic conditions during childhood, evidence indicates a relevant role for males in affecting cardiometabolic outcomes in their offspring ^14^. It has been proven that paternal BMI during preconception impairs the cardiometabolic profile of children ^15–17^.

To halt the global rise in cardiometabolic disease, prevention of risk factor development in early life is of key importance. A suboptimal fetal and postnatal development needs to be addressed as early as possible according to evidence-based recommendations to prevent the perpetuation of adiposity, which can lead to obesity in adolescence, adulthood, and subsequent generations ^18,19^. Moreover, a better understanding of early life exposures and risk factors that are associated with an increased cardiometabolic disease risk is crucial.

Hence, we conducted this systematic review to determine whether/which early-life exposures result in increased rates of cardiometabolic comorbidities in children between 2 and 18 years. By analyzing the first 1,000 days of life, we aimed to assess the quality of potential crucial early-life risk factors during preconception, pregnancy and birth, and early infancy periods for the prediction of children at risk for cardiometabolic outcomes such as glucose intolerance, dyslipidemia, hypertension, metabolic syndrome, and T2D.

## METHODS

### Systematic review protocol development

This study was embedded in a large collaborative project to conduct a systematic review on risk factors in the first 1,000 days of life for the development of cardiometabolic disorders in childhood. The purpose of our systematic review protocol was to include and evaluate all individual research studies on risk factors and noninvasive biomarkers in children and adolescents for cardiometabolic disorders during preconception, pregnancy and infancy. To investigate early childhood risk factors for cardiometabolic outcomes between 2 and 18 years, we aimed to identify case‒control, prospective observational, retrospective observational, and randomized control trials (RCTs) focusing on those risk factors and specific outcomes. Studies that included diseased populations only, such as mothers with type 1 diabetes, were excluded.

Early exposure variables related to sociodemographic factors, lifestyle factors, physical factors, environmental factors, pregnancy-related factors, and noninvasive biomarkers are referred to as risk factors. Here, risk factors correspond to exposure variables of interest that are associated with cardiometabolic outcomes such as glucose intolerance, dyslipidemia, hypertension, metabolic syndrome, and T2D. We assessed risk factors in mothers, fathers, and offspring during preconception, pregnancy, and infancy up to 2 years of age, together covering the first 1000 days of life. The outcomes of interest in this current review were self-reported, physician- or researcher-diagnosed; glucose intolerance; dyslipidemia; hypertension; metabolic syndrome; and T2D.

### Data sources, search strategy, and screening criteria

This systematic review protocol was registered to PROSPERO, CRD42022355152. Our search terms for Medline, EMBASE, WEB OF SCIENCE, SCOPUS, and Cochrane CENTRAL (**Text S1**) relate to eligible citations published in English before August 17, 2022. We included prospective and retrospective longitudinal observational studies which identified risk factors associated with offspring outcomes of our interest. A prospective comparison of treatment allocation effects on outcome was also considered. The exclusion criteria were: studies with outcomes before 2 years, studies reporting only intermediate phenotypes or continuous traits, or studies assessing only endpoints other than the cardiometabolic outcomes of interest. Cross-sectional studies and studies of diseased populations were excluded, except for genetics studies. Two independent reviewers screened at title and abstract level. For accepted citations, two independent reviewers examined the entire manuscript. A third reviewer resolved conflicts at all screening levels. The consensus of the full group was followed on conflicts not resolved by the third reviewer. The covidence online systematic review tracking platform was used for all screenings.

### Data extraction and synthesis of results

For eligible manuscripts, we developed and piloted a data extraction template. In addition to manuscript information, the data-set included details about the study design, demographics of the population, exposure and outcome assessment, and diagnostic criteria. A total of 8 broad categories of exposures were recorded: (i) parental lifestyle factors, (ii) parental physical factors, (iii) environmental factors, (iv) pregnancy-related factors, (v) birth anthropometrics, (vi) feeding patterns, (vii) infant anthropometrics, and (viii) cord blood biomarkers.

### Quality assessment (risk of bias) and synthesis

For cohort studies, case‒control studies, and randomized clinical trials (RCTs), the Joanna Briggs Institute (JBI) critical appraisal tool was used to assess the quality of the included studies ^20^. For case‒control studies, 9 items were used to assess quality, including group’s comparison, assessment of the exposure and outcome, confounding, exposure period of interest, and statistical methodology. For cohort studies, 11 items were used to assess quality, including recruitment, assessment of exposure and outcome, confounding, statistical methodology, and follow-up. In RCTs, 10 items were evaluated according to the JBI criteria, including selection and allocation, intervention, administration, outcome assessment, follow-up, and statistical analysis. Using the guidelines, each JBI item was categorized as ‘yes’, ‘no’, ‘unclear’ or ‘not applicable’. Any uncertainties in the assessment were further discussed by the entire research team. Studies with overall scores of 50% or less on questions answered with ‘Yes’, 50% until 70% questions answered with ‘Yes’, and 70% or more questions answered with ‘Yes’, were considered low, moderate and high quality, respectively.

### Quality assessment of risk factors for prediction and prevention

For quality assessment, we selected those risk factors we considered consistently associated with childhood obesity. Generally, we defined consistently associated risk factors as those for which more than 50% of studies report an association in the same direction, in at least two studies of moderate or high quality. Based on the International Life Sciences Institute (ILSI) Europe Marker Validation Initiative, we developed and piloted a criteria template for quality assessment of risk factors ^21^ (**Table S1**). Based on methodological aspects, the study objective, modifiability, predictiveness, and early-life risk factors were evaluated. Two independent reviewers scored the risk factors at four different levels: proven, strong, moderate, and low. The whole group discussed conflicts until consensus was reached.

## RESULTS

### Characteristics of participants and the screening process

Figure S1 shows the flow chart for screening all studies (n=35584), of which about one-half were excluded as duplicates (n=17974). A total of 17610 full-text or abstract articles were screened by two independent reviewers, and 16059 were scored irrelevant. Full-text screening was conducted on 1551 studies; 302 were extracted, and 112 were analyzed for associations. Ultimately, 68 articles were meeting the inclusion criteria and were included for the current systematic review.

**Figure 1.**
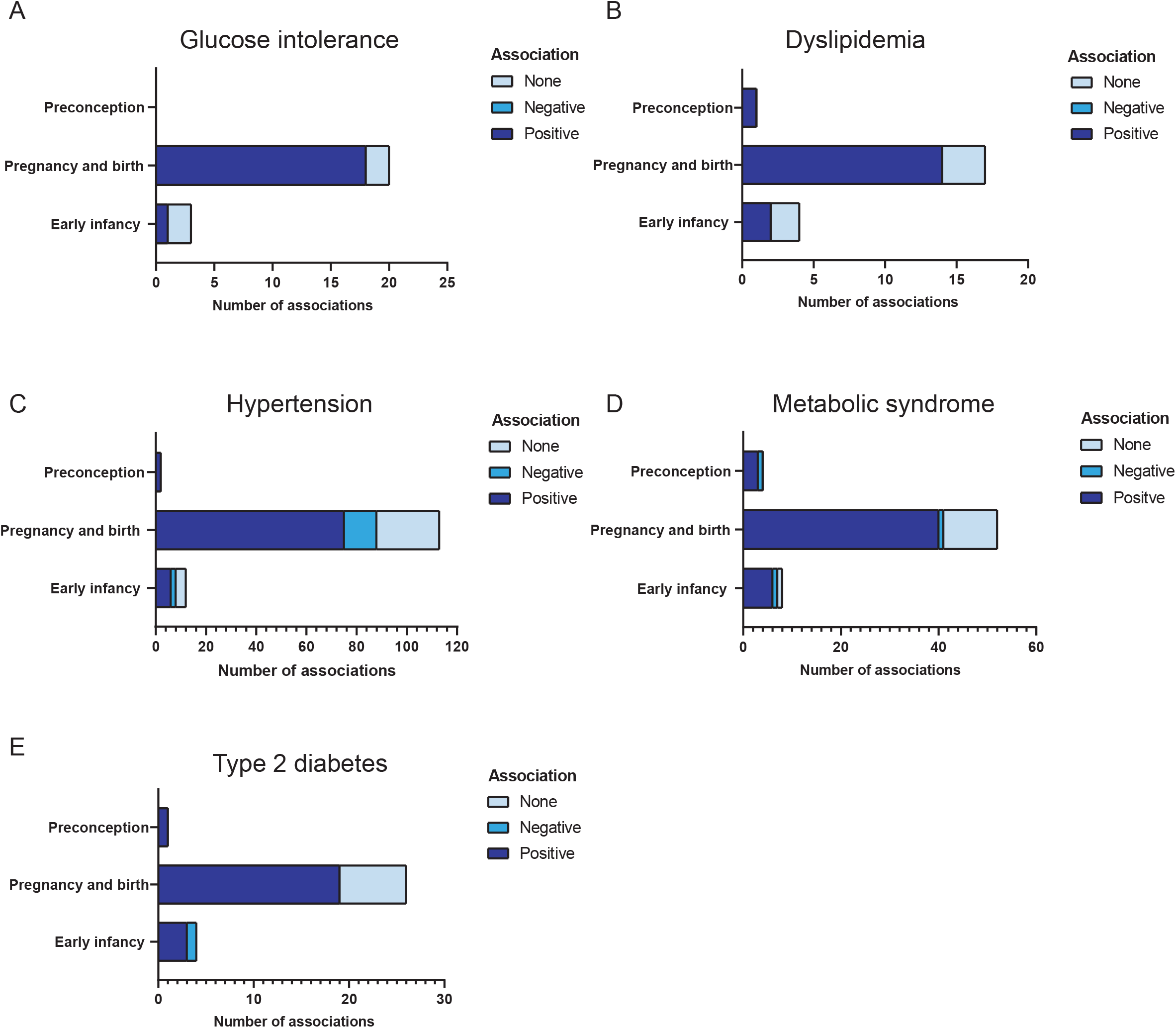
Positive, negative, and null associations between the main categories of risk factors preconception, pregnancy and birth, and early infancy and cardiometabolic outcomes: glucose intolerance (A), dyslipidemia (B), hypertension (C), metabolic syndrome (D), and type 2 diabetes (E).

The studies were published between 1991 and 2022 in 31 different countries, with the majority of the data coming from America, Asia, and Europe. Among the 68 articles included, 59 were observational studies, of which 38 were prospective and 21 were retrospective studies. Seven were case-control studies, and only 2 were RCT interventional studies. The sample sizes of the different studies ranged from 50 to 155,411 participants. **Table 1** shows the characteristics, the main results and the cardiometabolic outcome definitions for each study.

**Table 1.**
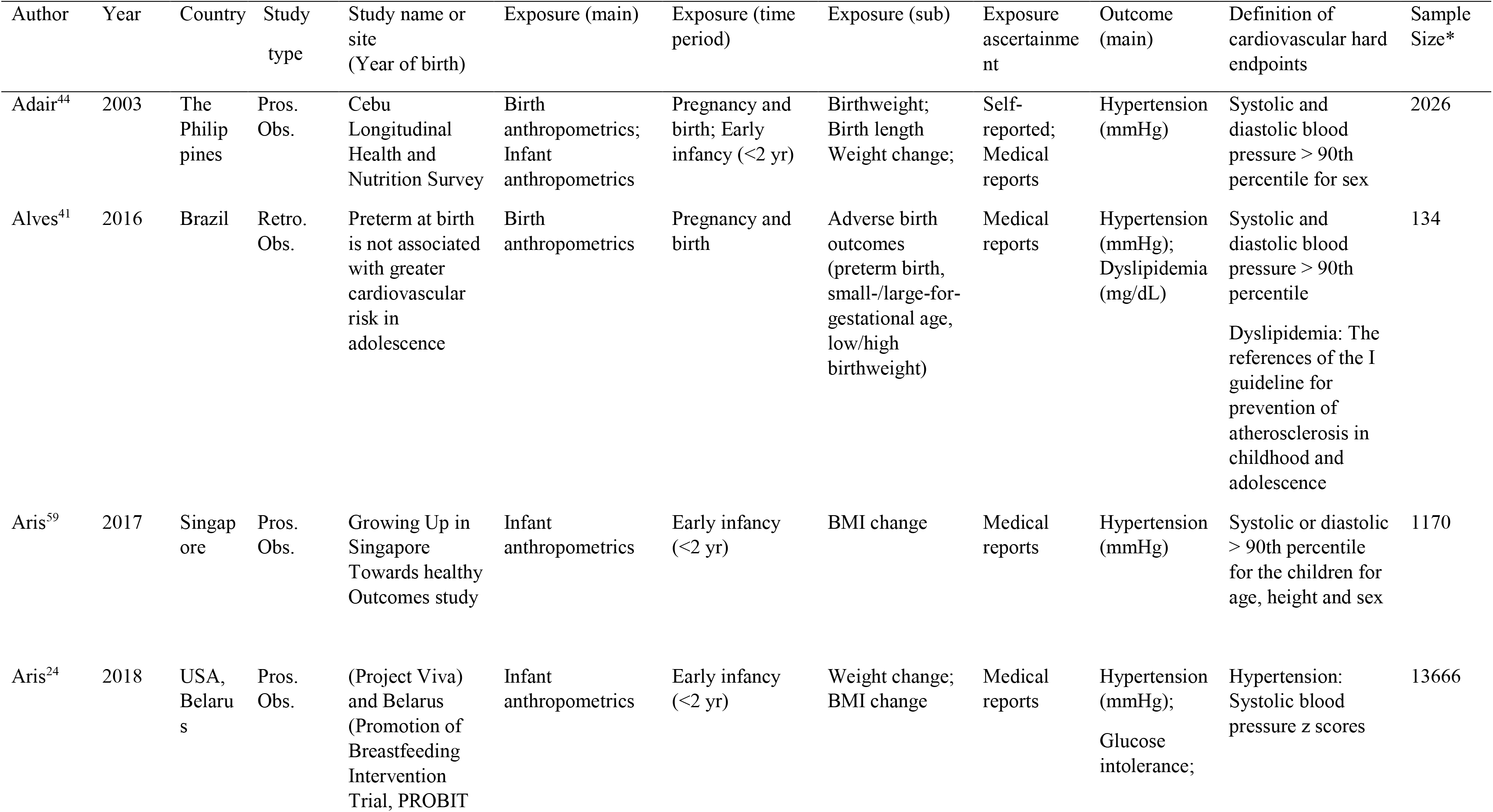
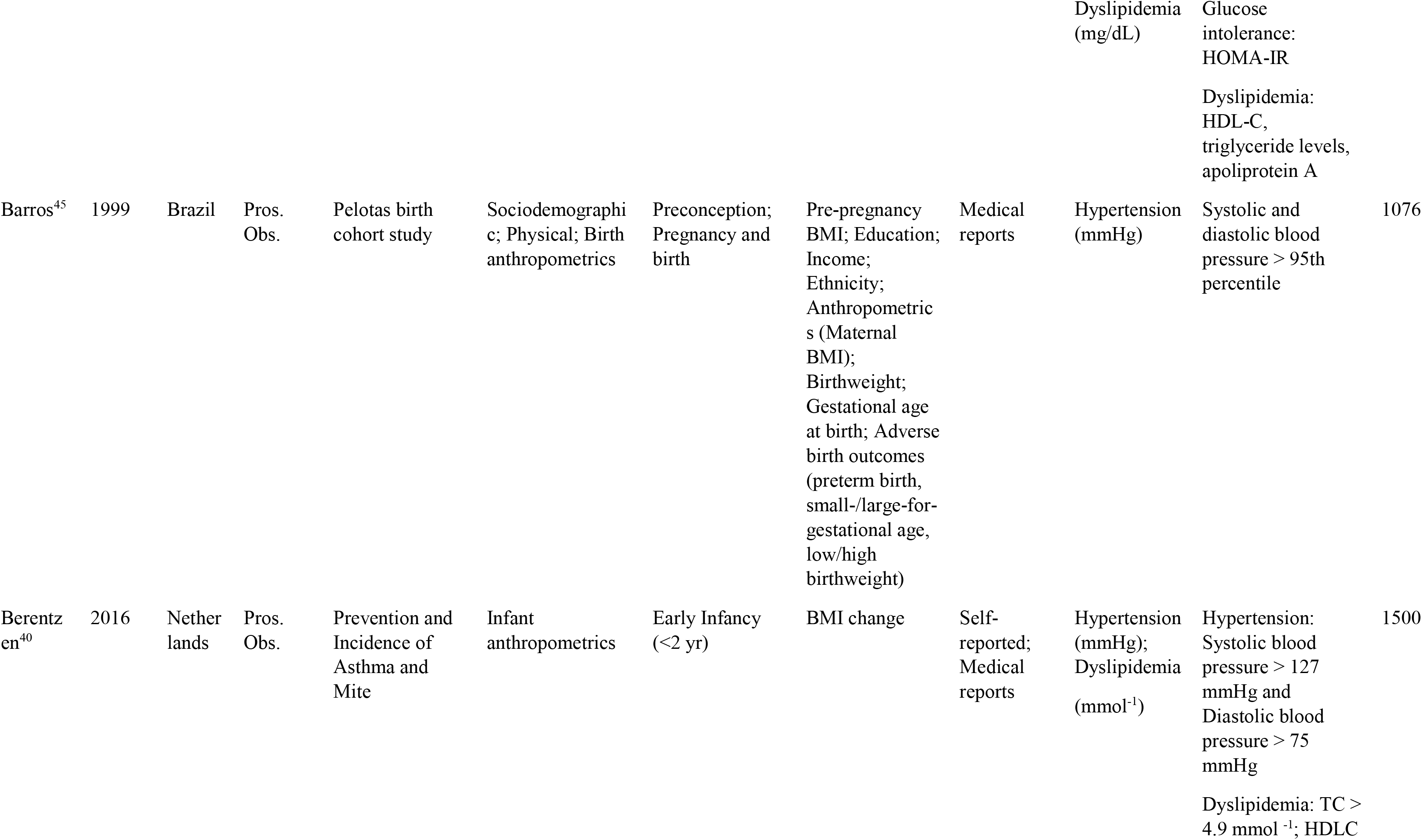
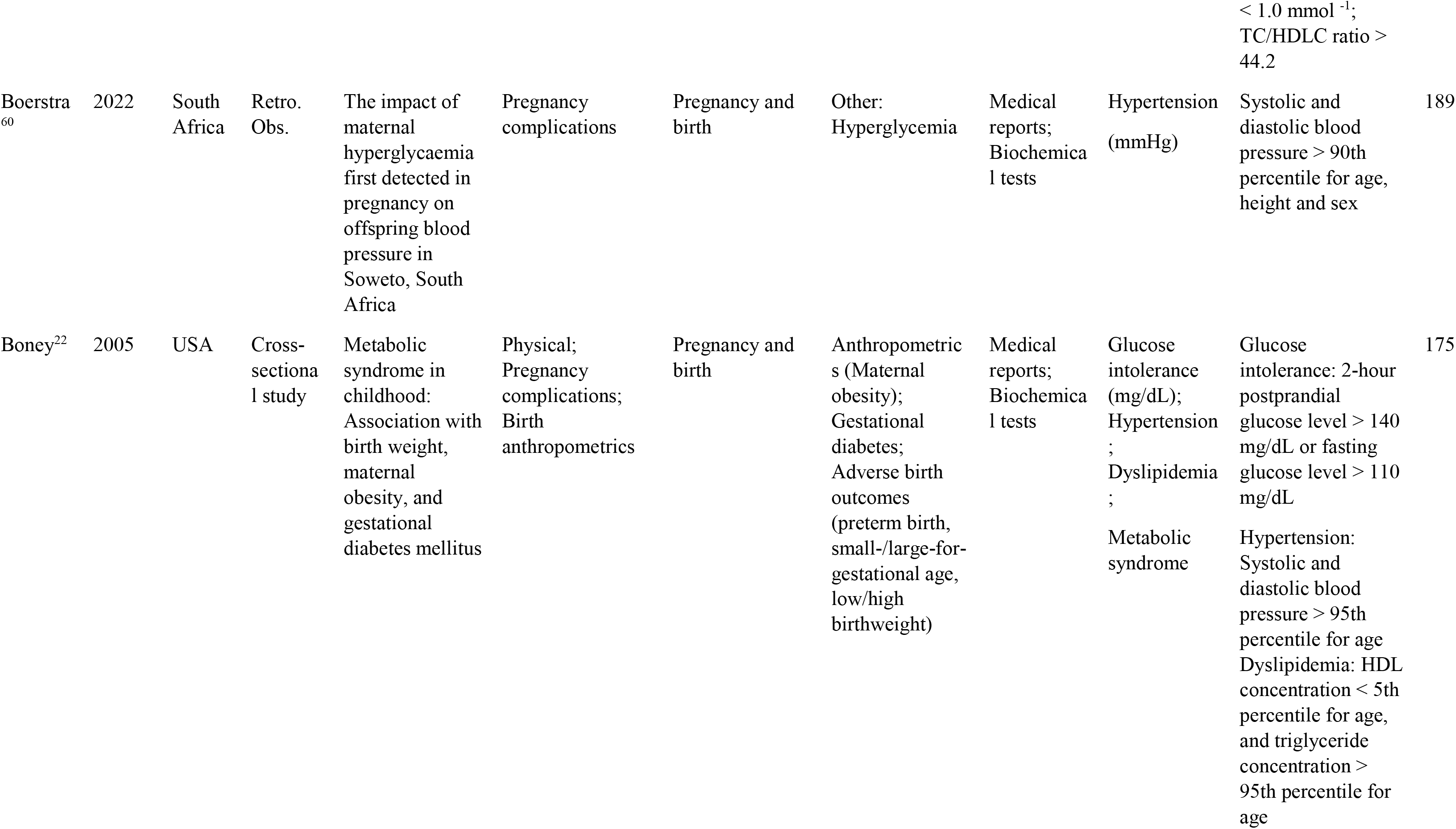
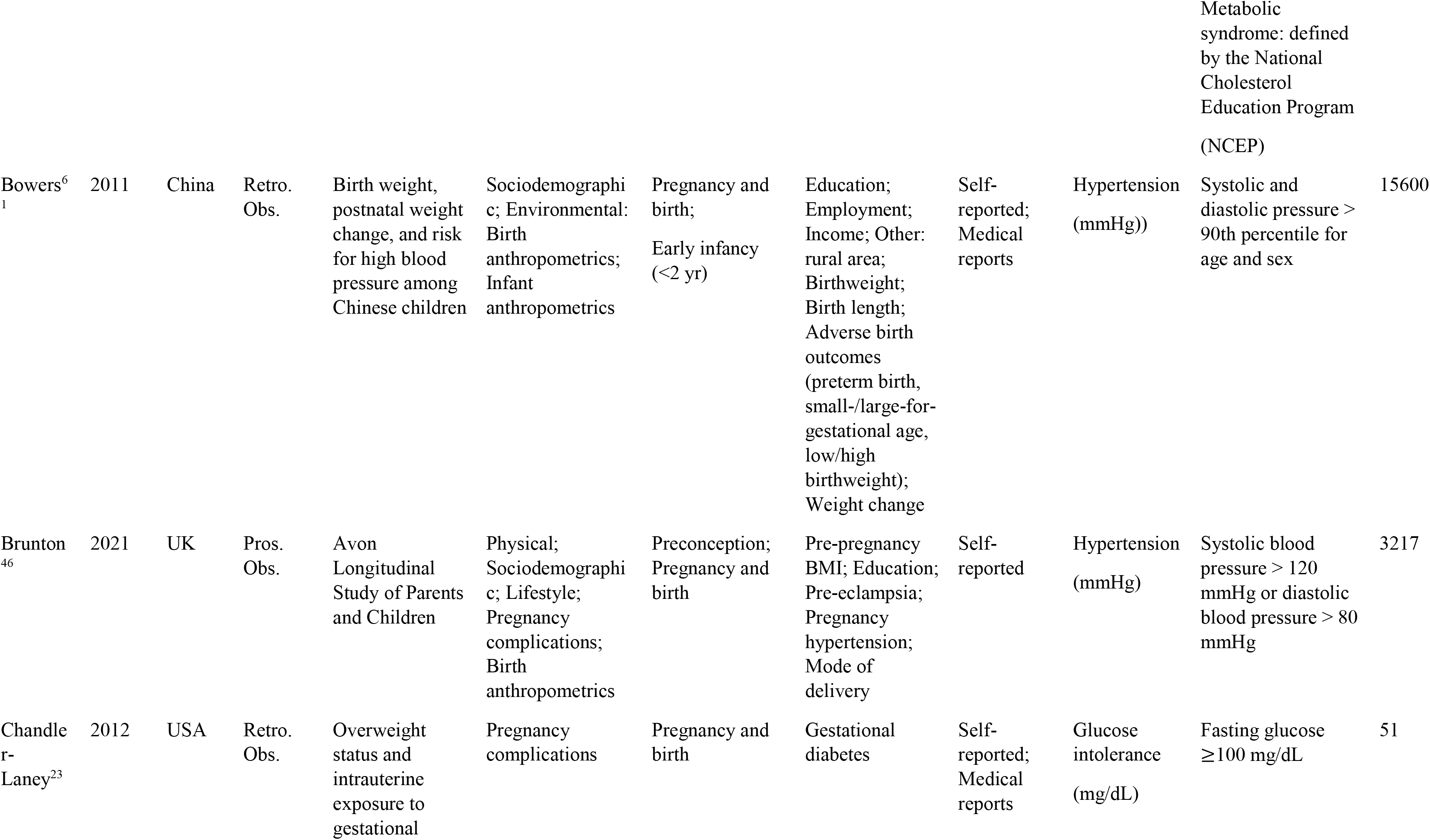
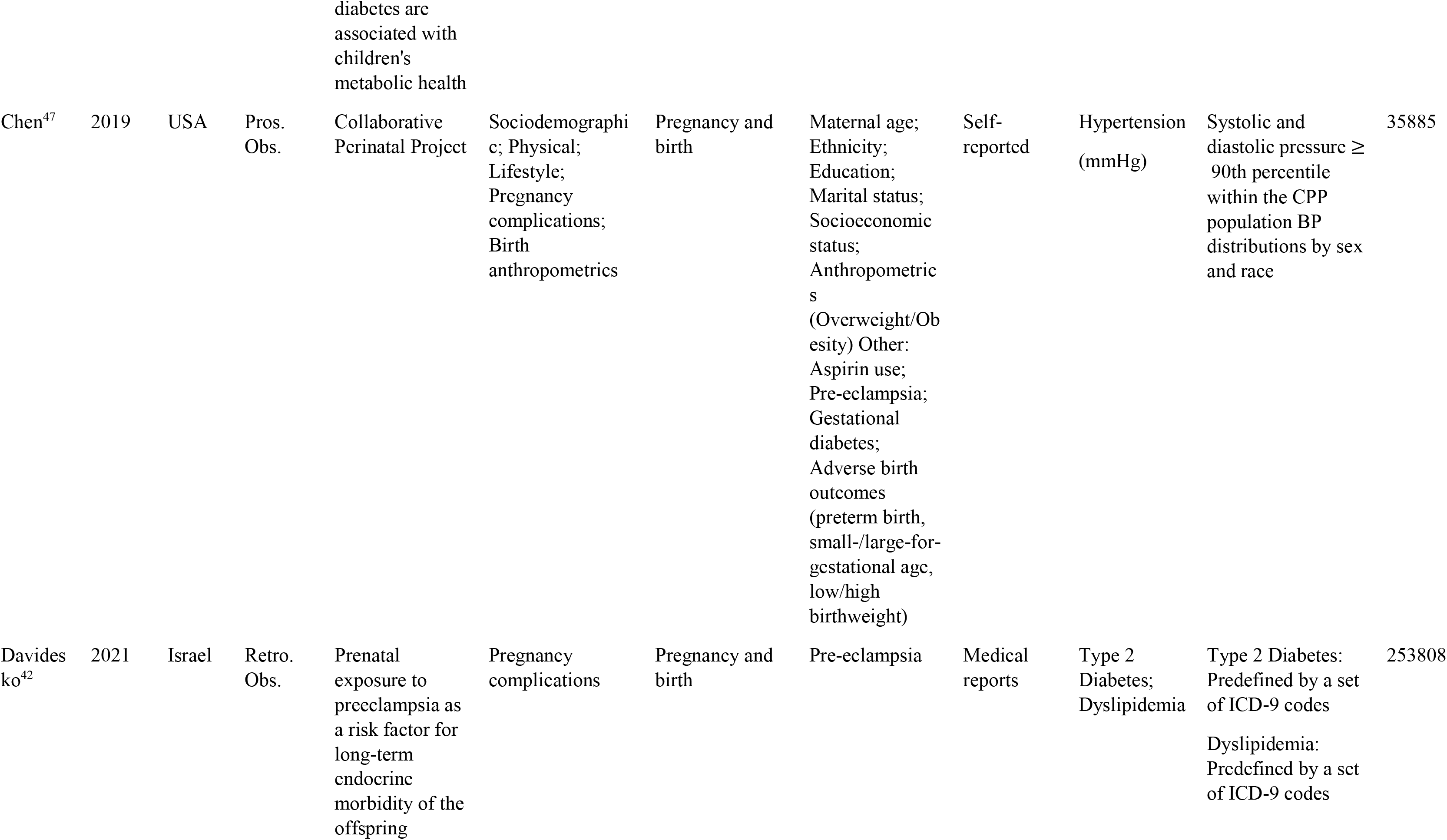
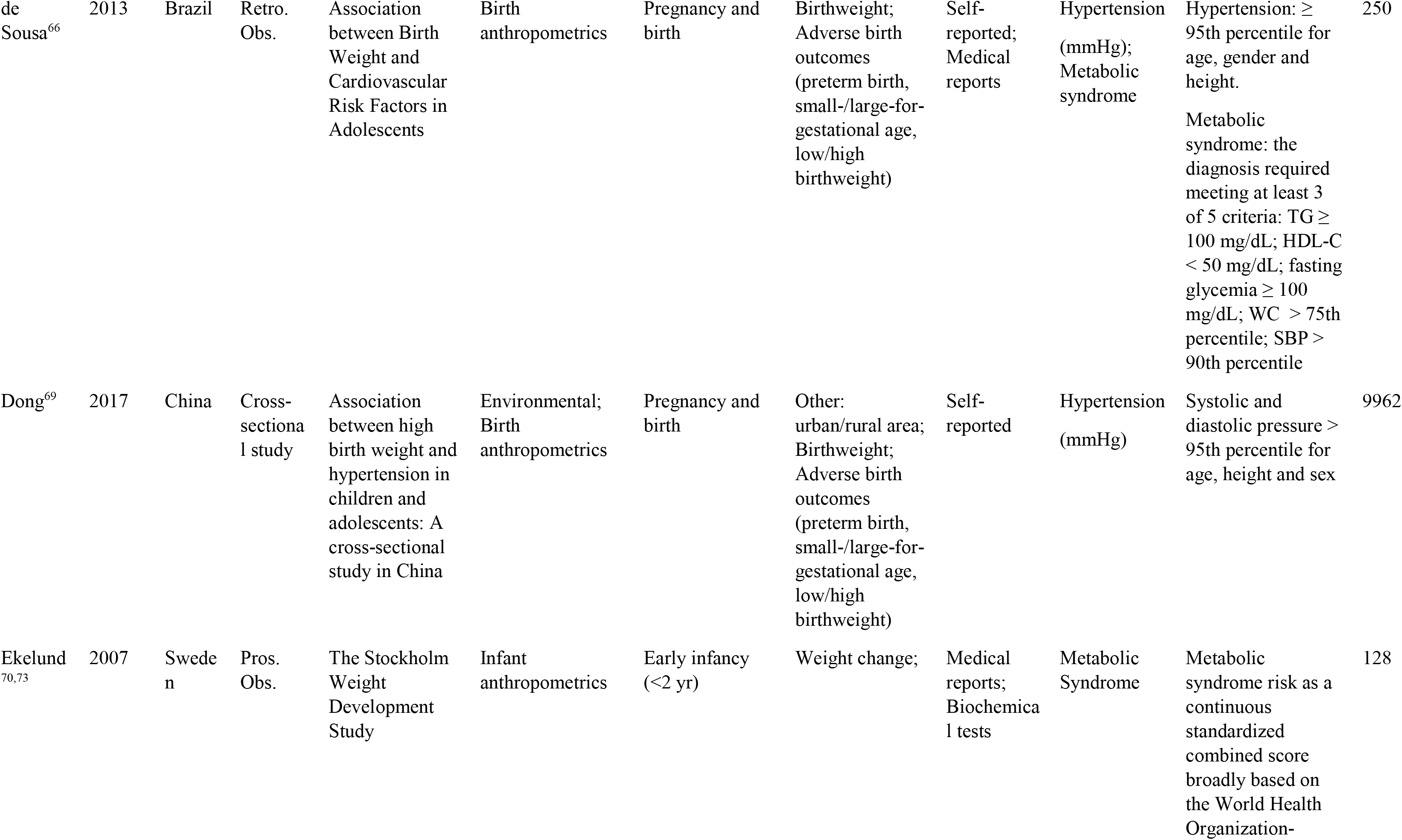
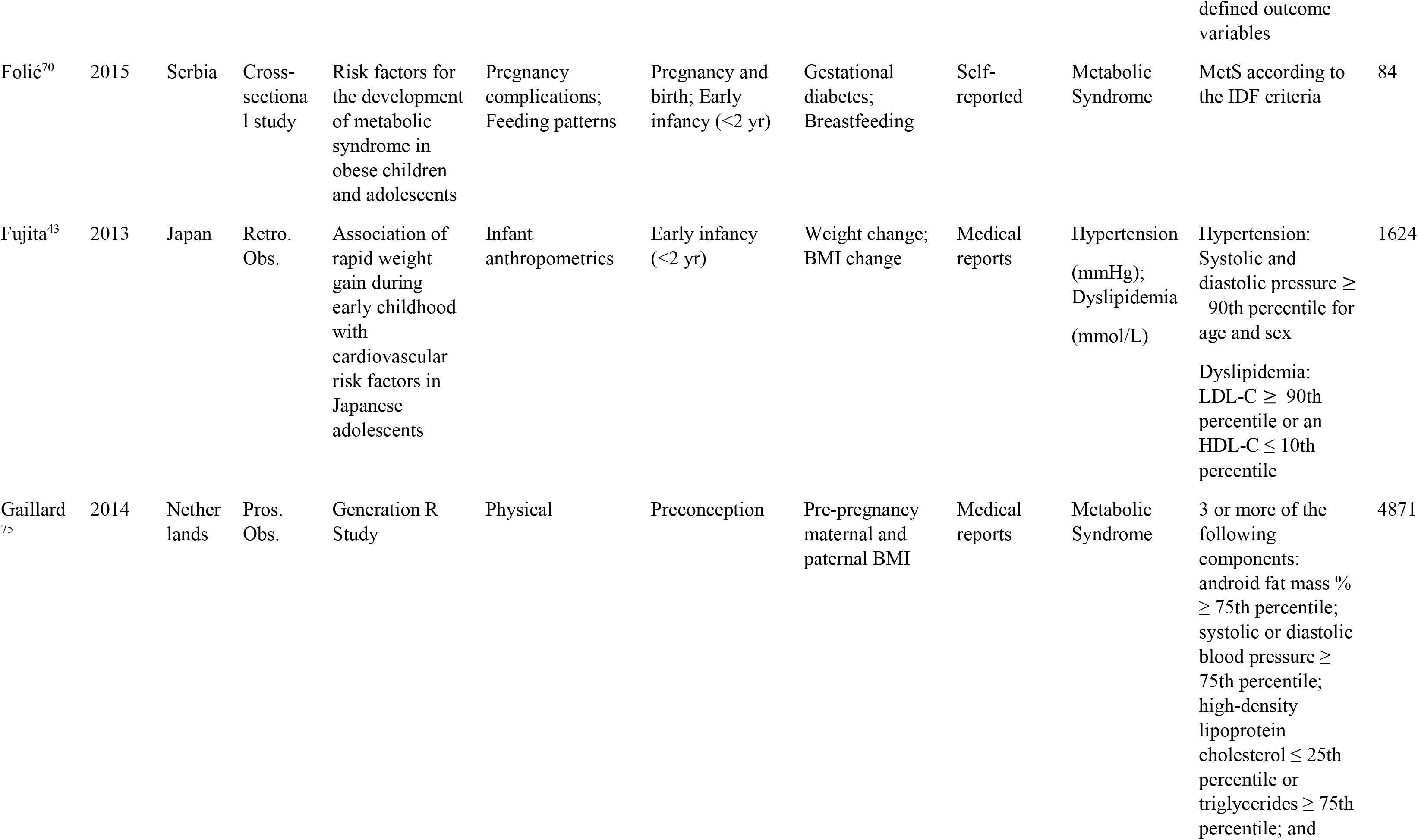
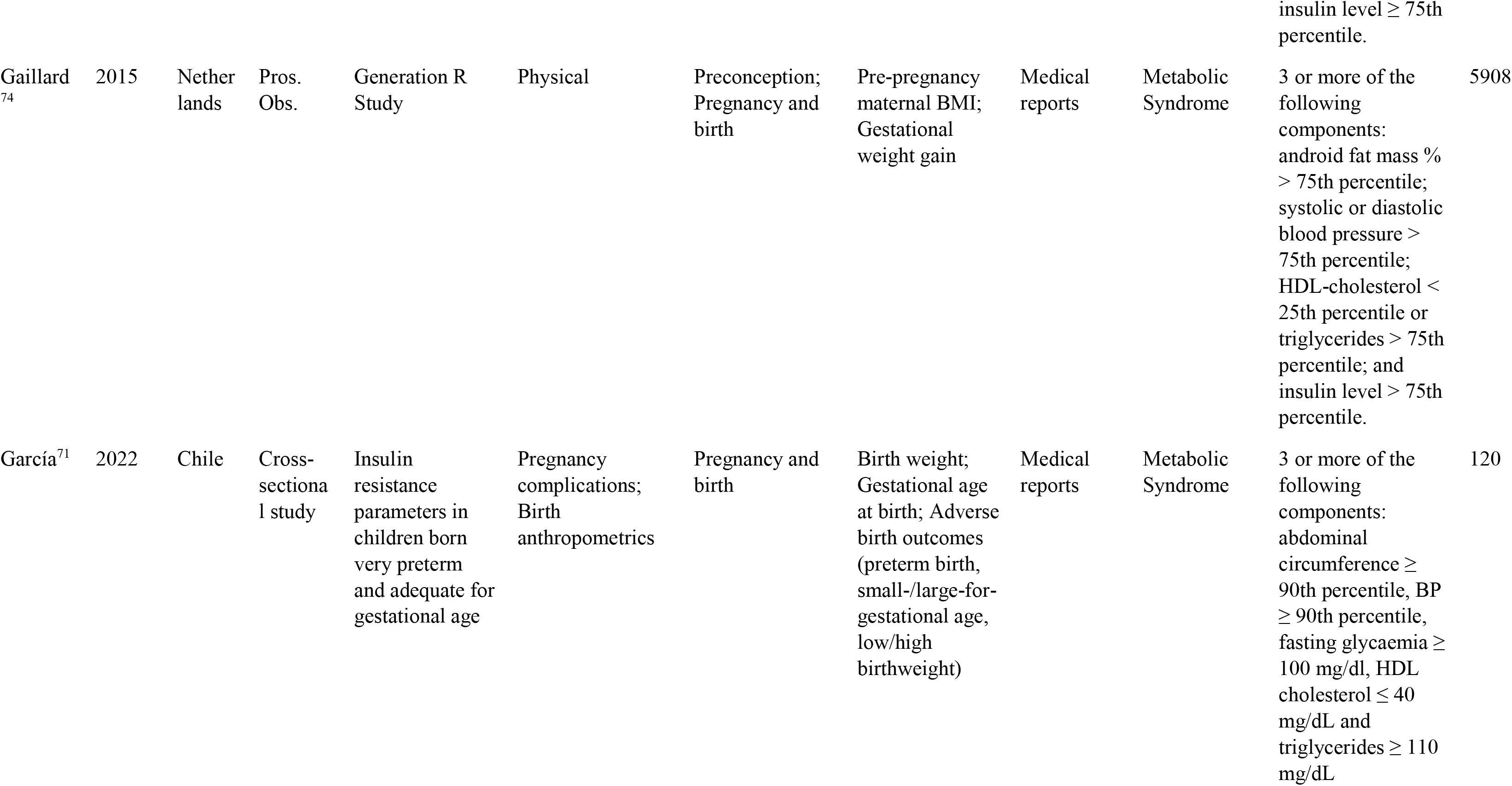
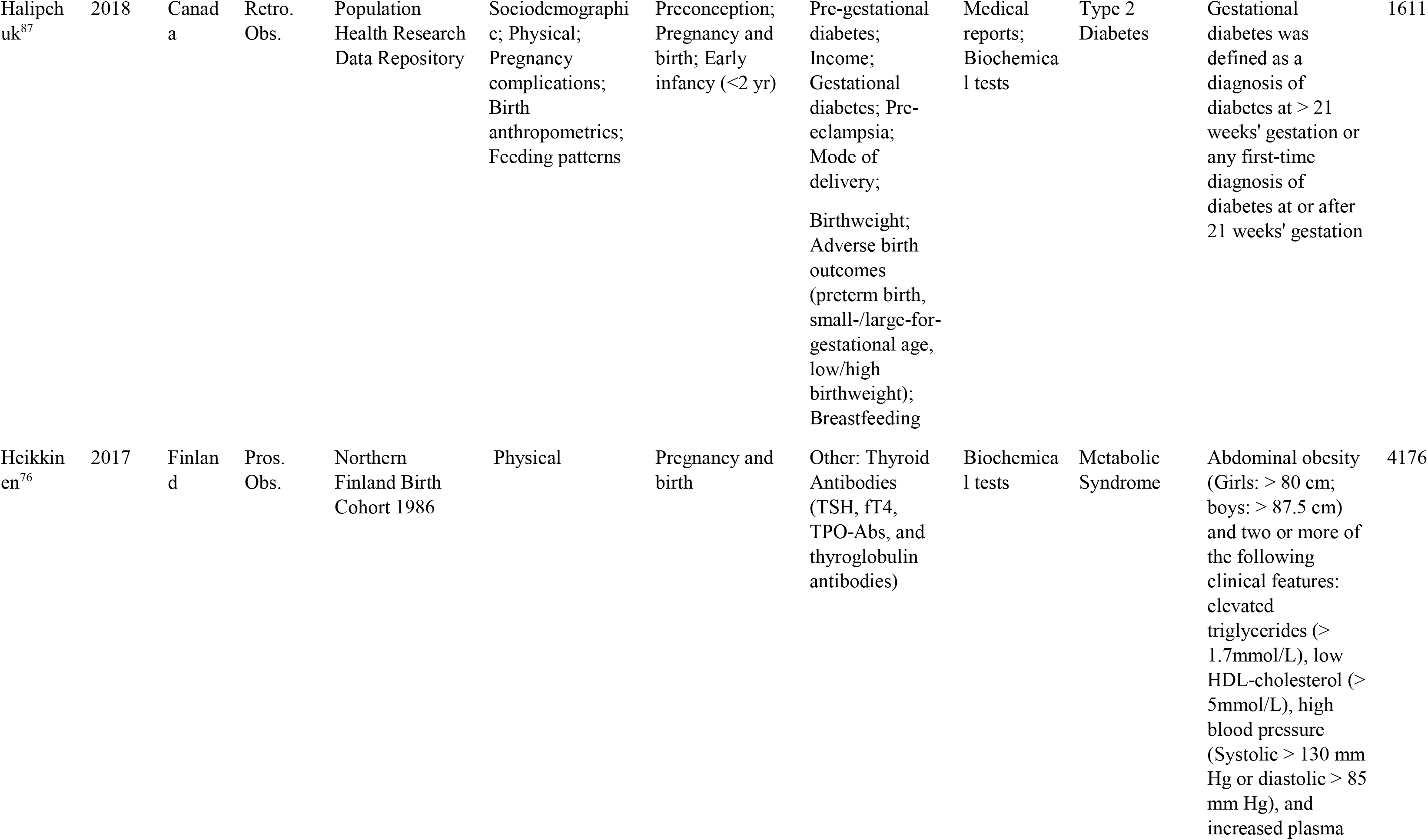
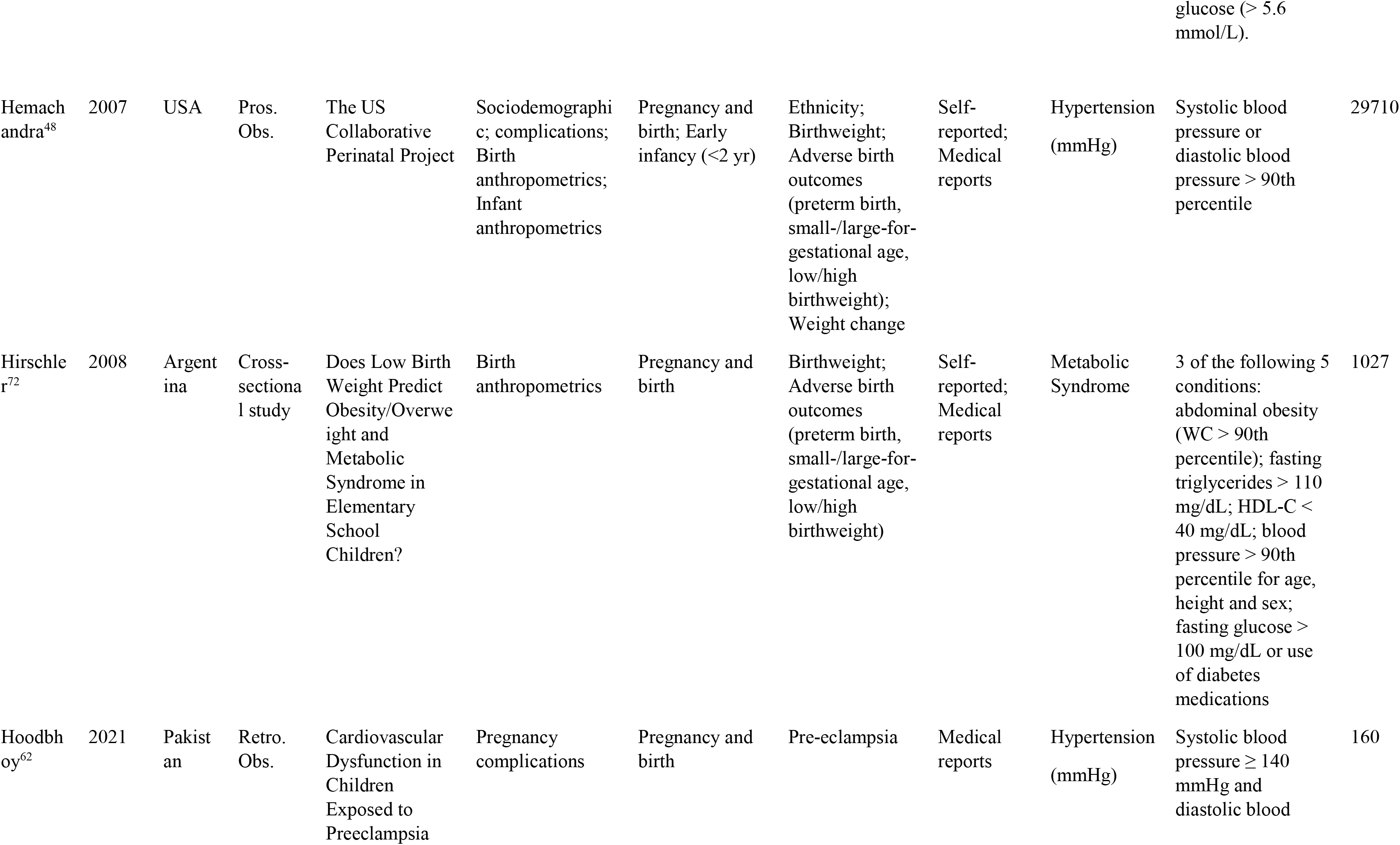
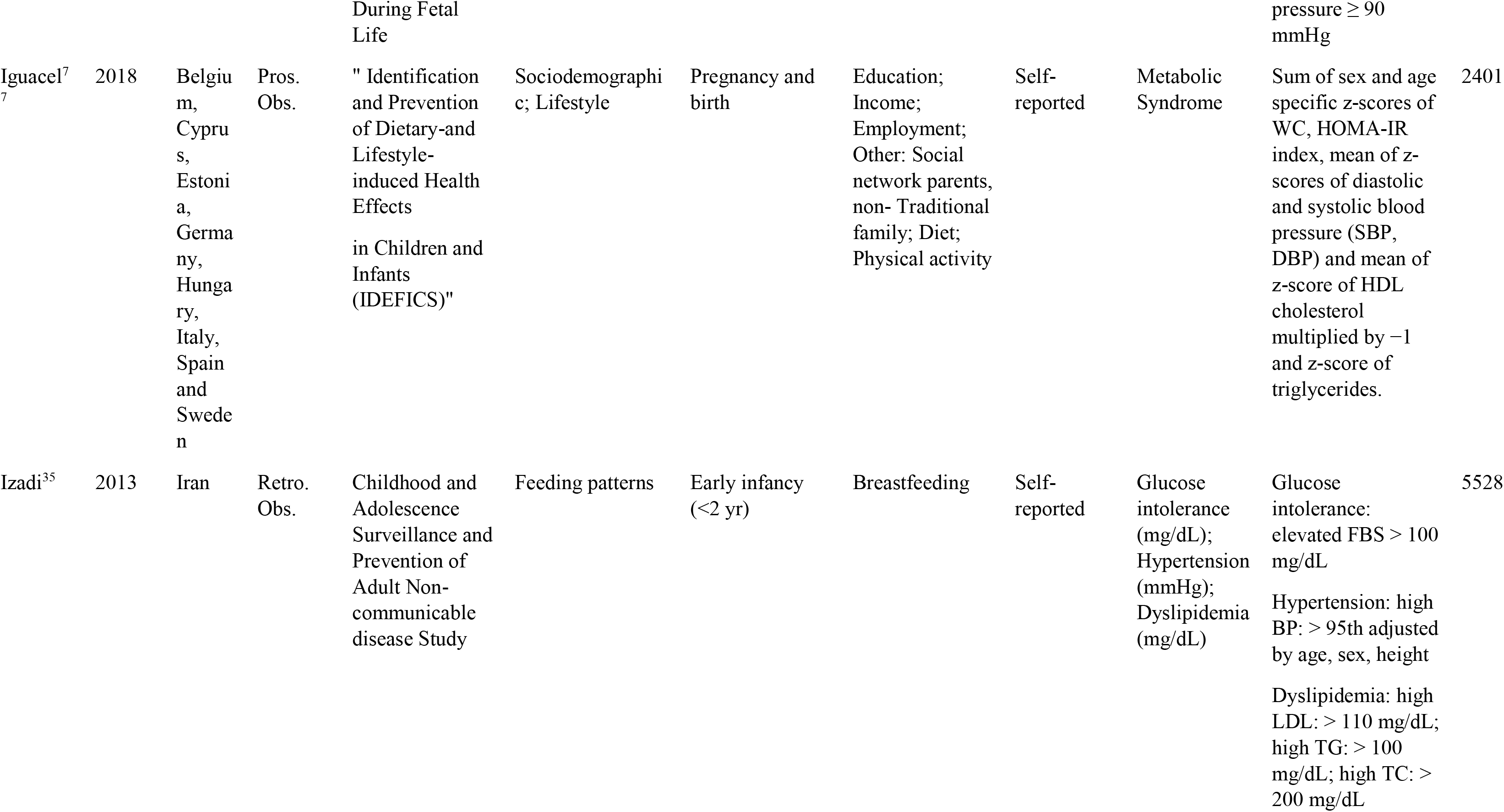
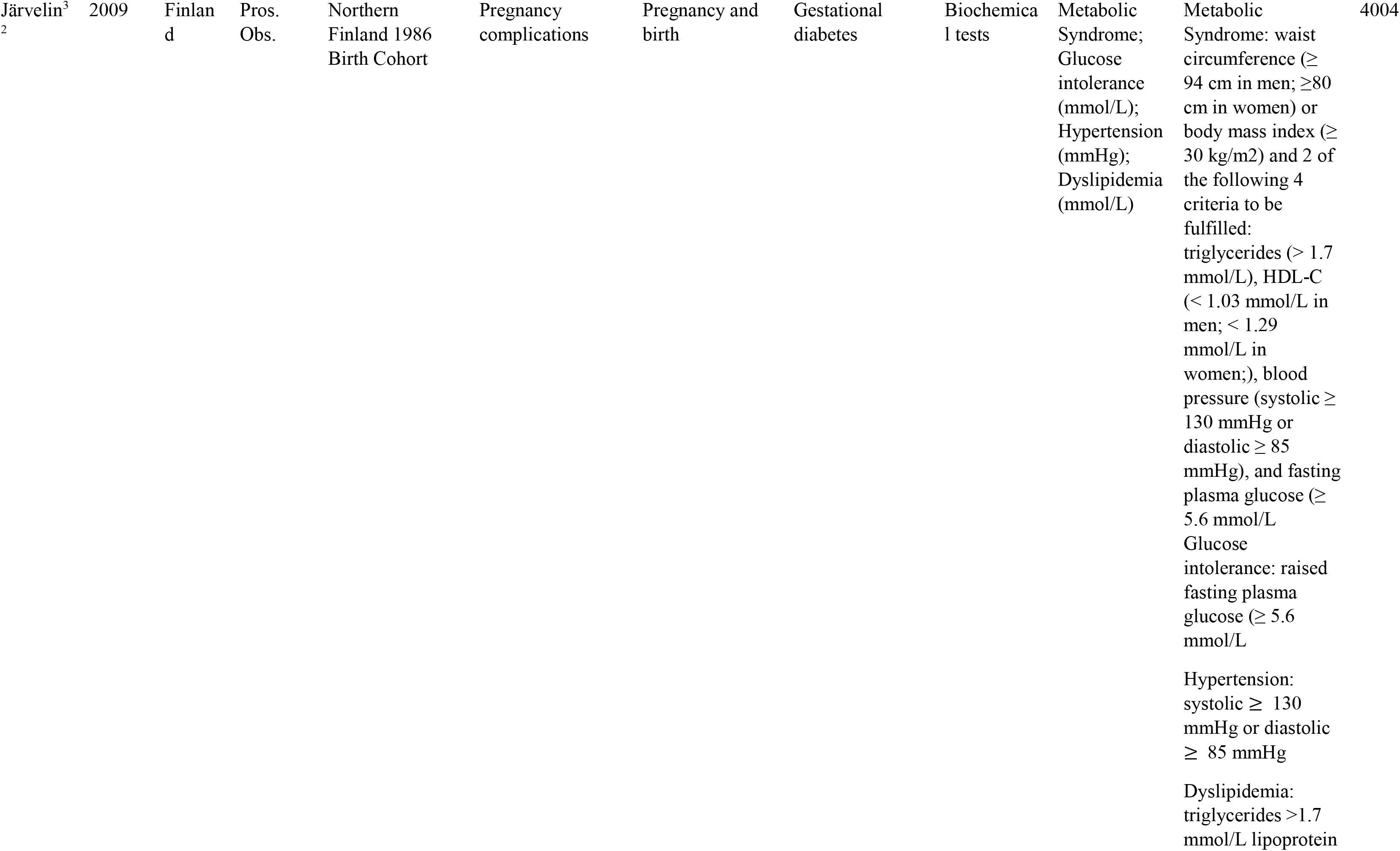
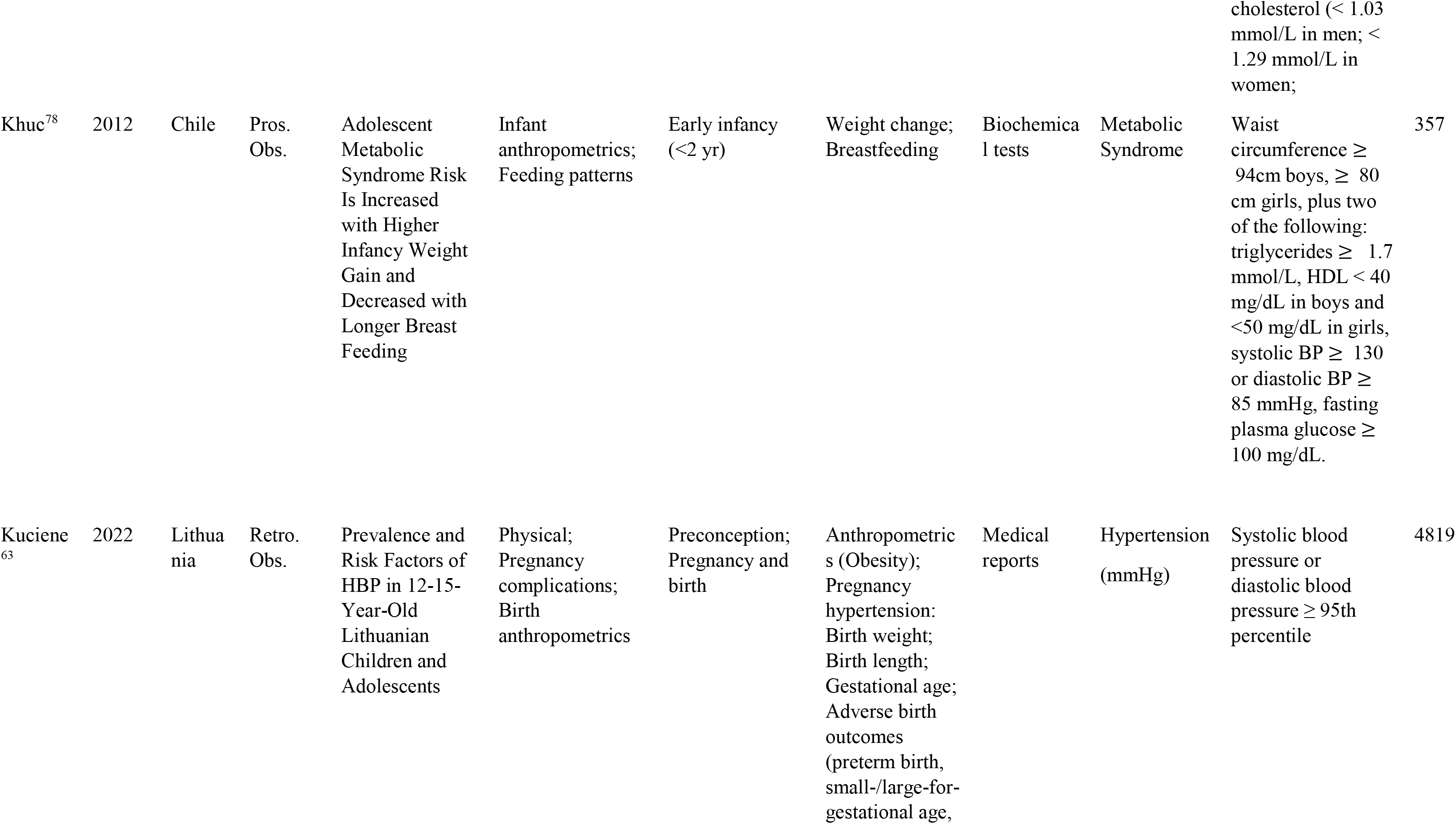
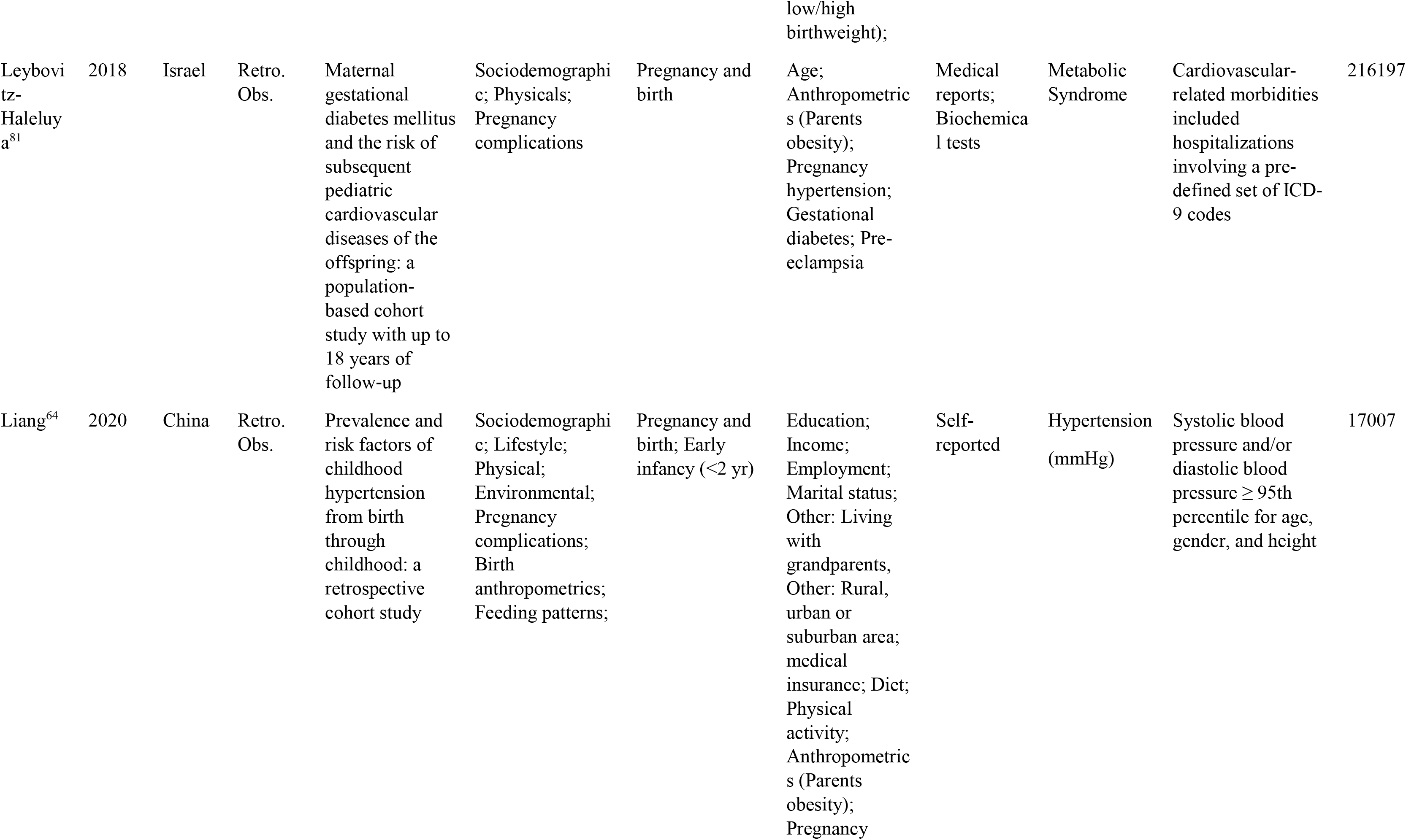
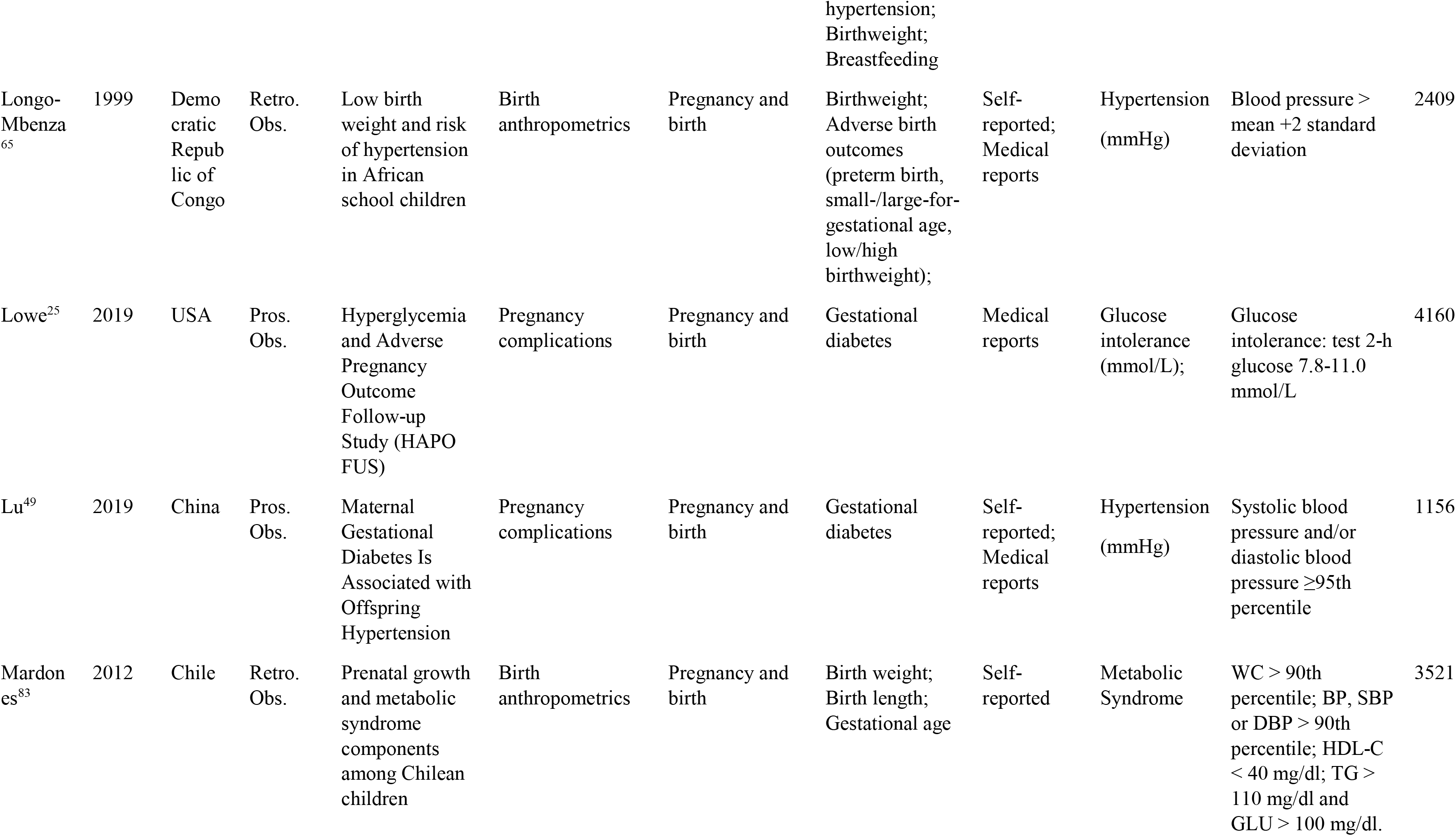
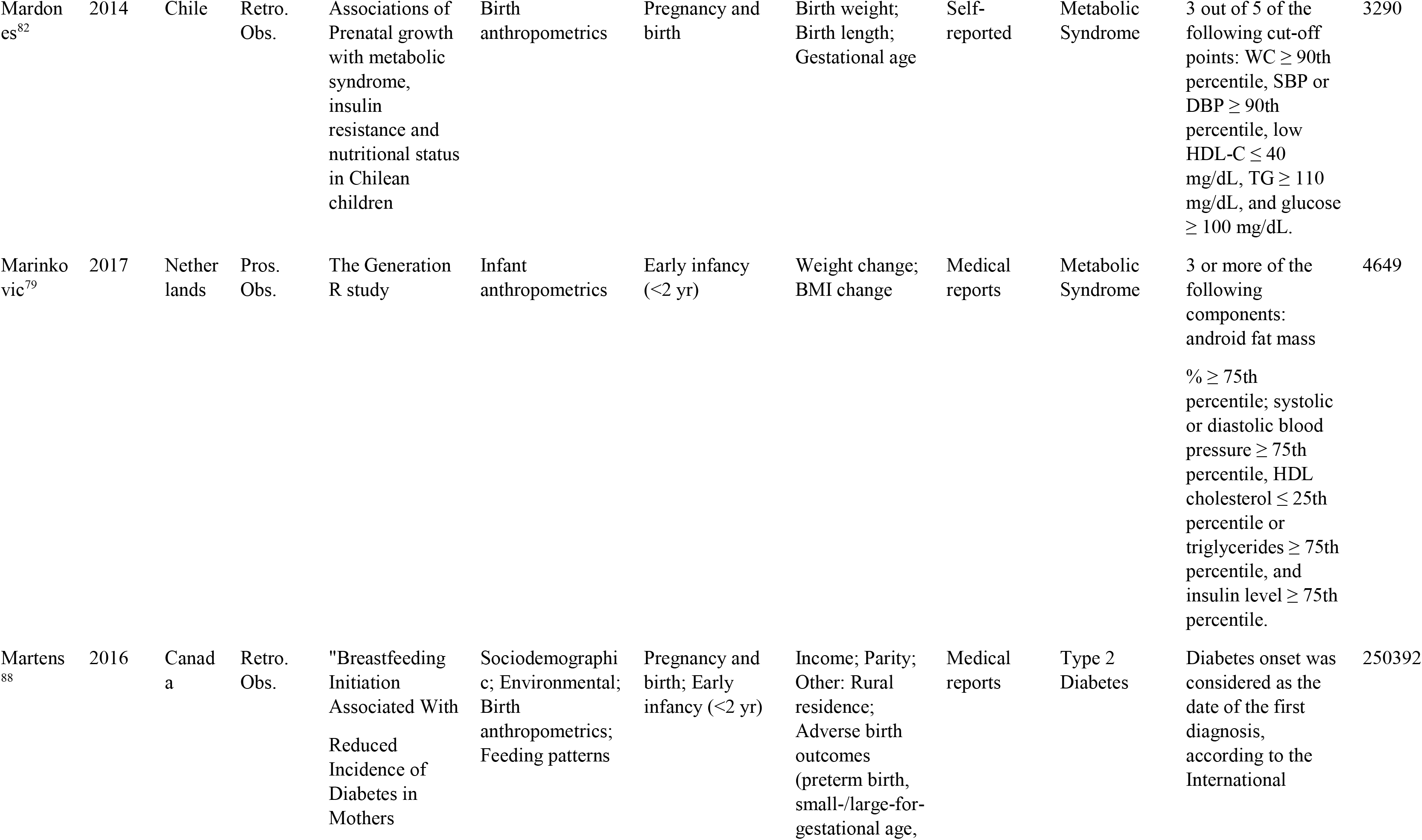
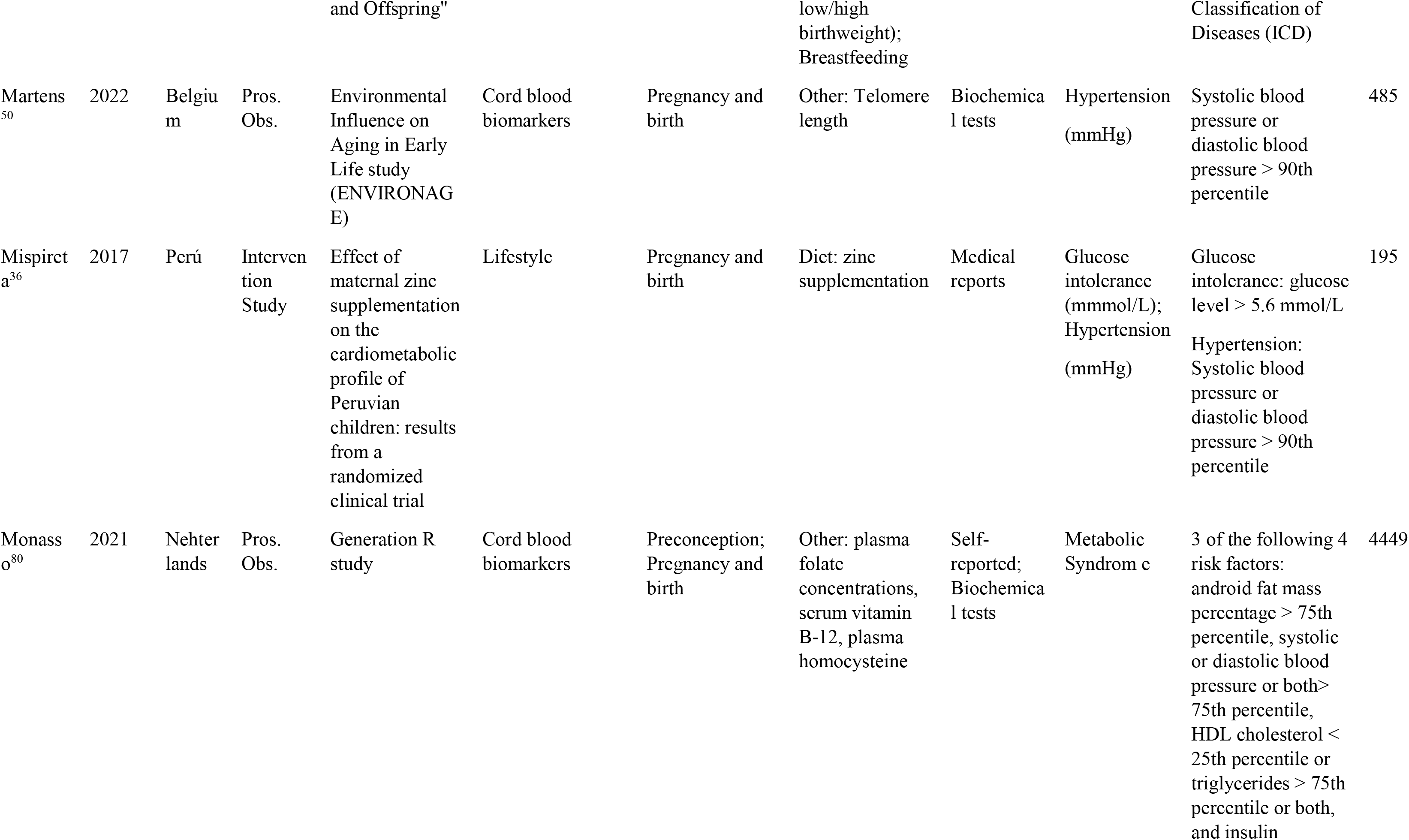
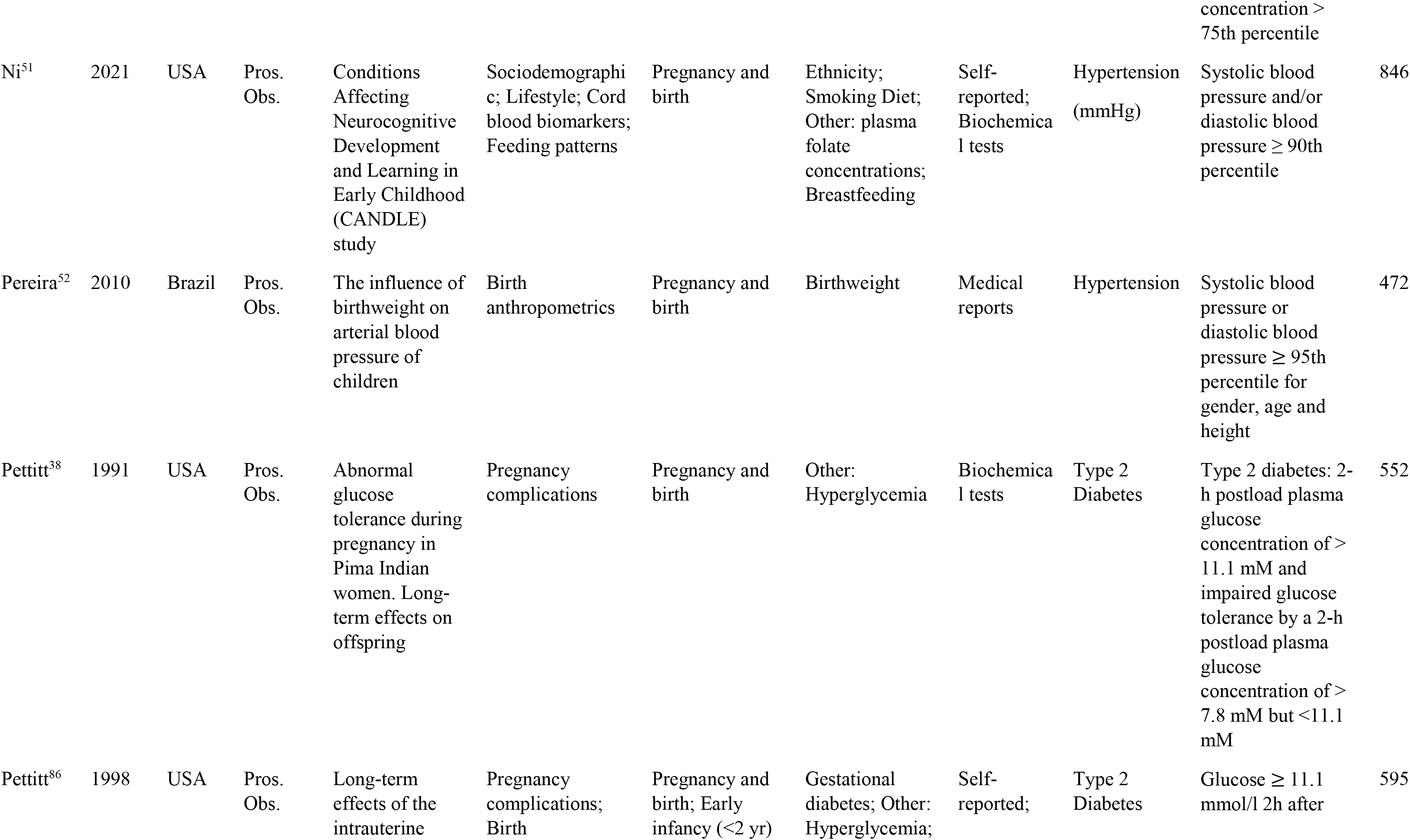
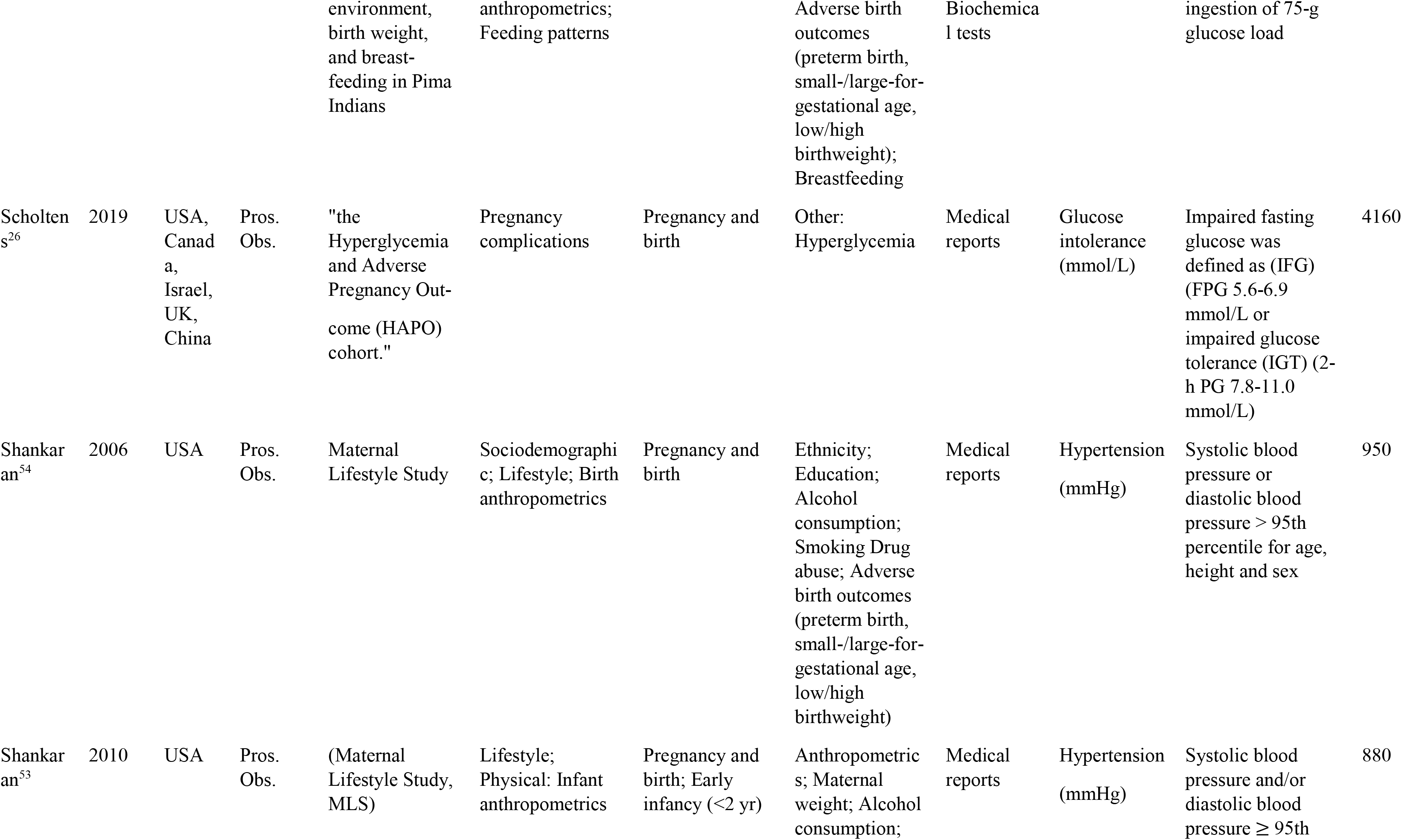
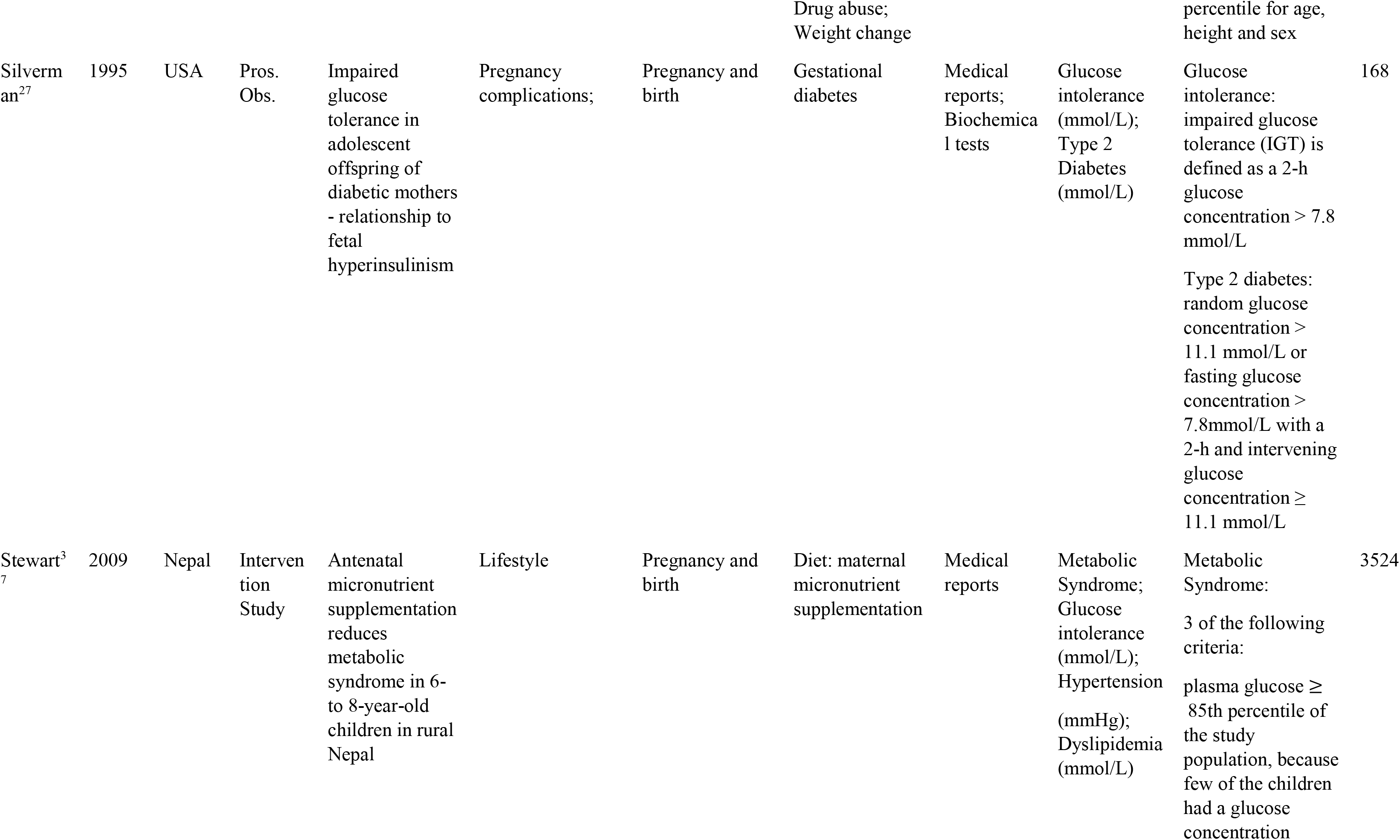
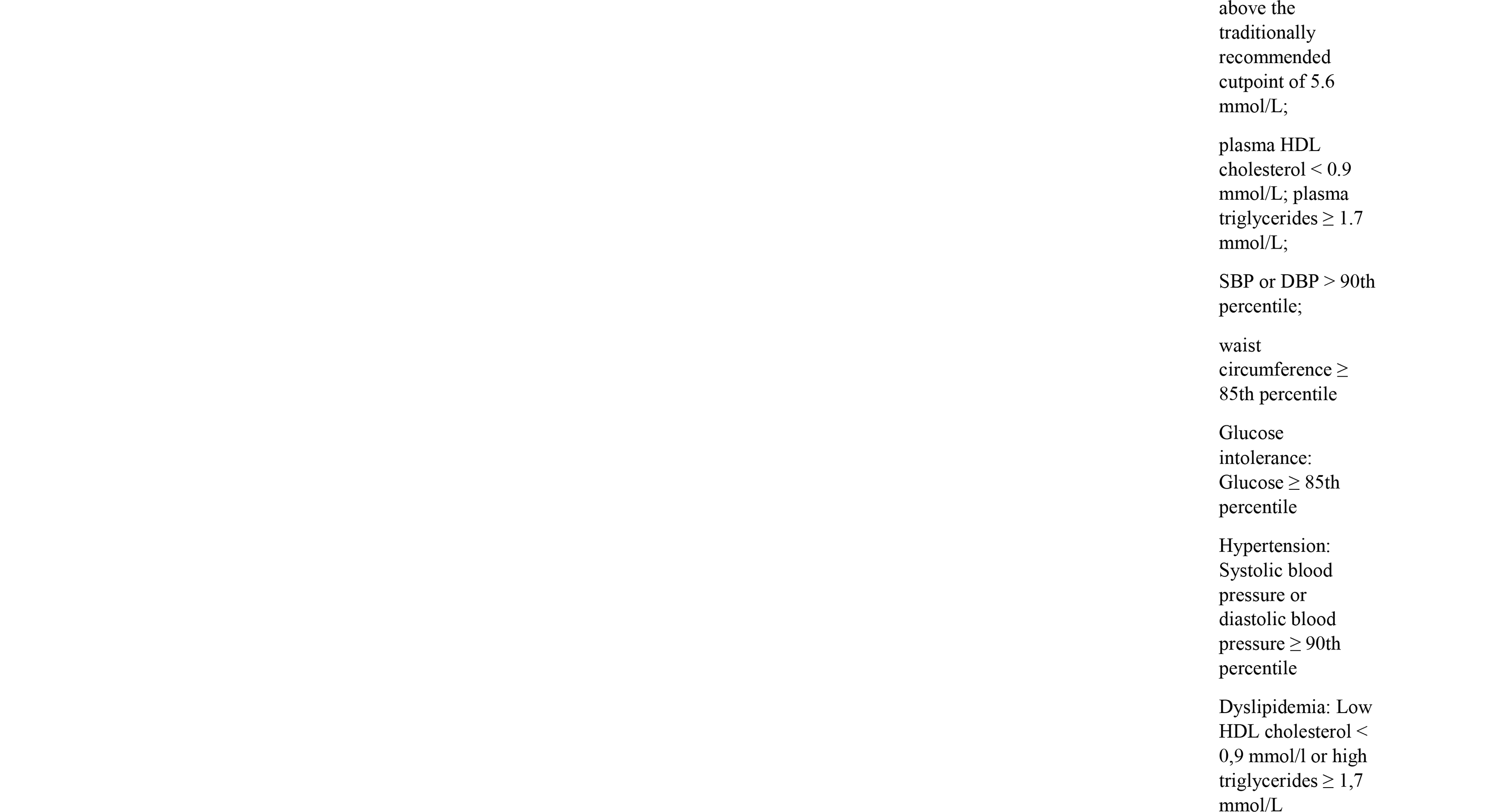
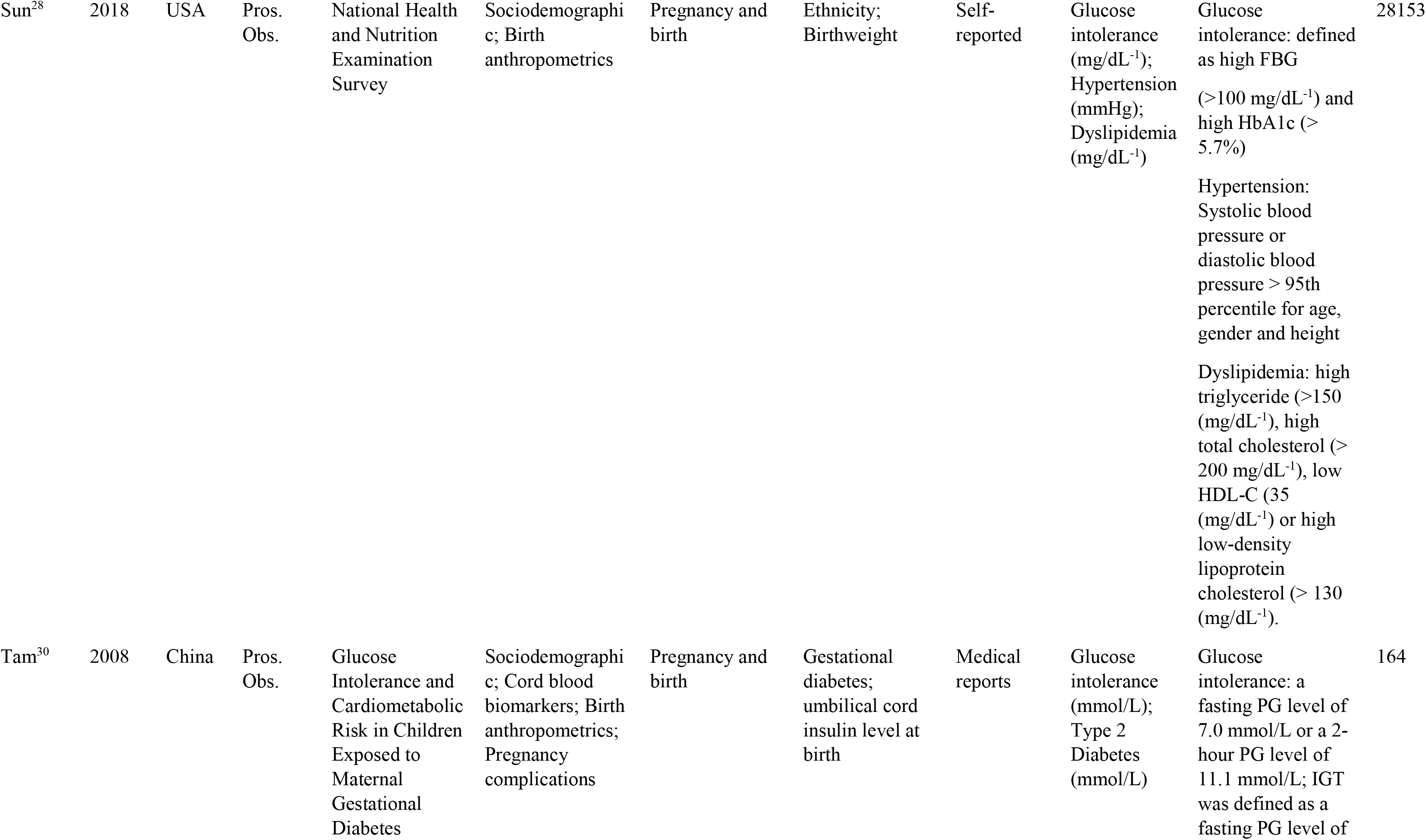
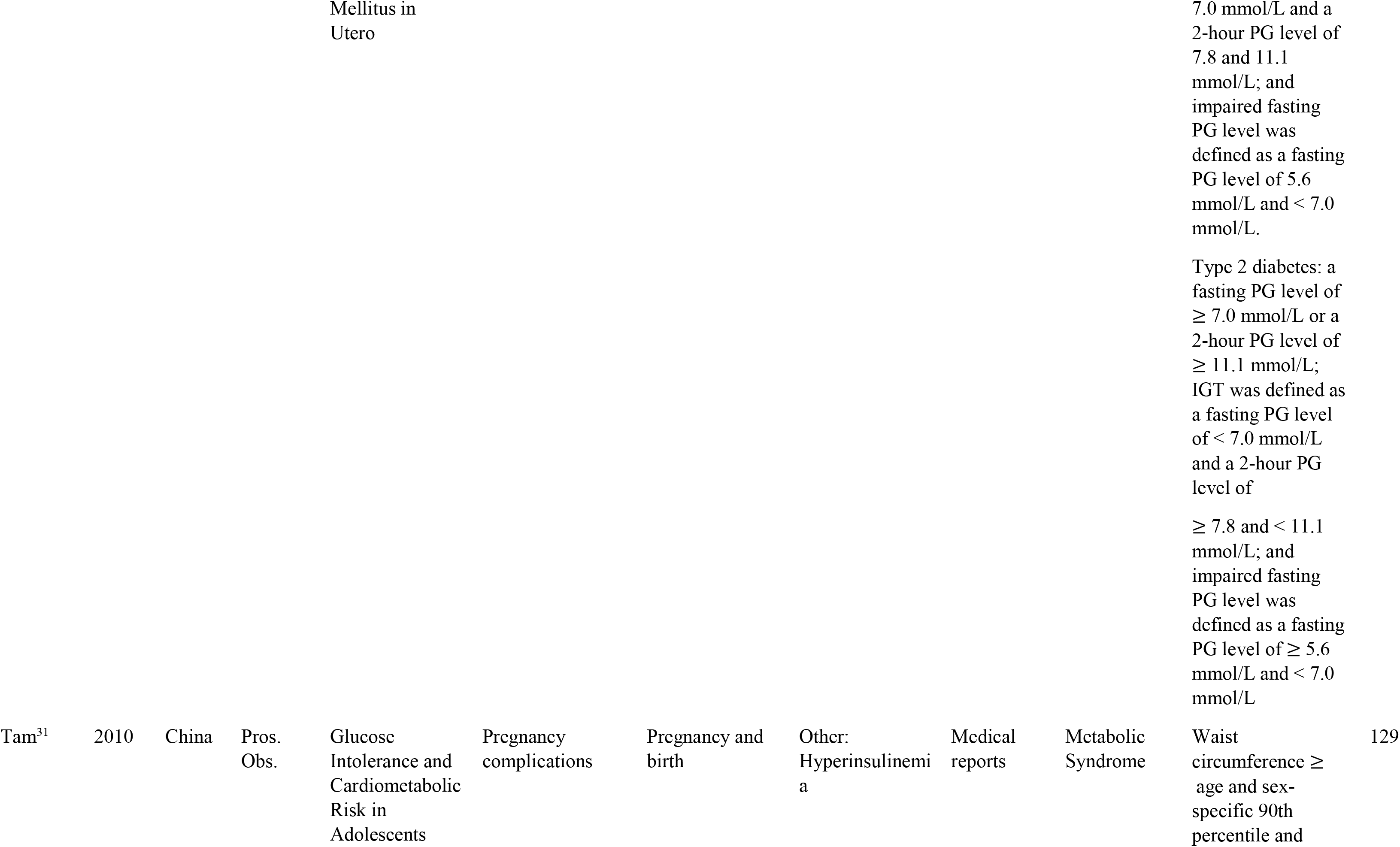
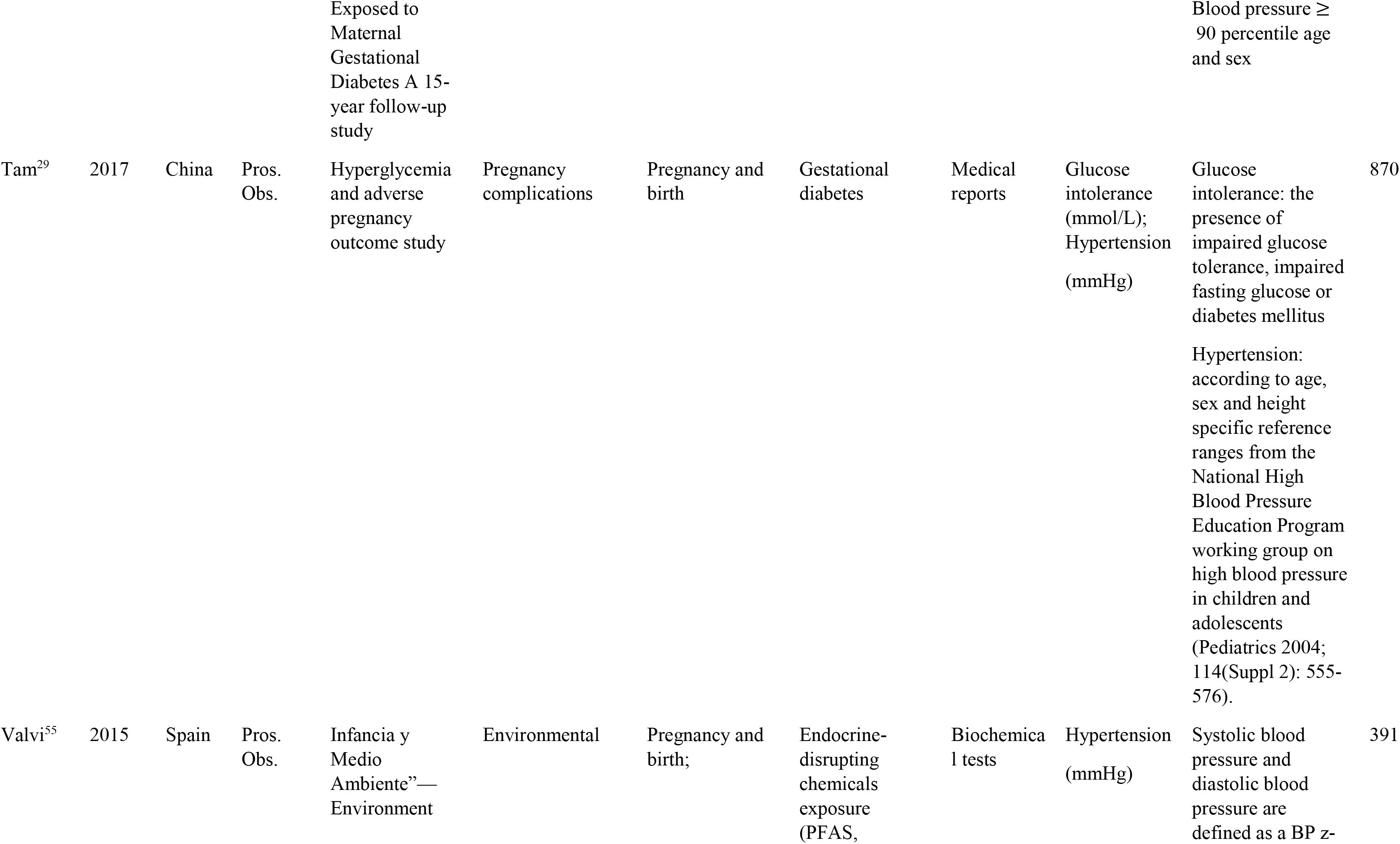
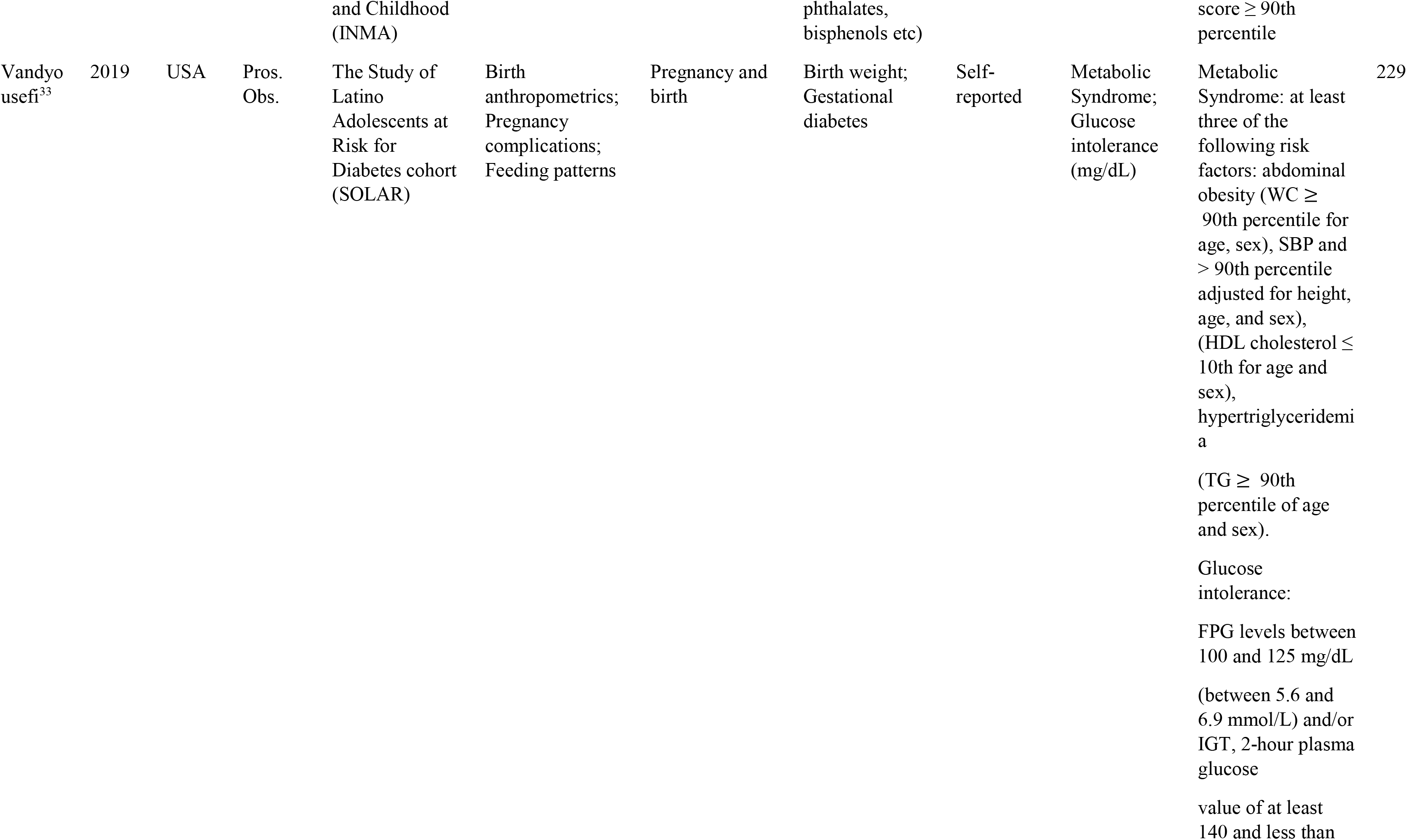
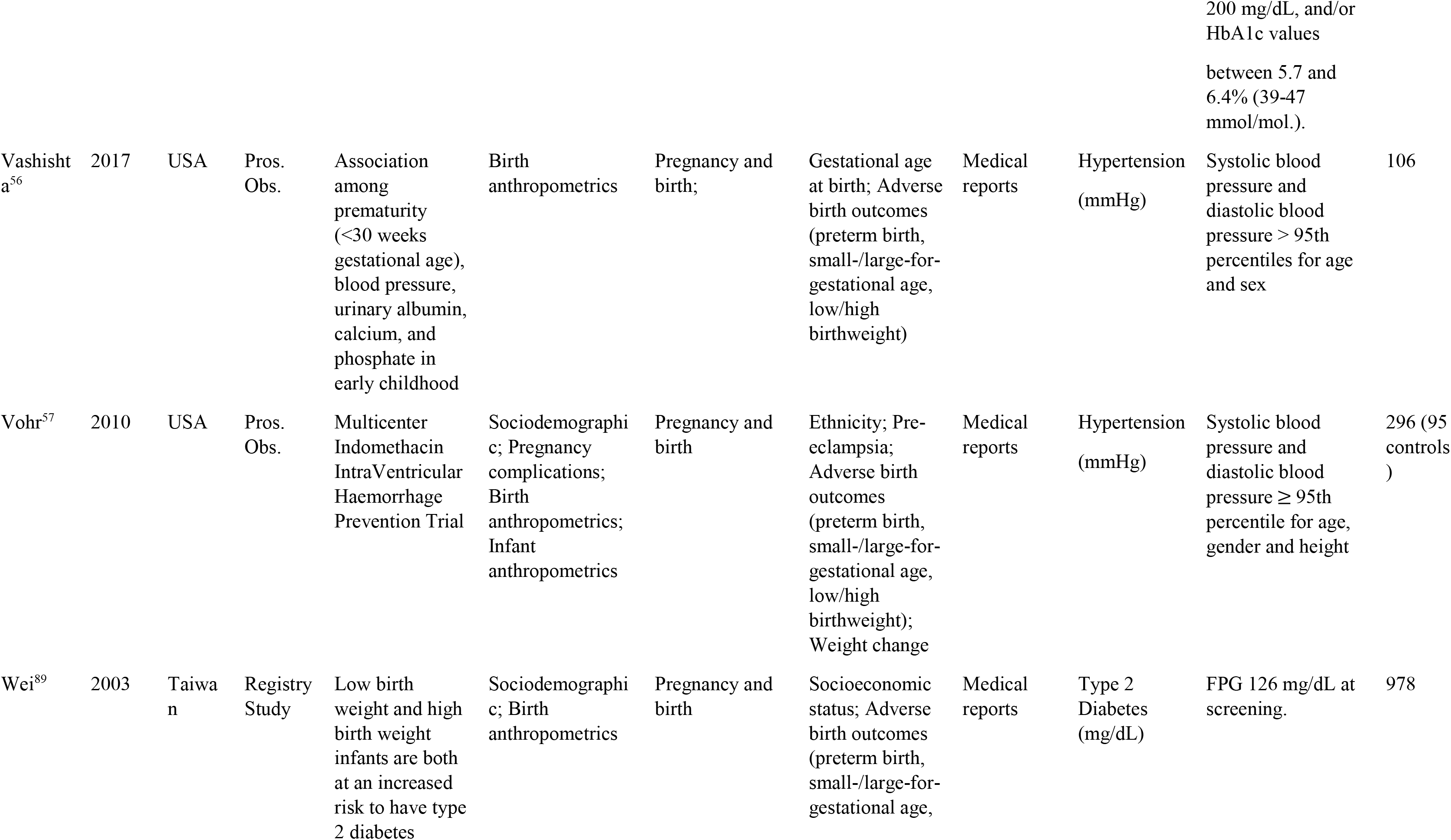
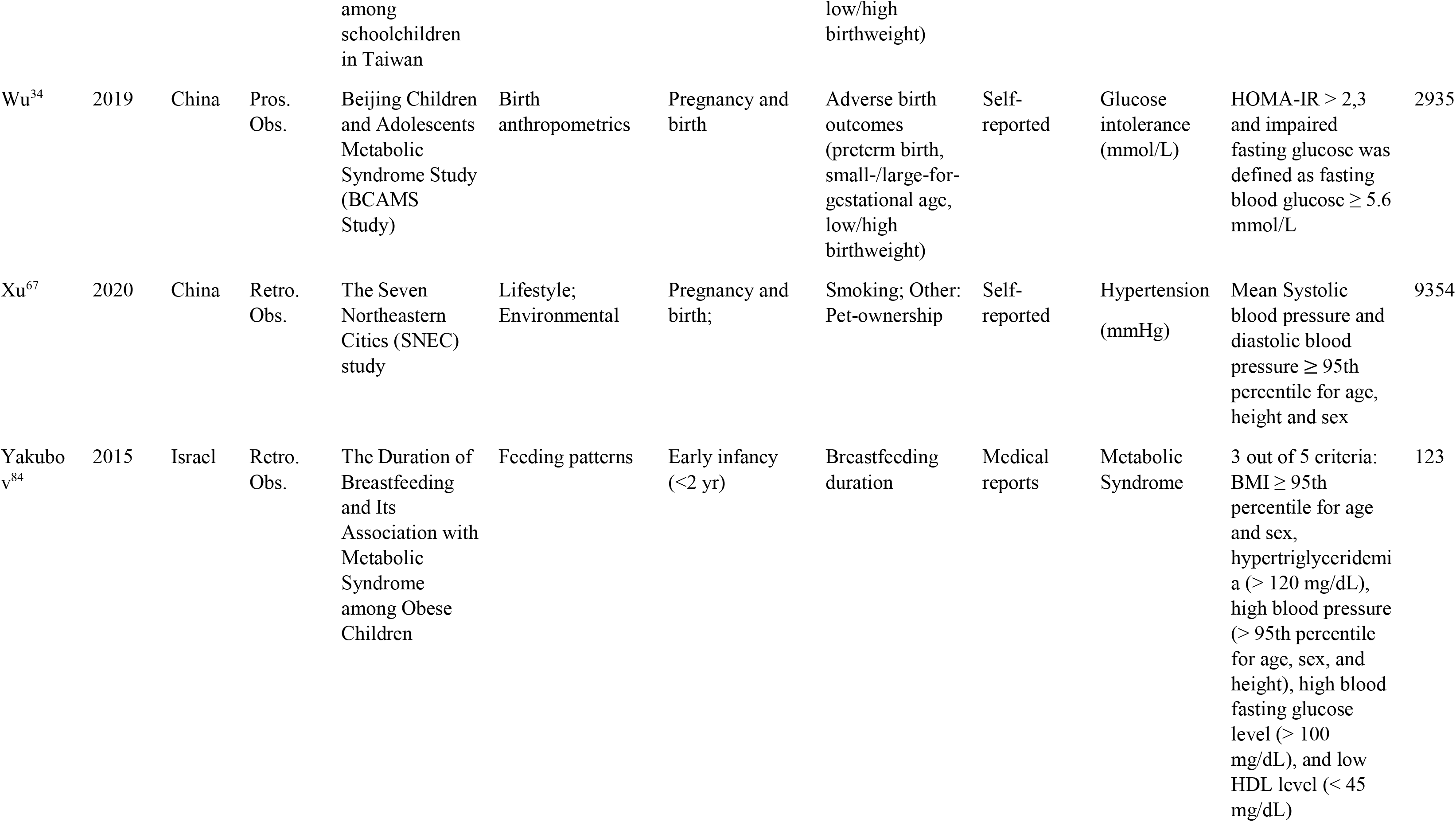
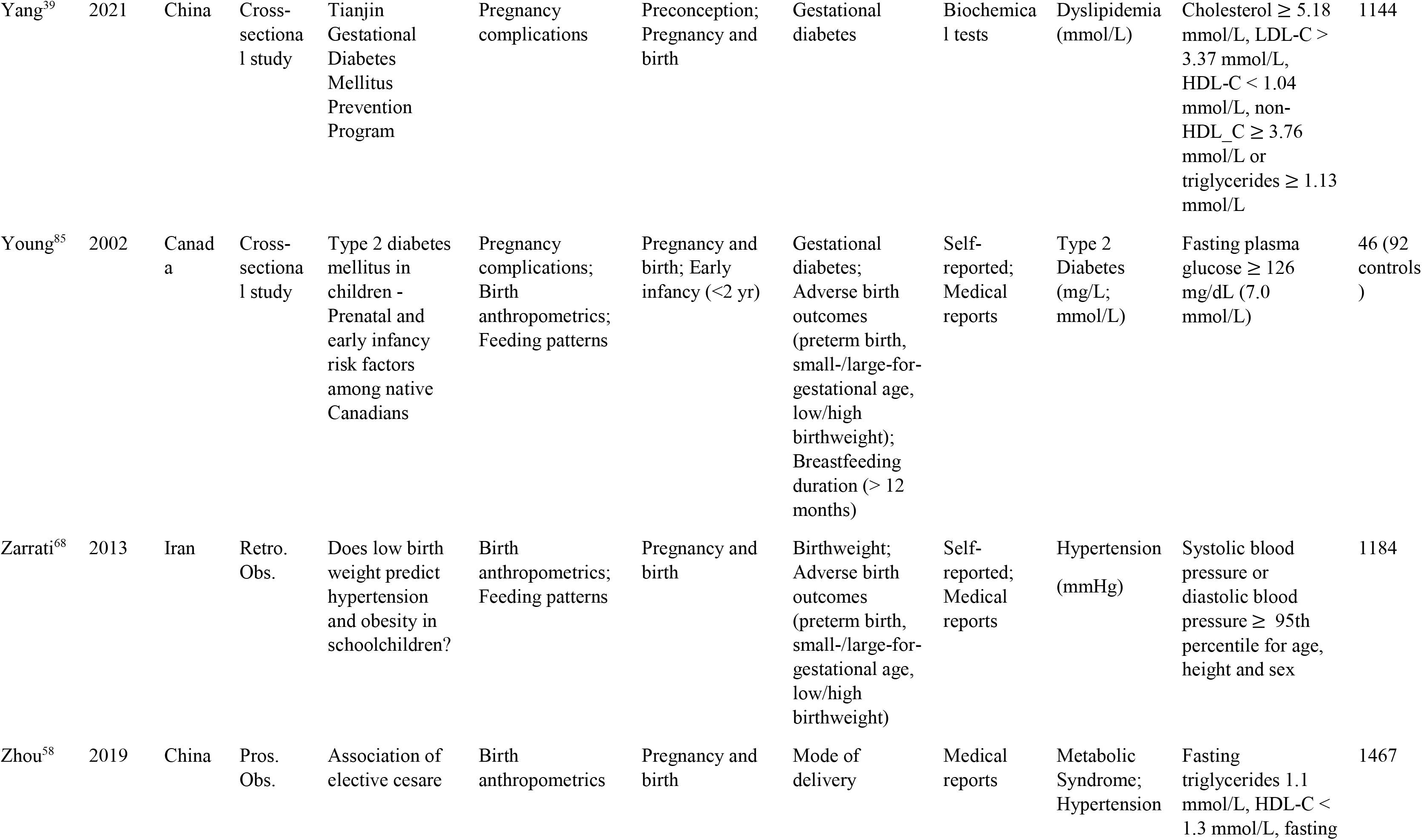
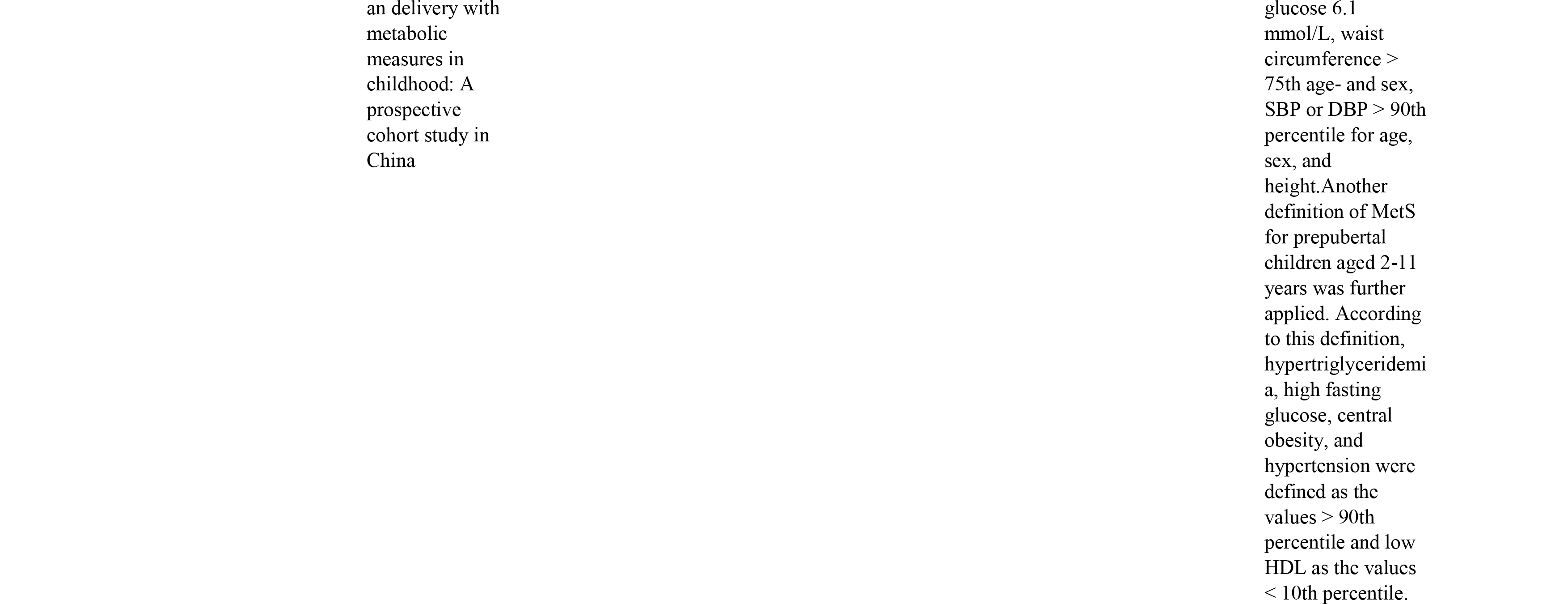
Characteristics of studies included in the systematic review.

### Associations of exposures and risk factors for cardiometabolic outcomes in childhood

We reported 229 associations between the main categories of risk factors in the first 1,000 days of life for cardiometabolic outcomes in childhood, including glucose intolerance, dyslipidemia, hypertension, metabolic syndrome, and T2D (**Table S2**). These associations were classified as positive, negative, or none. The majority of associations were positive (n=162) for all cardiometabolic risk factors, with pregnancy and birth categories exhibiting the greatest associations (Figure 1).

We found that risk factors including birth anthropometrics, pregnancy complications, cord blood biomarkers, lifestyle, physical pregnancy, infant anthropometrics, and feeding patterns categories were associated with glucose intolerance later in childhood. Of 16 evaluated studies, 2 were case-control studies^22,23^, 11 were prospective studies^24–34^, 1 was a retrospective study^35^, and 2 were RCTs^36,37^. These associations were mainly studied in pregnancy and birth period, and to a lesser extent in the early infancy period; however, investigation of the association of exposure and risk factors during preconception and later glucose intolerance was not reported in any of the reviewed studies. There were 23 associations (82% were positive associations between risk factors and glucose intolerance in children). Among them, 18 were identified positively associated pregnancy and at birth period category^22,23,25–28,32–34,37,38^ (Figure 1A).

11 unique studies reported data on dyslipidemia in children, 2 case-control studies^22,39^, 4 prospective studies^24,28,32,40^, 4 retrospective studies^35,41–43^, and 1 RCT^37^. Within these studies, 22 different risk factors included in birth anthropometrics, pregnancy complications, lifestyle, sociodemographics, physical pre- and pregnancy, infant anthropometrics, and feeding patterns categories, were investigated. Pregnancy and birth factors were the most reported period for associations with dyslipidemia. Indeed, 14 risk factors were significantly associated with an increased risk for dyslipidemia in childhood ^22,28,32,37,39^ (Figure 1B).

We identified 37 studies evaluating risk factors for hypertension in children. The majority were prospective studies (n=21) ^24,28,29,32,40,44–59^, followed by retrospective studies (n=12) ^35,41,43,60–68^, case-control studies (n=2) ^22,69^ and RCTs (n=2) ^36,37^. We identified 127 associations between early life factors and childhood hypertension, of which 66% significantly associated with increased risk for hypertension in childhood^22,28–32,37,43,45–50,53,54,56,57,61–68^ (Figure 1C).

We identified 22 studies related to childhood metabolic syndrome, including 4 case-control studies^22,70–72^, 12 prospective studies^31–33,58,73–80^, 5 retrospective studies^66,81–84^, and 1 RCT^37^. Figure 1D shows how associations were distributed during preconception, pregnancy, and infancy. There were 64 different risk factors (including physical pre- and pregnancy, birth anthropometrics, sociodemographics, pregnancy complications, cord blood biomarkers, lifestyle, infant anthropometrics, and feeding patterns categories) that associated with metabolic syndrome as outcome, from which the majority investigated risk factors during pregnancy and birth. A total of 77% of the associations were positive (49 of 64 total associations), observed in the pregnancy and birth period (40 positive associations)^22,31–33,37,58,66,70–72,74,76,77,81–83^, as well as the preconception (3 positive associations) ^74,75^ and early infancy period (6 positive associations) ^33,70,73,78,79^ were identified.

There were 9 studies that reported associations between risk factors (including physical prepregnancy, birth anthropometrics, sociodemographics, pregnancy complications, cord blood biomarkers, lifestyle, environmental, and feeding patterns categories) for developing T2D in children. Of these, 1 was a case-control study^85^, 4 were prospective studies^27,30,38,86^, and 4 were retrospective studies^42,87–89^. For this outcome, we did not find RCTs. Thirty-one associations were evaluated. Among them, 23 risk factors were positively associated with T2D^27,30,38,85–89^, 7 were not associated^42,85,88,89^, and only 1 reported a decreased risk for this outcome ^87^. Most of the associations analyzed were risk factors during pregnancy and birth periods^27,30,42,85,86,88,89^, followed by early infancy^86–88^ and the preconception period^87^ (Figure 1E).

### Preconception risk factors associated with childhood cardiometabolic outcomes

During the preconception period, 7 studies identified exposures and risk factors to childhood cardiometabolic outcomes, 6 of which were observational studies, 5 of which were prospective studies^45,46,74,75,80^, 1 of which was a retrospective study^87^, and 1 of which was a case-control study^39^. No RCTs were reported. We identified 4 risk factors measured during preconception, 3 of which were associated with a higher cardiometabolic risk in children. The prepregnancy maternal BMI and the prepregnancy paternal BMI as exposures were linked positively to cardiometabolic risk factors^39,45,46,74,75^. Moreover, pregestational diabetes and diabetes were also positively associated with cardiometabolic risk^87^. Finally, 1 study reported that higher concentrations of plasma folate, serum vitamin B12 and plasma homocysteine were associated with metabolic syndrome^80^ (**Table S2**). Our current review of the available evidence shows that physical prepregnancy risk factors during the preconception period, which includes both paternal and maternal exposures, were mostly associated with metabolic syndrome and hypertension in childhood^45,46,74,75^ (Figure 2A).

**Figure 2.**
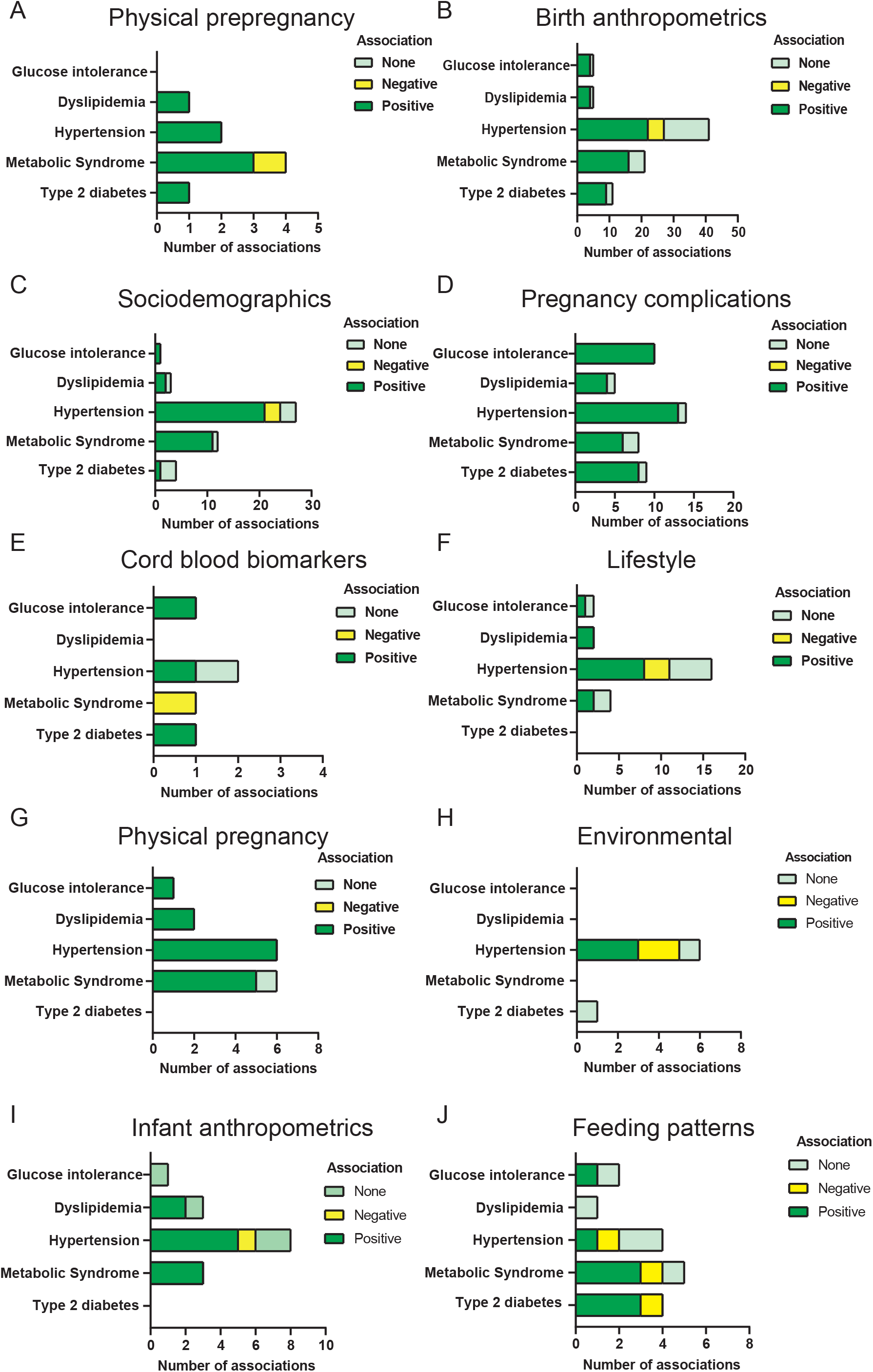
Positive, negative, and null associations between risk factors physical prepregnancy (A), birth anthropometrics (B), pregnancy complications (C), sociodemographic (D), cord blood biomarkers (E), lifestyle (F), physical pregnancy (G), environmental (H), infant anthropometrics (I), feeding patterns (J) and cardiometabolic outcomes: glucose intolerance, dyslipidemia, hypertension, metabolic syndrome and type 2 diabetes.

### Pregnancy and birth risk factors associated with childhood cardiometabolic outcomes

The vast majority of articles included in this study (58 of 68) reported on associations between pregnancy and birth exposures and cardiometabolic risk factors in childhood. Of those, 48 were observational studies (31 prospective studies^25–34,38,44–58,74,76,77,80,86^ and 17 retrospective studies^41,42,60–68,81–83,87–89^), 8 were case-control studies^22,23,39,69–72,85^, and 2 were RCTs^36,37^. Risk factors identified during pregnancy and birth period were the most frequently studied, with 197 (86%) associations for all cardiometabolic outcomes. 7 categories of risk factors were evaluated: birth anthropometrics, sociodemographics, pregnancy complications, lifestyle, physical, environmental, and cord blood biomarkers (**Figure S2B**). The most frequently assessed characteristics were birth anthropometrics (n=75), sociodemographic data (n=47), and pregnancy complications factors (n=34). Concerning the birth anthropometric category, we found that the majority of the associations were positively associated with cardiometabolic risk, highlighting hypertension, and metabolic syndrome as the most frequently associated^22,28,34,39,45,46,54,56–58,61,63–66,68,71,82,83,86–89^. However, we reported a high number of associations that were null^33,41,45,48,65,66,68,69,71,72,85^, most of which were of risk factors with childhood hypertension, and only 5 were negative associations^44,52,63,64^. Alike the preconception period, hypertension ^22,28,34,45–48,54,56,57,61,63–66,68^ and metabolic syndrome ^22,28,34,45–48,54,56–58,61,63–66,68,71,72,82,83^ were the two outcomes most frequently associated with birth anthropometrics (Figure 2B). We identified 7 birth anthropometric risk factors correlated with cardiometabolic outcomes. Among them, the birth weight of the child had the most frequently associated with all of outcomes (n=25) ^28,33,39,44,45,48,52,54,58,61,63–66,68,69,71,72,82,83,87^, 13 of which were linked to an increased risk for childhood cardiometabolic issues^28,39,45,48,54,58,61,63,82,83,87^, while 9 had no relationship^33,45,65,66,68,69,71,72^, and 3 had inverse associations ^44,52,64^ (**Figure S3A**). The most frequent cardiometabolic outcome associated with birth weight was hypertension. Furthermore, low birth weight or small size for gestational age (LBW/SGA) was identified as a risk factor for glucose intolerance, hypertension, metabolic syndrome and T2D: 10 positive associations^15,34,54,58,61,64,65,68,86,88,89^ and 6 no associations were observed^45,48,66,72,85^ (**Figure S3B**). High birth weight (HBW) or large for gestational age (LGA), was evaluated by several studies as a potential risk factor for cardiometabolic outcomes. In fact, we identified 15 positive associations^22,34,58,63,66,72,87,89^ for HBW/LGA and cardiometabolic disorder risk and 4 no associations^48,68,69,85^ (**Figure S3C**). As part of the birth anthropometrics group, other exposures were identified, such as gestational age^39,45,56,58,63,71,82,83^, preterm birth^41,45,47,56,57,71,87^, birth length^44,61,63,82,83^, and mode of delivery (cesarean delivery)^46,58,87^. Despite the small amount of data, most associations were positive, highlighting both hypertension and metabolic syndrome as the most adverse cardiometabolic outcomes.

Regarding the sociodemographics category, most associations reported a risk of developing all cardiometabolic outcomes^28,39,45–48,51,53,54,57,58,61,64,77,81,87–89^. Figure 2C shows that sociodemographic factors increased the risk of hypertension^28,45–48,54,57,61,64^ and metabolic syndrome^35,58,77^ during childhood. The strongest associations were found between ethnicity^28,45,47,48,51,54,57,58^, maternal education^39,45–47,54,58,61,64,77^, and income^45,61,64,77,87,88^. Ethnicity was mostly associated with hypertension^28,45,57,58^ (**Figure S3D**). Similarly, low maternal education was positively associated with the risk to develop hypertension, and metabolic syndrome^45–47,54,61,64^ (**Figure S3E**). We also reported that low parental income is mostly associated with higher risk to develop hypertension, metabolic syndrome and T2D^45,61,64^ **(Figure S3F)**. Cardiometabolic outcomes were also influenced by other parental factors such as employment^61,64,77^, socioeconomic status^47,77,89^, and marital status^47^. Pregnancy complications were associated with all cardiometabolic outcomes^22,23,25,27,29,30,32,33,39,42,46,47,49,57,62-^ 64,70,81,85-87, highlighting hypertension^22,29,32,46,47,49,57,62–64^ and glucose intolerance^22,23,25,27,29,30,32,33^ as the most reported associations (Figure 2D). Among them, we identified gestational diabetes, pregnancy hypertension, and preeclampsia as the main exposures associated with cardiometabolic outcomes in children. Almost all associations between these risk factors were positive. All 27 studies on gestational diabetes had a positive association with all outcomes^22,23,25,27,29,30,32,33,39,47,49,63,70,81,85–87^ (**Figure S3G**). Regarding pregnancy-related hypertension, we found associations for dyslipidemia^39^ (1 positive association), hypertension^46,63,64^ (3 positive associations), and metabolic syndrome^81^ (1 no association) (**Figure S3H**). We evaluated the associations of preeclampsia with dyslipidemia^42^ (1 for no association), hypertension^42,46,47,57,62,81,87^ (4 for positive associations and 1 for no association), metabolic syndrome^81^ (1 for no association), and T2D^42,87^ (1 for positive association and 1 for no association) (**Figure S3I**).

In addition to the above categories, we also evaluated cord blood biomarkers, lifestyle, physical pregnancy, and environmental exposures (Figure 2). For the cord blood biomarkers, we found 1 study reporting that telomere length was positively associated with hypertension^48^; plasma folate, serum vitamin B12, and plasma homocysteine were negatively associated with metabolic syndrome^80^; and umbilical insulin levels were positively associated with glucose intolerance and T2D^28^. No evidence was found between plasma folate concentrations and hypertension^51^ (Figure 2E). The lifestyle category (including exposures such as smoking, alcohol, drugs, and diet) presented 24 associations with cardiometabolic outcomes^36,37,39,47,51,53,54,58,64,67,77^. The most frequent associations were related to hypertension during childhood^36,37,47,51,53,54,64,67^ (Figure 2F). Concerning physical pregnancy (such as parental anthropometrics, parental BMI, gestational weight gain), we identified 15 associations^22,39,45,47,53,58,63,64,74,76,81^, 14 of which were related to an increased risk of cardiometabolic outcomes, especially hypertension^22,45,47,63,64^ and metabolic syndrome^22,58,74,76,81^ (Figure 2G). The results in the environmental category contrasted, being mostly associated with hypertension. In this category, there are 7 associations, 3 of which were related to an increased risk of hypertension ^61,64,67^, while the other 2 were not associated with T2D^69,88^. Finally, two negative associations were also found with hypertension^55,64^ (Figure 2H).

### Early infancy-related risk factors associated with childhood cardiometabolic outcomes

We identified 19 studies reporting associations between risk factors in early infancy and adverse cardiometabolic outcomes in childhood. 2 were case-control studies^58,70^, 7 were retrospective studies^35,43,61,64,84,87,88^, and 10 were prospective studies^24,40,44,48,53,59,73,78,79,86^. We found 24 associations between risk factors and cardiometabolic outcomes in children, of which 15 positive associations were reported^33,40,43,48,53,61,68,70,73,78,79,86–88^, 4 negative associations were observed^44,64,70,87^, and 5 no associations were found^24,35,51,59,84^ (**Figure S2C**). During the period between birth and 2 years of age, feeding patterns and infant anthropometrics were the main risk factors evaluated. Anthropometrics in infancy were associated with dyslipidemia^40,43^ in 2 out of 3 studies, with hypertension^40,43,48,53,61^ in 5 out of 8 studies and with metabolic syndrome^73,78,79^ in 3 out of 3 studies. Only one study investigated anthropometrics and glucose intolerance, and did not report a significant association^24^, for T2D outcomes, no studies were identified. We did not find any study reporting infant anthropometrics or cardiometabolic outcomes (Figure 2I). Feeding patterns, which means breastfeeding, were the only category correlated mostly with a decreased risk for cardiometabolic outcomes. Indeed, 16 associations were reported, 8 of which were associated with a lower risk for glucose intolerance^33,68^, hypertension^68^, metabolic syndrome^33,70,78^ and T2D^86–88^. Only one study investigated breastfeeding and dyslipidemia, but did not report any significant associations ^35^, while we found 3 associations reporting a lower risk for hypertension^64^, metabolic syndrome^70^ and T2D^87^ (Figure 2J).

### Risk of bias assessment

Case-control studies, observational studies (prospective and retrospective studies), and RCTs were evaluated for risk of bias. Of the total studies analyzed (n=68), only 15% had low risk of bias. Regarding case-control studies, we found that 38% of studies showed low bias risk (n=8). For observational studies, the rate was 10% of low risk of bias (n=58). The percentage of these were distributed between prospective studies, which showed 16% of low bias risk (n=38). However, for retrospective studies, all exhibited at least one item unclear and/or high risk of bias (n=20). For the randomized control trials, 50% showed low risk of bias (n=2) (**Figure S3-S13**). In case-control studies, we identified that unreliable exposure measurements, and inadequate confounding strategies were the main conditions associated with high risk of bias (**Figure S14A**). In observational studies, we identified several conditions associated with high risk of bias, such as loss to follow-up, unreliable exposure measurements, and inadequate confounding strategies (**Figure S14B**). In the evaluated RCTs, we did not find high risk of bias for any of the included studies.

### Assessment of risk factors for prevention and prediction

We evaluated the quality of the assessment of risk factors included in the present systematic review (Figure 3). Risk factors with high scores on reproducibility, accuracy, standardization, stability, and technical variation were for the preconception period: maternal prepregnancy weight, in pregnancy and birth: maternal age, maternal and paternal education, income, parity, anthropometrics, paternal BMI, and gestational weight gain continuous; and for the early infancy period: breastfeeding, and infant BMI change. Risk factors quality scores were generally moderate to strong, except for cord blood biomarkers, which were low. A change in the marker that is linked with a change in the endpoint in one or more target populations was proven for maternal prepregnancy weight, birth weight, LBW/SGA, HBW/LGA, and gestational diabetes. The strongest theoretically modifiable risk factors on an individual level were diet, and the quality of the majority of risk factors were scored low based on evidence from the available intervention studies. Finally, for prediction, the proven risk factors were maternal prepregnancy weight, gestational diabetes, pregnancy hypertension, maternal education, and parental BMI. Concentrations of plasma homocysteine, parity, and marital status had the lowest scores (Figure 3).

**Figure 3.**
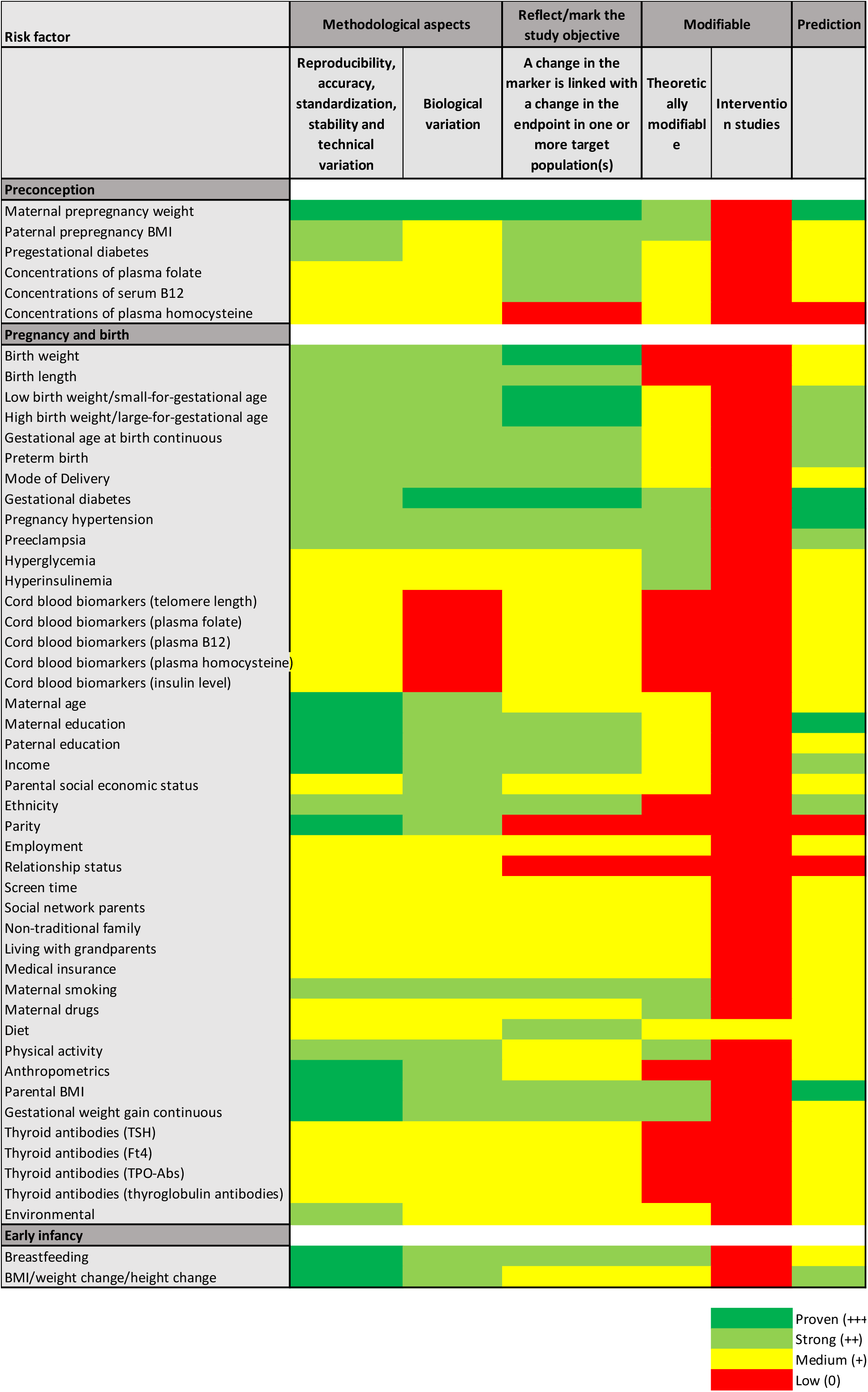
Quality assessment of identified risk factors based on ‘Guideline on quality assessment of risk factor’ from Calder et al. 2017. Four different levels has been represented: proven (hard green), strong (light green), moderate (yellow), and low (red).

## DISCUSSION

In the present systematic review, we examined the associations of early-life exposures during preconception, pregnancy and birth, and the early infancy period, with adverse cardiometabolic outcomes during childhood such as glucose intolerance, dyslipidemia, hypertension, metabolic syndrome, and T2D. We identified 59 observational studies (of which 38 prospectives, and 21 retrospectives), 7 case-control and 2 intervention studies, and we found 228 reported associations between risk factors and all cardiometabolic outcomes, of which 162 were positive. Exposures during pregnancy and at birth were the most frequently studied in relation to later life cardiometabolic risk factors. More specifically, high or low birth weight, sociodemographics factors including education, income and ethnicity, and pregnancy complications (including gestational diabetes and preeclampsia) were the most frequently reported risk factors associated with cardiometabolic risk. Additionally, we identified metabolic syndrome and hypertension as the most frequently and positive associated cardiometabolic outcomes reported.

This systematic review offers relevant information on the association between risk factors in the first 1,000 days and childhood cardiometabolic health. It encompasses 3 different timings of exposure: preconception, pregnancy and birth, and the early life period, as well risk factors such as parental and offspring conditions that might affect the cardiometabolic profile for up to 18 years. In preconception period, both paternal and maternal BMI were associated with adverse cardiometabolic health in childhood, including dyslipidemia, hypertension, metabolic syndrome and T2D. Maternal obesity before pregnancy was associated with impaired cardiometabolic health. Furthermore, paternal prepregnancy BMI was associated with adverse cardiometabolic profiles in offspring such as a higher childhood BMI, systolic blood pressure, insulin levels, and lower high-density lipoprotein cholesterol^75^, although strongest/most profound associations were observed for maternal prepregnancy BMI. In addition, during the preconception period, we found that pregestational diabetes was positively associated with T2D in children in one study. Thus, either pregestational or gestational diabetes in the mother has been associated with childhood-onset T2D^73,87^.

Pregnancy and birth were the most frequently studied period to investigate associations of early life exposures and later in life cardiometabolic outcomes. Our results add support to the developmental origins of health and disease (DOHaD) hypothesis, which states that adverse intrauterine exposure causes several metabolic changes during fetal growth, increasing the risk of cardiometabolic disease development ^90^. Birth anthropometrics, particularly having a high or low birth weight, showed a consistent positive association with all cardiometabolic outcomes, being hypertension the most frequently reported. A cross-sectional study suggests that obesity and hypertension in children and adolescents may be modifiable by birth weight^91^. Using Mendelian randomization, a study examined whether birth weight could be a risk factor independent of the intrauterine environment for cardiometabolic outcomes^92^. In the first 1,000 days of life, children born outside the normal birth weight range tend to regain their genetically determined growth trajectory to compensate for both restricted and excessive intrauterine growth^93^. In this way, the rapid weight gain in SGA infants increases the risk of obesity and metabolic diseases such as insulin resistance, impaired glucose tolerance, T2D, hypertension, and ultimately cardiovascular diseases^94,95^. It has also been demonstrated that birth weight has a direct impact on plasma triacylglycerols and insulin resistance, whereas obesity has a direct impact on hypertension, impaired glucose metabolism, and hypertriglyceridemia. Considering that birth weight and obesity cannot be separated from metabolic syndrome and its components, large prospective studies involving normal-weight individuals are needed. A recent meta-analysis showed that LBW might increase metabolic syndrome risk in childhood and adulthood^96^. To assess the potential interaction effects of LBW on metabolic syndrome, more high-quality studies are needed.

At least 30% of women exhibit pregnancy complications, which are among the leading causes of mortality among pregnant women^97^. These complications can be caused by the natural changes in the mother’s body as a response to her pregnancy, including increased cardiac output, an increased inflammatory response, and metabolic abnormalities related to insulin resistance^98^. As a consequence, pregnant women are more prone to develop metabolic disorders such as hypertension and gestational diabetes mellitus ^97,99^. Worldwide, pregnancy-related hypertension is the third leading cause of maternal mortality, affecting between 3% and 10% of pregnancies. A higher incidence of stillbirths and large for gestational-age babies is associated with gestational diabetes, which affects approximately 7–10% of pregnancies worldwide^100,101^. Both pregnancy complications are strongly associated with an increased risk of T2D and hypertension. In this systematic review, we found that gestational diabetes, which is one of the most common current diseases during pregnancy^102^, is associated with all cardiometabolic outcomes, with glucose intolerance having the greatest association. Hypertension during pregnancy is associated with dyslipidemia, hypertension and metabolic syndrome in children. We found associations between preeclampsia and dyslipidemia, hypertension, metabolic syndrome and T2D. Preeclampsia, like pregnancy hypertension, is associated with increased risks for hypertension, vascular disease, stroke, and venous thromboembolism later in life^103^. Therefore, gestational diabetes, pregnancy hypertension, and preeclampsia are risk factors for metabolic diseases, and the importance of pregnancy history to future health should be discussed with all care providers and women.

In the current research, we observed that several parental sociodemographic factors have been linked to adverse cardiometabolic outcomes. Among these factors, several risk factors, such as ethnicity, maternal education, and income, were associated with hypertension and other cardiometabolic outcomes. We found that ethnicity is mostly linked to hypertension, metabolic syndrome, dyslipidemia, and glucose intolerance. It is well known that ethnicity differs in cardiovascular disease prevalence^104^, with minorities having a more adverse cardiovascular profile than some European countries. Parental prepregnancy factors, pregnancy factors, and childhood BMI all contribute to these differences. In terms of income, we found a positive association with hypertension in children. The effect of household income has been analyzed for Black adolescents. This showed that households with incomes greater than $75,000 were more likely to have hypertensive-range blood pressure due to their black race compared to households with incomes less than $75,000 in United States of America, however, further research is needed ^105^. Both maternal and paternal education were associated with cardiometabolic outcomes, especially hypertension. In particular, evidence suggests that educational status might affect the development of adverse cardiometabolic health^106,107^. The majority of the studies show that a low level of maternal and paternal education is associated with a greater risk of hypertension during childhood^108,109^. Taken together, these data indicate that parental education level, rather than the individual’s education level, is independently better, showing that infants with low parental education levels are more likely to develop hypertension^110^.

Regarding the early infancy period, we found that breastfeeding is associated with a lower risk of cardiometabolic outcomes during childhood. Exclusive breastfeeding is highly recommended for infants for at least the first 6 months of life^111^, and breastfeeding has been shown to reduce the risk of obesity and chronic noncommunicable diseases, such as T2D^112^. The protein content of infant formulas is traditionally higher than that of human milk, and all essential amino acids are present in sufficient amounts. While these factors promote weight gain and growth, they also increase the risk of obesity and metabolic diseases^113^. For a formulation to be effective, its protein content needs to be as close as possible to that of human milk. Concerning the early infancy period, we also found that infant anthropometrics, especially weight gain and changes in BMI, are associated with hypertension and metabolic syndrome. These findings are relevant since previous studies have reported that rapid weight gain during infancy is associated with high blood pressure at 17 years^43^. Furthermore, rapid weight gain during infancy until 6 months could predict metabolic risk at 17 years^73^; therefore, maintaining or controlling early infancy weight gain might help to prevent future adult cardiovascular disease risks.

The main strengths of this systematic review has been the extensive search of the literature until 2022 (17610 articles in the first screening), and the implementation of at least two independent reviewers across the phases of screening and extraction, along with a third independent reviewer to resolve conflicts. The quality of the manuscript has been performed by evaluating the risk of bias assessment based on JBI guidelines. Using a standardized procedure, at least two independent reviewers conducted the review. All reviewers discussed to resolve the conflicts. As part of our attempt to minimize bias in the selection of studies, we adhered to predefined inclusion and exclusion criteria. By including a large body of evidence, we were able to provide a comprehensive and robust overview of the available evidence. We only identified 2 RCTs and all the associations have been based on observational research, either prospective or retrospective studies. Hence, causality cannot be established between the identified risk factors and cardiometabolic outcomes. However, there is a huge evidence about the potential risk factor to develop hypertension and metabolic syndrome later in childhood.

## Conclusions

We found that parental prepregnancy weight, birth weight, SGA, LGA, pregnancy complications such as hypertension and gestational diabetes, infant weight gain, and sociodemographics factors such as income, maternal education level and ethnicity, and not breastfeeding, were associated with cardiometabolic outcomes, highlighting hypertension and metabolic syndrome in children. These findings provide relevant information early-life risk prediction and modifying several behavioral patterns from conception to infancy, not only from mother to father but also from preconception to infancy. Finally, this systematic review provide useful information that could be applied in clinical practice to prevent or predict the metabolic complications later in childhood. Future studies focused on preconception factors, paternal factors, and sociodemographics are urgently needed to prevent metabolic disease.

## Article information

### Author contributions

MBH: provided extraction of the data and provided critical intellectual feedback and completed quality assessment of papers, and drafted the final version of manuscript. FJRO: Drafted the initial version of the manuscript, completed the review, extraction, and quality assessment of papers, and drafted the final version of manuscript. SMB: Drafted the initial version of the manuscript, completed the review, extraction, and quality assessment of papers. ASJK: Drafted the initial version of the manuscript, completed the review, extraction, and quality assessment of papers. EFV: provided extraction of the data and provided critical intellectual feedback. MAB: provided extraction of the data and provided critical intellectual feedback. MCC: provided extraction of the data and provided critical intellectual feedback. JvD: Set up the initial aim of the study and study protocol, provided extraction of the data and provided critical intellectual feedback. PI: provided extraction of the data and provided critical intellectual feedback. KK: provided extraction of the data and provided critical intellectual feedback. CAvLB: provided extraction of the data and provided critical intellectual feedback. RG: Set up the initial aim of the study and study protocol, completed the review, extraction, and quality assessment of papers. AG: Set up the initial aim of the study and study protocol, completed the review, extraction, and quality assessment of papers, and revised the final version of the manuscript. All authors approved the final version for publication.

## Data Availability

All data produced in the present work are contained in the manuscript

## Acknowledgements

RG received funding from the Netherlands Organization for Health Research and Development (ZonMW, grant number 543003109). RG has received funding from the European Union Horizon 2020 research and innovation program under the ERA-NET Cofund action (No. 727565), European Joint Programming Initiative A Healthy Diet for a Healthy Life (JPI HDHL), EndObesity, ZonMW Netherlands (No. 529051026). SMB was supported by the KNAW Ter Meulen Grant/KNAW Medical Science Fund, Royal Netherlands Academy of Arts & Sciences, and by ILSI Europe. ASJK was supported by ISLI Europe. MCC acknowledges the support by the Spanish Ministry of Science and Innovation (MCIN) research grant (ref. PID2022-139475OB-I00) and the award from MCIN/AEI to the Institute of Agrochemistry and Food Technology (IATA-CSIC) as Centre of Excellence Severo Ochoa (CEX2021-001189-S MCIN/AEI /10.13039/ 501100011033). EFV was supported by a Predoctoral grant awarded by the Spanish Ministry of Science and Innovation and the European Social Fund Plus (ESF+) for the training of doctors within the framework of the State Plan for Scientific, Technical and Innovation Research 2021 - 2023. (ref. CEX2021-001189-S-20-1). This paper will be part of Marco Brandimonte-Hernández’s doctorate, which is being completed as part of the “Nutrition and Food Sciences Program” at the University of Granada, Spain. FJRO was supported by the Spanish Government (Juan de la Cierva program, Ministry of Science and Innovation). This Project was executed in collaboration with the Early Nutrition and Long-term Health Task Force of the European Branch of the International Life Sciences Institute (ILSI Europe). Industry members of this task force are listed on the website www.ilsi.eu. For more information, please contact ILSI Europe by or call +32 2 771.00.14. The opinions expressed herein, and the conclusion of the project are those of the authors and do not necessarily represent the view of ILSI Europe nor those of its member companies.

## REFERENCES

1. Maximova K, Kuhle S, Davidson Z, Fung C, Veugelers PJ. Cardiovascular risk-factor profiles of normal and overweight children and adolescents: insights from the Canadian Health Measures Survey. Canadian Journal of Cardiology. 2013;29(8):976–982.

2. Jebeile H, Kelly AS, O’Malley G, Baur LA. Obesity in children and adolescents: epidemiology, causes, assessment, and management. Lancet Diabetes Endocrinol. 2022;10(5):351–365.

3. Saeedi P, Petersohn I, Salpea P, et al. Global and regional diabetes prevalence estimates for 2019 and projections for 2030 and 2045: Results from the International Diabetes Federation Diabetes Atlas. Diabetes research and clinical practice. 2019;157:107843.

4. WHO. Diabetes Atlas *10th ed Worl Helath Organization*, Geneva2021. 2021.

5. Maitre L, Julvez J, López-Vicente M, et al. Early-life environmental exposure determinants of child behavior in Europe: a longitudinal, population-based study. Environment international. 2021;153:106523.

6. Lurbe E, Cifkova R, Cruickshank JK, et al. Management of high blood pressure in children and adolescents: recommendations of the European Society of Hypertension. J Hypertens. 2009;27(9):1719–1742.

7. Lurbe E, Ingelfinger J. Developmental and Early Life Origins of Cardiometabolic Risk Factors: Novel Findings and Implications. Hypertension. 2021;77(2):308–318.

8. Finger JD, Busch MA, Du Y, et al. Time Trends in Cardiometabolic Risk Factors in Adults. Dtsch Arztebl Int. 2016;113(42):712–719.

9. Martínez-Jiménez MD, Gómez-García FJ, Gil-Campos M, Pérez-Navero JL. Comorbidities in childhood associated with extrauterine growth restriction in preterm infants: a scoping review. Eur J Pediatr. 2020;179(8):1255–1265.

10. Ordóñez-Díaz MD, Gil-Campos M, Flores-Rojas K, et al. Impaired Antioxidant Defence Status Is Associated With Metabolic-Inflammatory Risk Factors in Preterm Children With Extrauterine Growth Restriction: The BIORICA Cohort Study. Front Nutr. 2021;8:793862.

11. Kankowski L, Ardissino M, McCracken C, et al. The Impact of Maternal Obesity on Offspring Cardiovascular Health: A Systematic Literature Review. Front Endocrinol (Lausanne). 2022;13:868441.

12. Brown HL, Smith GN. Pregnancy Complications, Cardiovascular Risk Factors, and Future Heart Disease. Obstet Gynecol Clin North Am. 2020;47(3):487–495.

13. Pathirana MM, Lassi ZS, Roberts CT, Andraweera PH. Cardiovascular risk factors in offspring exposed to gestational diabetes mellitus in utero: systematic review and meta-analysis. J Dev Orig Health Dis. 2020;11(6):599–616.

14. Eberle C, Kirchner MF, Herden R, Stichling S. Paternal metabolic and cardiovascular programming of their offspring: A systematic scoping review. PLoS One. 2020;15(12):e0244826.

15. Santos Ferreira DL, Williams DM, Kangas AJ, et al. Association of pre-pregnancy body mass index with offspring metabolic profile: Analyses of 3 European prospective birth cohorts. PLoS medicine. 2017;14(8):e1002376.

16. McCarthy K, Ye Y-l, Yuan S, He Q-q. Peer Reviewed: Parental Weight Status and Offspring Cardiovascular Disease Risks: a Cross-Sectional Study of Chinese Children. Preventing Chronic Disease. 2015;12.

17. Labayen I, Ruiz JR, Ortega FB, et al. Intergenerational cardiovascular disease risk factors involve both maternal and paternal BMI. Diabetes care. 2010;33(4):894–900.

18. Black RE, Victora CG, Walker SP, et al. Maternal and child undernutrition and overweight in low-income and middle-income countries. The lancet. 2013;382(9890):427–451.

19. Larqué E, Labayen I, Flodmark C-E, et al. From conception to infancy—early risk factors for childhood obesity. Nature Reviews Endocrinology. 2019;15(8):456–478.

20. Barker TH, Stone JC, Sears K, et al. Revising the JBI quantitative critical appraisal tools to improve their applicability: an overview of methods and the development process. JBI Evidence Synthesis. 2023;21(3):478–493.

21. Calder PC, Boobis A, Braun D, et al. Improving selection of markers in nutrition research: evaluation of the criteria proposed by the ILSI Europe Marker Validation Initiative. Nutrition Research Reviews. 2017;30(1):73–81.

22. Boney CM, Verma A, Tucker R, Vohr BR. Metabolic syndrome in childhood: association with birth weight, maternal obesity, and gestational diabetes mellitus. Pediatrics. 2005;115(3):e290–e296.

23. Chandler-Laney PC, Bush NC, Granger WM, Rouse DJ, Mancuso MS, Gower BA. Overweight status and intrauterine exposure to gestational diabetes are associated with children’s metabolic health. Pediatric obesity. 2012;7(1):44–52.

24. Aris IM, Rifas-Shiman SL, Li L-J, et al. Association of weight for length vs body mass index during the first 2 years of life with cardiometabolic risk in early adolescence. JAMA network open. 2018;1(5):e182460–e182460.

25. Lowe WL Jr, Scholtens DM, Kuang A, et al. Hyperglycemia and adverse pregnancy outcome follow-up study (HAPO FUS): maternal gestational diabetes mellitus and childhood glucose metabolism. Diabetes care. 2019;42(3):372–380.

26. Scholtens DM, Kuang A, Lowe LP, et al. Hyperglycemia and Adverse Pregnancy Outcome Follow-up Study (HAPO FUS): maternal glycemia and childhood glucose metabolism. Diabetes care. 2019;42(3):381–392.

27. Silverman BL, Metzger BE, Cho NH, Loeb CA. Impaired glucose tolerance in adolescent offspring of diabetic mothers: relationship to fetal hyperinsulinism. Diabetes care. 1995;18(5):611–617.

28. Sun D, Wang T, Heianza Y, et al. Birthweight and cardiometabolic risk patterns in multiracial children. International journal of obesity. 2018;42(1):20–27.

29. Tam WH, Ma RCW, Ozaki R, et al. In utero exposure to maternal hyperglycemia increases childhood cardiometabolic risk in offspring. Diabetes care. 2017;40(5):679–686.

30. Tam WH, Ma RCW, Yang X, et al. Glucose intolerance and cardiometabolic risk in children exposed to maternal gestational diabetes mellitus in utero. Pediatrics. 2008;122(6):1229–1234.

31. Tam WH, Ma RCW, Yang X, et al. Glucose intolerance and cardiometabolic risk in adolescents exposed to maternal gestational diabetes: a 15-year follow-up study. Diabetes care. 2010;33(6):1382–1384.

32. Vääräsmäki M, Pouta A, Elliot P, et al. Adolescent manifestations of metabolic syndrome among children born to women with gestational diabetes in a general-population birth cohort. American journal of epidemiology. 2009;169(10):1209–1215.

33. Vandyousefi S, Goran MI, Gunderson EP, et al. Association of breastfeeding and gestational diabetes mellitus with the prevalence of prediabetes and the metabolic syndrome in offspring of Hispanic mothers. Pediatric obesity. 2019;14(7):e12515.

34. Wu Y, Yu X, Li Y, et al. Adipose tissue mediates associations of birth weight with glucose metabolism disorders in children. Obesity. 2019;27(5):746-755.

35. Izadi V, Kelishadi R, Qorbani M, et al. Duration of breast-feeding and cardiovascular risk factors among Iranian children and adolescents: the CASPIAN III study. Nutrition. 2013;29(5):744–751.

36. Mispireta M, Caulfield L, Zavaleta N, et al. Effect of maternal zinc supplementation on the cardiometabolic profile of Peruvian children: results from a randomized clinical trial. Journal of developmental origins of health and disease. 2017;8(1):56–64.

37. Stewart CP, Christian P, Schulze KJ, LeClerq SC, West Jr KP, Khatry SK. Antenatal micronutrient supplementation reduces metabolic syndrome in 6-to 8-year-old children in rural Nepal. The Journal of nutrition. 2009;139(8):1575–1581.

38. Pettitt DJ, Bennett PH, Saad MF, Charles MA, Nelson RG, Knowler WC. Abnormal glucose tolerance during pregnancy in Pima Indian women: long-term effects on offspring. Diabetes. 1991;40(Supplement_2):126–130.

39. Yang X, Leng J, Liu H, et al. Maternal gestational diabetes and childhood hyperlipidemia. Diabetic Medicine. 2021;38(11):e14606.

40. Berentzen NE, van Rossem L, Gehring U, et al. Overweight patterns throughout childhood and cardiometabolic markers in early adolescence. International Journal of Obesity. 2016;40(1):58–64.

41. Alves PJS, Araujo Junior E, Henriques ACPT, Carvalho FHC. Preterm at birth is not associated with greater cardiovascular risk in adolescence. The Journal of Maternal-Fetal & Neonatal Medicine. 2016;29(20):3351–3357.

42. Davidesko S, Nahum Sacks K, Friger M, Haim A, Sheiner E. Prenatal exposure to preeclampsia as a risk factor for long-term endocrine morbidity of the offspring. Hypertension in Pregnancy. 2021;40(1):21–28.

43. Fujita Y, Kouda K, Nakamura H, Iki M. Association of rapid weight gain during early childhood with cardiovascular risk factors in Japanese adolescents. Journal of Epidemiology. 2013;23(2):103–108.

44. Adair LS, Cole TJ. Rapid child growth raises blood pressure in adolescent boys who were thin at birth. Hypertension. 2003;41(3):451–456.

45. Barros FC, Victora CG. Increased blood pressure in adolescents who were small for gestational age at birth: a cohort study in Brazil. International Journal of Epidemiology. 1999;28(4):676–681.

46. Brunton NM, Dufault B, Dart A, Azad MB, McGavock JM. Maternal body mass index, offspring body mass index, and blood pressure at 18 years: a causal mediation analysis. International Journal of Obesity. 2021;45(12):2532–2538.

47. Chen Y, Zhao D, Wang B, Zhu J, Zhang J, Zhang Y. Association of intrauterine exposure to aspirin and blood pressure at 7 years of age: a secondary analysis. BJOG: An International Journal of Obstetrics & Gynaecology. 2019;126(5):599–607.

48. Hemachandra AH, Howards PP, Furth SL, Klebanoff MA. Birth weight, postnatal growth, and risk for high blood pressure at 7 years of age: results from the Collaborative Perinatal Project. Pediatrics. 2007;119(6):e1264–e1270.

49. Lu J, Zhang S, Li W, et al. Maternal gestational diabetes is associated with offspring’s hypertension. American journal of hypertension. 2019;32(4):335–342.

50. Martens DS, Sleurs H, Dockx Y, Rasking L, Plusquin M, Nawrot TS. Association of newborn telomere length with blood pressure in childhood. JAMA Network Open. 2022;5(8):e2225521–e2225521.

51. Ni Y, Szpiro A, Loftus C, et al. Associations Between Maternal Nutrition in Pregnancy and Child Blood Pressure at 4 –6 Years: A Prospective Study in a Community-Based Pregnancy Cohort. The Journal of nutrition. 2021;151(4):949–961.

52. Pereira JA, Rondó PH, Lemos JO, de Souza JMP, Dias RSC. The influence of birthweight on arterial blood pressure of children. Clinical nutrition. 2010;29(3):337–340.

53. Shankaran S, Bann CM, Bauer CR, et al. Prenatal cocaine exposure and BMI and blood pressure at 9 years of age. Journal of hypertension. 2010;28(6):1166–1175.

54. Shankaran S, Das A, Bauer CR, et al. Fetal origin of childhood disease: intrauterine growth restriction in term infants and risk for hypertension at 6 years of age. Archives of pediatrics & adolescent medicine. 2006;160(9):977–981.

55. Valvi D, Casas M, Romaguera D, et al. Prenatal phthalate exposure and childhood growth and blood pressure: evidence from the Spanish INMA-Sabadell Birth Cohort Study. Environmental health perspectives. 2015;123(10):1022–1029.

56. Vashishta N, Surapaneni V, Chawla S, Kapur G, Natarajan G. Association among prematurity (< 30 weeks’ gestational age), blood pressure, urinary albumin, calcium, and phosphate in early childhood. Pediatric Nephrology. 2017;32:1243–1250.

57. Vohr BR, Allan W, Katz KH, Schneider KC, Ment LR. Early predictors of hypertension in prematurely born adolescents. Acta Paediatrica. 2010;99(12):1812–1818.

58. Zhou Y-B, Li H-T, Si K-Y, Zhang Y-L, Wang L-L, Liu J-M. Association of elective cesarean delivery with metabolic measures in childhood: A prospective cohort study in China. *Nutrition*, Metabolism and Cardiovascular Diseases. 2019;29(8):775–782.

59. Aris IM, Chen L-W, Tint MT, et al. Body mass index trajectories in the first two years and subsequent childhood cardio-metabolic outcomes: a prospective multi-ethnic Asian cohort study. Scientific reports. 2017;7(1):8424.

60. Boerstra BA, Soepnel LM, Nicolaou V, et al. The impact of maternal hyperglycaemia first detected in pregnancy on offspring blood pressure in Soweto, South Africa. Journal of Hypertension. 2022;40(5):969–977.

61. Bowers K, Liu G, Wang P, et al. Birth weight, postnatal weight change, and risk for high blood pressure among Chinese children. Pediatrics. 2011;127(5):e1272–e1279.

62. Hoodbhoy Z, Mohammed N, Rozi S, et al. Cardiovascular dysfunction in children exposed to preeclampsia during fetal life. Journal of the American Society of Echocardiography. 2021;34(6):653–661.

63. Kuciene R, Dulskiene V. Associations of maternal gestational hypertension with high blood pressure and overweight/obesity in their adolescent offspring: a retrospective cohort study. Scientific reports. 2022;12(1):3800.

64. Liang X, Xiao L, Luo Y, Xu J. Prevalence and risk factors of childhood hypertension from birth through childhood: a retrospective cohort study. Journal of human hypertension. 2020;34(2):151–164.

65. Longo-Mbenza B, Ngiyulu R, Bayekula M, et al. Low birth weight and risk of hypertension in African school children. European Journal of Cardiovascular Prevention & Rehabilitation. 1999;6(5):311–314.

66. Sousa MACAd, Guimarães ICB, Daltro C, Guimarães AC. Association between birth weight and cardiovascular risk factors in adolescents. Arquivos brasileiros de cardiologia. 2013;101:09–17.

67. Xu S-L, Liu A-P, Wu Q-Z, et al. Pet ownership in utero and in childhood decreases the effects of environmental tobacco smoke exposure on hypertension in children: A large population based cohort study. Science of the total environment. 2020;715:136859.

68. Zarrati M, Shidfar F, Razmpoosh E, et al. Does low birth weight predict hypertension and obesity in schoolchildren? Annals of Nutrition and Metabolism. 2013;63(1-2):69–76.

69. Dong Y, Zou Z, Yang Z, et al. Association between high birth weight and hypertension in children and adolescents: a cross-sectional study in China. Journal of Human Hypertension. 2017;31(11):737–743.

70. Folic N, Folic M, Markovic S, Andjelkovic M, Jankovic S. Risk factors for the development of metabolic syndrome in obese children and adolescents. 2015.

71. García H, Loureiro C, Poggi H, et al. Insulin resistance parameters in children born very preterm and adequate for gestational age. *Endocrinology*, Diabetes & Metabolism. 2022;5(3):e00329.

72. Hirschler V, Bugna J, Roque M, Gilligan T, Gonzalez C. Does low birth weight predict obesity/overweight and metabolic syndrome in elementary school children? Archives of medical research. 2008;39(8):796–802.

73. Ekelund U, Ong KK, Linné Y, et al. Association of weight gain in infancy and early childhood with metabolic risk in young adults. The Journal of Clinical Endocrinology & Metabolism. 2007;92(1):98–103.

74. Gaillard R, Steegers E, Franco O, Hofman A, Jaddoe V. Maternal weight gain in different periods of pregnancy and childhood cardio-metabolic outcomes. The Generation R Study. International journal of obesity. 2015;39(4):677–685.

75. Gaillard R, Steegers EA, Duijts L, et al. Childhood cardiometabolic outcomes of maternal obesity during pregnancy: the Generation R Study. Hypertension. 2014;63(4):683–691.

76. Heikkinen A-L, Päkkilä F, Hartikainen A-L, Vääräsmäki M, Männistö T, Suvanto E. Maternal thyroid antibodies associates with cardiometabolic risk factors in children at the age of 16. The Journal of Clinical Endocrinology & Metabolism. 2017;102(11):4184–4190.

77. Iguacel I, Michels N, Ahrens W, et al. Prospective associations between socioeconomically disadvantaged groups and metabolic syndrome risk in European children. Results from the IDEFICS study. International Journal of Cardiology. 2018;272:333–340.

78. Khuc K, Blanco E, Burrows R, et al. Adolescent metabolic syndrome risk is increased with higher infancy weight gain and decreased with longer breast feeding. International journal of pediatrics. 2012;2012.

79. Marinkovic T, Toemen L, Kruithof CJ, et al. Early infant growth velocity patterns and cardiovascular and metabolic outcomes in childhood. The Journal of pediatrics. 2017;186:57–63. e54.

80. Monasso GS, Santos S, Geurtsen ML, Heil SG, Felix JF, Jaddoe VW. Associations of early pregnancy and neonatal circulating folate, vitamin B-12, and homocysteine concentrations with cardiometabolic risk factors in children at 10 y of age. The Journal of Nutrition. 2021;151(6):1628–1636.

81. Leybovitz-Haleluya N, Wainstock T, Landau D, Sheiner E. Maternal gestational diabetes mellitus and the risk of subsequent pediatric cardiovascular diseases of the offspring: a population-based cohort study with up to 18 years of follow up. Acta diabetologica. 2018;55:1037–1042.

82. Mardones F, Arnaiz P, Pacheco P, et al. Associations of prenatal growth with metabolic syndrome, insulin resistance, and nutritional status in Chilean children. BioMed Research International. 2014;2014.

83. Mardones F, Villarroel L, Arnaiz P, et al. Prenatal growth and metabolic syndrome components among Chilean children. Journal of Developmental Origins of Health and Disease. 2012;3(4):237–244.

84. Yakubov R, Nadir E, Stein R, Klein-Kremer A. The duration of breastfeeding and its association with metabolic syndrome among obese children. The Scientific World Journal. 2015;2015.

85. Young TK, Martens PJ, Taback SP, et al. Type 2 diabetes mellitus in children: prenatal and early infancy risk factors among native Canadians. Archives of pediatrics & adolescent medicine. 2002;156(7):651–655.

86. Pettitt DJ, Knowler WC. Long-term effects of the intrauterine environment, birth weight, and breast-feeding in Pima Indians. Diabetes care. 1998;21:B138.

87. Halipchuk J, Temple B, Dart A, Martin D, Sellers EA. Prenatal, obstetric and perinatal factors associated with the development of childhood-onset type 2 diabetes. Canadian journal of diabetes. 2018;42(1):71–77.

88. Martens PJ, Shafer LA, Dean HJ, et al. Breastfeeding initiation associated with reduced incidence of diabetes in mothers and offspring. Obstetrics & Gynecology. 2016;128(5):1095–1104.

89. Wei J-N, Sung F-C, Li C-Y, et al. Low birth weight and high birth weight infants are both at an increased risk to have type 2 diabetes among schoolchildren in Taiwan. Diabetes care. 2003;26(2):343–348.

90. Barker DJ. The fetal and infant origins of adult disease. BMJ: British Medical Journal. 1990;301(6761):1111.

91. Nugent JT, Lu Y, Deng Y, Sharifi M, Greenberg JH. Effect measure modification by birth weight on the association between overweight or obesity and hypertension in children and adolescents. JAMA pediatrics. 2023;177(7):735–737.

92. Warrington NM, Beaumont RN, Horikoshi M, et al. Maternal and fetal genetic effects on birth weight and their relevance to cardio-metabolic risk factors. Nature genetics. 2019;51(5):804–814.

93. Ong KK, Ahmed ML, Emmett PM, Preece MA, Dunger DB. Association between postnatal catch-up growth and obesity in childhood: prospective cohort study. Bmj. 2000;320(7240):967–971.

94. Bizerea-Moga TO, Pitulice L, Pantea CL, Olah O, Marginean O, Moga TV. Extreme birth weight and metabolic syndrome in children. Nutrients. 2022;14(1):204.

95. Chiavaroli V, Derraik JG, Hofman PL, Cutfield WS. Born large for gestational age: bigger is not always better. The Journal of pediatrics. 2016;170:307–311.

96. Liao L, Deng Y, Zhao D. Association of low birth weight and premature birth with the risk of metabolic syndrome: a meta-analysis. Frontiers in Pediatrics. 2020;8:405.

97. Fraser A, Nelson SM, Macdonald-Wallis C, et al. Associations of pregnancy complications with calculated cardiovascular disease risk and cardiovascular risk factors in middle age: the Avon Longitudinal Study of Parents and Children. Circulation. 2012;125(11):1367–1380.

98. Seely EW, Solomon CG. Insulin resistance and its potential role in pregnancy-induced hypertension. The journal of clinical endocrinology & metabolism. 2003;88(6):2393–2398.

99. Umesawa M, Kobashi G. Epidemiology of hypertensive disorders in pregnancy: prevalence, risk factors, predictors and prognosis. Hypertension Research. 2017;40(3):213–220.

100. Behboudi-Gandevani S, Amiri M, Bidhendi Yarandi R, Ramezani Tehrani F. The impact of diagnostic criteria for gestational diabetes on its prevalence: a systematic review and meta-analysis. Diabetology & metabolic syndrome. 2019;11:1–18.

101. Wambua S, Singh M, Okoth K, et al. Association between pregnancy-related complications and development of type 2 diabetes and hypertension in women: an umbrella review. BMC medicine. 2024;22(1):66.

102. Saravanan P, Magee LA, Banerjee A, et al. Gestational diabetes: opportunities for improving maternal and child health. The Lancet Diabetes & Endocrinology. 2020;8(9):793–800.

103. Leslie MS, Briggs LA. Preeclampsia and the Risk of Future Vascular Disease and Mortality: A Review. Journal of Midwifery & Women’s Health. 2016;61(3):315–324.

104. Bhopal RS, Humphry RW, Fischbacher CM. Changes in cardiovascular risk factors in relation to increasing ethnic inequalities in cardiovascular mortality: comparison of cross-sectional data in the Health Surveys for England 1999 and 2004. BMJ open. 2013;3(9):e003485.

105. Nagata JM, Shim JE, Balasubramanian P, et al. Sociodemographic Associations With Blood Pressure in 10–14-Year-Old Adolescents. Journal of Adolescent Health. 2024.

106. Cundiff JM, Lin SS-H, Faulk RD, McDonough IM. Educational quality may be a closer correlate of cardiometabolic health than educational attainment. Scientific Reports. 2022;12(1):18105.

107. Yu S, Sun Y. Low educational status correlates with a high incidence of mortality among hypertensive subjects from Northeast Rural China. Frontiers in Public Health. 2022;10:951930.

108. Suh SH, Song SH, Choi HS, et al. Parental educational status independently predicts the risk of prevalent hypertension in young adults. Scientific Reports. 2021;11(1):3698.

109. Amiri P, Rezaei M, Jalali-Farahani S, et al. Risk of hypertension in school-aged children with different parental risk: a longitudinal study from childhood to young adulthood. BMC pediatrics. 2021;21:1–11.

110. Kwok MK, Schooling CM, Subramanian SV, Leung GM, Kawachi I. Pathways from parental educational attainment to adolescent blood pressure. Journal of hypertension. 2016;34(9):1787–1795.

111. Butte NF, Lopez-Alarcon MG, Garza C. Nutrient adequacy of exclusive breastfeeding for the term infant during the first six months of life. World Health Organization; 2002.

112. Kelishadi R, Farajian S. The protective effects of breastfeeding on chronic non-communicable diseases in adulthood: A review of evidence. Advanced biomedical research. 2014;3(1):3.

113. Koletzko B, Bergmann K, Brenna JT, et al. Should formula for infants provide arachidonic acid along with DHA? A position paper of the European Academy of Paediatrics and the Child Health Foundation. The American journal of clinical nutrition. 2020;111(1):10–16.

