## Supplemental material for "A systematic review of associations between risk factors during the first 1000 days of life and cardiometabolic outcomes later in childhood"

**Text S1.** Search terms for the systematic review,
**Table S1.** Guideline on quality assessment of risk factors.
**Table S2.** Associations for each risk factor per period.
**Figure S1.** Flow chart showing selection of eligible studies
**Figure S2**. Number of positive, negative, and null associations for identified risk factors in A) preconception period, B) pregnancy and birth, and C) early infancy (<2 years).
**Figure S3**. Number of positive, negative, and null associations for birth weight (A), LBW/SGA (B), HBW/LGA (C), ethnicity (D), maternal education (E), income (F), gestational diabetes (G), pregnancy hypertension (H), and preeclampsia (I) exposures with each cardiometabolic outcome.
**Figure S4.** Risk of bias assessment of preconception risk factors in case-control studies.
**Figure S5.** Risk of bias assessment of preconception risk factors prospective studies.
**Figure S6.** Risk of bias assessment of preconception risk factors retrospective studies.
**Figure S7.** Risk of bias assessment of pregnancy and birth risk factors in case-control studies.
**Figure S8.** Risk of bias assessment of pregnancy and birth risk factors in randomized-controlled trials.
**Figure S9.** Risk of bias assessment of pregnancy and birth risk factors in prospective studies.
**Figure S10.** Risk of bias assessment of pregnancy and birth risk factors in retrospective studies.
**Figure S11.** Risk of bias assessment of early infancy risk factors in case-control studies.
**Figure S12.** Risk of bias assessment of early infancy risk factors in prospective studies.
**Figure S13.** Risk of bias assessment of early infancy risk factors in retrospective studies.
**Figure S14.** Assessment of bias in 5 domains.

**Text S1.** Search terms for the systematic review,

**Embase**

**https://www.embase.com/#advancedSearch/default – 10659 records**

('pregnancy'/exp OR 'pregnancy' OR 'pregnant woman'/exp OR 'pregnant woman' OR 'mother'/exp OR 'mother' OR 'prenatal exposure'/exp OR 'prenatal exposure' OR 'pregnancy outcome'/exp OR 'pregnancy outcome' OR 'pregnancy disorder'/exp OR 'pregnancy disorder' OR 'pregnancy complication'/exp OR 'pregnancy complication' OR 'high-risk pregnancy'/exp OR ‘adolescent pregnancy’/exp OR ‘maternal smoking’/exp OR 'prenatal period'/exp OR 'prenatal period' OR 'prenatal growth'/exp OR 'prenatal growth' OR 'fetus disease'/exp OR 'fetus disease' OR 'newborn disease'/exp OR 'newborn disease' OR 'low birth weight'/exp OR 'low birth weight' OR 'umbilical cord blood'/exp OR 'umbilical cord blood' OR 'infant disease'/exp OR 'infant disease' OR 'adolescent pregnancy'/exp OR 'adolescent pregnancy' OR gravidit*:ab,ti,kw OR gestation*:ab,ti,kw OR pregnan*:ab,ti,kw OR mother*:ab,ti,kw OR prenatal*:ab,ti,kw OR preeclamp*:ab,ti,kw OR 'pre eclamp*':ab,ti,kw OR (((prematur* OR preterm* OR live*) NEAR/3 (labor OR birth OR labour OR delivery OR ruptur*)):ab,ti,kw) OR birthweight*:ab,ti,kw OR (((birth OR neonat* OR newborn OR infant*) NEAR/3 weight*):ab,ti,kw) OR (((infant* OR child*) NEAR/3 ('breast feed*' OR breastfeed* OR 'milk composit*' OR weaning)):ab,ti,kw) OR ((infant* NEAR/3 (biomarker* OR saliva OR fecal OR urine OR hair)):ab,ti,kw) OR postpartum:ab,ti,kw OR 'post partum':ab,ti,kw OR puerperium:ab,ti,kw OR (((fetal OR foetus OR fetus OR newborn OR neonat* OR infant* OR intrauterin* OR 'intra uterin*') NEAR/3 (diseas* OR abnormal* OR growth* OR infect*)):ab,ti,kw) OR ((gestational NEAR/3 (diabet* OR hypertens* OR weight*)):ab,ti,kw) OR ((placenta* NEAR/3 abruption):ab,ti,kw) OR ((('small for date' OR 'small for gestational age' OR 'large for date' OR 'large for gestational age') NEAR/3 infant*):ab,ti,kw) OR preconcept*:ab,ti,kw OR maternal*:ab,ti,kw OR paternal*:ab,ti,kw OR father*:ab,ti,kw OR parent*:ab,ti,kw OR 'cord blood*':ab,ti,kw OR 'fetal blood*':ab,ti,kw OR lifestyle*:ab,ti,kw OR sociodemographic*:ab,ti,kw) **AND** ('follow up'/exp OR 'follow up' OR 'longitudinal study'/exp OR 'longitudinal study' OR 'prospective study'/exp OR 'prospective study' OR 'retrospective study'/exp OR 'retrospective study' OR 'cohort analysis'/exp OR 'cohort analysis' OR 'prediction'/exp OR 'prediction' OR followup*:ab,ti,kw OR 'follow up*':ab,ti,kw OR longitudinal*:ab,ti,kw OR prospective*:ab,ti,kw OR retrospective*:ab,ti,kw OR cohort*:ab,ti,kw OR predict*:ab,ti,kw) **AND** ((((obes* OR hypertension* OR diabetes* OR diabetic* OR bmi OR 'body mass*' OR 'waist circum*' OR 'fat distribut*' OR dyslipidemia* OR lipidemia OR lipoedema OR 'glucos* intoleranc*' OR 'glucos* toleranc*' OR overweight* OR overnutrition*) NEAR/6 (child* OR progeny OR offspring*)):ab,ti,kw) OR (((metabolic* OR cardiometabolic* OR 'insulin resist*' OR dysmetabolic* OR metabolism*) NEAR/2 (syndrom* OR diseas* OR outcome* OR disorder*) NEAR/9 (child* OR progeny OR offspring*)):ab,ti,kw) OR (((increas* OR elevat* OR high*) NEAR/2 (bloodpress* OR 'blood press*') NEAR/9 (child* OR progeny OR offspring*)):ab,ti,kw)) **NOT** (('animal'/exp OR animal OR animal*:de OR 'nonhuman'/exp OR nonhuman) NOT ('human'/exp OR 'human')) AND [english]/lim NOT ([conference abstract]/lim OR [preprint]/lim) NOT ('case report'/exp OR 'case report' OR 'case report*':ti)

**Medline**

**https://ovidsp.dc1.ovid.com/ovid-b/ovidweb.cgi – 11364 results.**
(exp Pregnancy/ OR Pregnant Women/ OR exp Parents/ OR exp Pregnancy Complications/ OR Pregnancy Outcome/ OR exp Infant, Newborn, Diseases/ OR exp Infant, Low Birth Weight/ OR Fetal Blood/ OR exp Pregnancy, High-Risk/ OR exp Pregnancy in Adolescence/ OR exp Biomarkers/ OR exp Cardiometablic Risk Factors/ OR exp sociodemographic factors/ OR exp maternal exposure/ OR exp Feeding Behavior/ OR exp Infant, Newborn, Disease/ OR (gravidit* OR gestation* OR pregnan* OR prenatal* OR maternal* OR preeclamp* OR pre-eclamp* OR ((prematur* OR preterm* OR live*) ADJ3 (labor OR birth OR labour OR delivery OR ruptur*)) OR birthweight* OR ((birth OR neonat* OR newborn OR infant*) ADJ3 (weight*)) OR ((infant* OR child*) ADJ3 (breast-feed* OR breastfeed* OR milk-composit* OR weaning)) OR ((infant*) ADJ3 (biomarker* OR saliva OR fecal OR urine OR hair)) OR postpartum OR post-partum OR puerperium OR ((fetal OR foetus OR fetus OR newborn OR neonat* OR infant* OR intrauterin* OR intra-uterin*) ADJ3 (diseas* OR abnormal* OR growth* OR infect*)) OR ((gestational) ADJ3 (diabet* OR hypertens* OR weight*)) OR ((placenta*) ADJ3 (abruption)) OR ((small-for-date OR small-for-gestational-age OR large-for-date OR large-for-gestational-age) ADJ3 (infant*)) OR preconcept* OR maternal* OR paternal* OR father* OR parent* OR cord-blood* OR fetal-blood* OR lifestyle* OR sociodemographic*).ab,ti,kf.) **AND** (exp Cohort Studies/ OR (followup* OR follow-up* OR longitudinal* OR prospective* OR retrospective* OR cohort* OR predict*).ab,ti,kf.) **AND** (exp Pediatric Obesity/ OR (((obes* OR hypertension* OR diabetes* OR diabetic* OR BMI OR body-mass* OR waist-circum* OR fat-distribut* OR dyslipidemia* OR lipidemia OR lipoedema OR glucos*-intoleranc* OR glucos*-toleranc* OR overweight* OR overnutrition*) ADJ6 (child* OR progeny OR offspring*)) OR ((metabolic* OR cardiometabolic* OR insulin-resist* OR dysmetabolic* OR metabolism*) ADJ2 (syndrom* OR diseas* OR outcome* OR disorder*) ADJ9 (child* OR progeny OR offspring*)) OR ((increas* OR elevat* OR high*) ADJ2 (bloodpress* OR blood-press*) ADJ9 (child* OR progeny OR offspring*))).ab,ti,kf.) NOT (exp animals/ NOT humans/) AND (english).lg NOT (news OR congres* OR abstract* OR book* OR chapter* OR dissertation abstract*).pt. NOT (Case Reports/ OR case-report*.ti.)

**Web of Science**
**https://www.webofscience.com/wos/woscc/advanced-search - 10034 records**
TS=(((gravidit* OR gestation* OR pregnan* OR mother* OR prenatal* OR preeclamp* OR pre-eclamp* OR ((prematur* OR preterm* OR live*) NEAR/2 (labor OR birth OR labour OR delivery OR ruptur*)) OR birthweight* OR ((birth OR neonat* OR newborn OR infant*) NEAR/2 (weight*)) OR ((infant* OR child*) NEAR/2 (breast-feed* OR breastfeed* OR milk-composit* OR weaning)) OR ((infant*) NEAR/2 (biomarker* OR saliva OR fecal OR urine OR hair)) OR postpartum OR post-partum OR puerperium OR ((fetal OR foetus OR fetus OR newborn OR neonat* OR infant* OR intrauterin* OR intra-uterin*) NEAR/2 (diseas* OR abnormal* OR growth* OR infect*)) OR ((gestational) NEAR/2 (diabet* OR hypertens* OR weight*)) OR ((placenta*) NEAR/2 (abruption)) OR ((small-for-date OR small-for-gestational-age OR large-for-date OR large-for-gestational-age) NEAR/2 (infant*)) OR preconcept* OR maternal* OR paternal* OR father* OR parent* OR cord-blood* OR fetal-blood* OR lifestyle* OR

sociodemographic*)) AND ((followup* OR follow-up* OR longitudinal* OR prospective* OR retrospective* OR cohort* OR predict*)) AND ((((obes* OR hypertension* OR diabetes* OR diabetic* OR BMI OR body-mass* OR waist-circum* OR fat-distribut* OR dyslipidemia* OR lipidemia OR lipoedema OR glucos*-intoleranc* OR glucos*-toleranc* OR overweight* OR overnutrition* ) NEAR/5 (child* OR progeny OR offspring*)) OR ((metabolic* OR cardiometabolic* OR insulin-resist* OR dysmetabolic* OR metabolism*) NEAR/2 (syndrom* OR diseas* OR outcome* OR disorder*) NEAR/9 (child* OR progeny OR offspring*)) OR ((increas* OR elevat* OR high*) NEAR/2 (bloodpress* OR blood-press*) NEAR/9 (child* OR progeny OR offspring*)))) NOT ((animal* OR rat OR rats OR mouse OR mice OR murine OR dog OR dogs OR canine OR cat OR cats OR feline OR rabbit OR cow OR cows OR bovine OR rodent* OR sheep OR ovine OR pig OR swine OR porcine OR veterinar* OR chick* OR zebrafish* OR baboon* OR nonhuman* OR primate* OR cattle* OR goose OR geese OR duck OR macaque* OR avian* OR bird* OR fish*) NOT (human* OR patient* OR women OR woman OR men OR man))) AND LA=(English) AND DT=(Article OR Review OR Letter OR Early Access)

**Scopus**

**Scopus - Advanced search – 1631 records**

( TITLE-ABS-KEY ( ( gravidit* OR gestation* OR pregnan* OR mother* OR prenatal* OR preeclamp* OR pre-eclamp* OR ( ( prematur* OR preterm* OR live* ) PRE/3 ( labor OR birth OR labour OR delivery OR ruptur* ) ) OR birthweight* OR ( ( birth OR neonat* OR newborn OR infant* ) PRE/3 ( weight ) ) OR ( ( infant* OR child* ) PRE/3 ( breast-feed* OR breastfeed* OR milk-composit* OR weaning ) ) OR ( ( infant* ) PRE/3 ( biomarker* OR saliva OR fecal OR urine OR hair ) ) OR postpartum OR post-partum OR puerperium OR ( ( fetal OR foetus OR fetus OR newborn OR neonat* OR infant* OR intrauterin* OR intra-uterin* ) PRE/3 ( diseas* OR abnormal* OR growth* OR infect* ) ) OR ( ( gestational ) PRE/3 ( diabet* OR hypertens* OR weight* ) ) OR ( ( placenta* ) PRE/3 ( abruption ) ) OR ( ( small-for-date OR small-for-gestational-age OR large-for-date OR large-for-gestational-age ) PRE/3 ( infant* ) ) OR preconcept* OR maternal* OR paternal* OR father* OR parent* OR cord-blood* OR fetal-blood* OR lifestyle* OR sociodemographic* ) ) ) AND ( TITLE-ABS-KEY ( "follow up" OR "longitudinal AND study" OR "prospective AND study" OR "retrospective AND study" OR "cohort AND analysis" OR "prediction" OR ( followup* OR follow-up* OR longitudinal* OR prospective* OR retrospective* OR cohort* OR predict* ) ) ) AND ( TITLE-ABS-KEY ( ( ( obes* OR hypertension* OR diabetes* OR diabetic* OR bmi OR body-mass* OR waist-circum* OR fat-distribut* OR dyslipidemia* OR lipidemia OR lipoedema OR glucos*-intoleranc* OR glucos*-toleranc* OR overweight* OR overnutrition*) PRE/6 ( child* OR progeny OR offspring* ) ) OR ( metabolic* OR cardiometabolic* OR insulin-resist* OR dymetabolic* OR metabolism* ) PRE/2 ( syndrom* OR diseas* OR outcome* OR disorder* ) PRE/9 ( child* OR progeny OR offspring* ) ) OR ( ( increas* OR elevat* OR high* ) PRE/2 ( bloodpress* OR blood-press* ) PRE/9 ( child* OR progeny OR offspring* ) ) ) AND NOT ( ( INDEXTERMS ( animals OR animal ) ) AND NOT ( INDEXTERMS ( humans OR human ) ) ) AND ( LIMIT-TO ( LANGUAGE,"English" ) ) AND ( EXCLUDE ( DOCTYPE,"cp" ) OR EXCLUDE ( DOCTYPE,"ch" ) OR EXCLUDE ( DOCTYPE,"le" ) )

**Cochrane**
**https://www.cochranelibrary.com/advanced-search/search-manager - 1896 records**
((gravidit* OR gestation* OR pregnan* OR mother* OR prenatal* OR preeclamp* OR pre NEXT/1 eclamp* OR ((prematur* OR preterm* OR live*) NEAR/3 (labor OR birth OR labour OR delivery OR ruptur*)) OR birthweight* OR ((birth OR neonat* OR newborn OR infant*) NEAR/3 (weight*)) OR ((infant* OR child*) NEAR/3 (breast NEXT/1 feed* OR breastfeed* OR milk NEXT/1 composit* OR weaning)) OR ((infant*) NEAR/3 (biomarker* OR saliva OR fecal OR urine OR hair)) OR postpartum OR post NEXT/1 partum OR puerperium OR ((fetal OR foetus OR fetus OR newborn OR neonat* OR infant* OR intrauterin* OR intra NEXT/1 uterin*) NEAR/3 (diseas* OR abnormal* OR growth* OR infect*)) OR ((gestational) NEAR/3 (diabet* OR hypertens* OR weight*)) OR ((placenta*) NEAR/3 (abruption)) OR ((small NEXT/1 for NEXT/1 date OR small NEXT/1 for NEXT/1 gestational NEXT/1 age OR large NEXT/1 for NEXT/1 date OR large NEXT/1 for NEXT/1 gestational NEXT/1 age) NEAR/3 (infant*)) OR preconcept* OR maternal* OR paternal* OR father* OR parent* OR cord NEXT/1 blood* OR fetal NEXT/1 blood* OR lifestyle* OR sociodemographic*):ab,ti,kw) **AND** ((followup* OR follow NEXT/1 up* OR longitudinal* OR prospective* OR retrospective* OR cohort* OR predict*):ab,ti,kw) **AND** ((((obes* OR hypertension* OR diabetes* OR diabetic* OR BMI OR body NEXT/1 mass* OR waist NEXT/1 circum* OR fat NEXT/1 distribut* OR dyslipidemia* OR lipidemia OR lipoedema OR glucos* NEXT/1 intoleranc* OR glucos* NEXT/1 toleranc* OR overweight* OR overnutrition*) NEAR/6 (child* OR progeny OR offspring*)) OR ((metabolic* OR cardiometabolic* OR insulin NEXT/1 resist* OR dysmetabolic* OR metabolism*) NEAR/2 (syndrom* OR diseas* OR outcome* OR disorder*) NEAR/9 (child* OR progeny OR offspring*)) OR ((increas* OR elevat* OR high*) NEAR/2 (bloodpress* OR blood NEXT/1 press*) NEAR/9 (child* OR progeny OR offspring*))):ab,ti,kw)

**Table S1.** Criteria template for quality assessment of the risk factors.

| | **Methodological aspects (excluding study design)** | | **Reflect/mark the study objective** | **Modifiable** | | **Prediction** |
|---|---|---|---|---|---|---|
| **Levels** | **Reproducibility, accuracy, standardization, stability (quality of the sample) and technical variation** | **Biological variation** | **A change in the marker is linked with a change in the endpoint in one or more target population(s)** | **Theoretically modifiable** | **Intervention studies** | |
| Proven (+++) | Marker is highly reproducible (intra class correlation coefficient >0.9) and accurate (less than 1% deviation from 'correct' value), assay is highly standardized, sample is stable or can easily be made stable | Minimal variation and relevant effects highly superior to variation: effects likely to be observed between groups of tens of individuals | Generally accepted marker (marker changes consistently linked with a change in the endpoint) | The risk factor is modifiable on an individual level and implementation of modifications in daily life is easy | A systematic review on intervention on this risk factor has been conducted and shows a positive effect of modification on the outcome | >80% of high quality papers report an association in the same direction and at least 5 publications |
| Strong (++) | Marker is reproducible and accurate enough to detect biological meaningful changes, assay is standardized, sample is stable or can be made stable | High variation explainable (for example, circadian cycle, age, sex, BMI, ethnicity and genotype) and possible to correct it and relevant effects reproducibly superior to variation: effects likely to be observed between groups of fifties to hundreds of individuals | Described as a cause- and effect relationship, but not (yet) generally accepted as a marker, due to a lack of (specific) studies | The risk factor is modifiable on an individual level and implementation of modifications in daily life is complicated | Multiple intervention studies have been conducted showing a potential positive effect of modification on the outcome | >65% of high quality papers report an association in the same direction and at least 5 publications |
| Medium (+) | Marker is reproducible and accurate enough for specific applications, assay is somewhat standardized and needs to be extensively processed or analyzed fast | High variation explainable (for example, circadian cycle, age, sex, BMI, ethnicity and genotype) and possible to correct it and relevant effects reproducibly close to variation: effects may be observed between groups of fifties to hundreds of individuals | Body of evidence suggesting correlation, but cause and effect not established | Modification of the risk factor is not possible on an individual and requires political, governmental, or medical interventions | There is some evidence of potential effect of intervention on the outcome, but literature is controversial | >50% of high quality report an association in the same direction or less than 5 publications |
| Low (0) | Reproducibility and accuracy of the marker, standardization of the assay and stability of the sample are either poor or not properly documented | High and unexplained variation in a short time span and relevant effects likely to be observed between groups of thousands of individuals | Plausible hypothesis, in use as an exploratory marker, but no substantial body of evidence yet. | The risk factor is not modifiable | No intervention studies have been conducted, or no potential effect of modification has been found | No consistent effect |

Adapted from Calder et al. 2017 (Nutrition Research Reviews).

**Table S2.** Associations for each risk factor per period.

| Author | Year | Country | Exposure (sub) | Cardiometabolic outcome | Association |
|---|---|---|---|---|---|
| **Preconception** | | | | | |
| **Physical** | | | | | |
| ***Maternal pre-pregnancy weight/BMI continuous*** | | | | | |
| *Barros | 1999 | Brazil | Maternal pre-pregnancy BMI | Hypertension | Positive |
| *Brunton | 2021 | UK | Maternal pre-pregnancy BMI | Hypertension | Positive |
| *Gaillard | 2014 | Netherlands | Maternal pre-pregnancy BMI | Metabolic syndrome | Positive |
| *Gaillard | 2015 | Netherlands | Maternal pre-pregnancy BMI | Metabolic syndrome | Positive |
| *Yang | 20216 | China | Pre-pregnancy BMI | Dyslipidemia | Positive |
| ***Paternal pre-pregnancy overweight/obesity*** | | | | | |
| *Gaillard | 2014 | Netherlands | Paternal pre-pregnancy BMI | Metabolic syndrome | Positive |
| ***Other*** | | | | | |
| *Halipchuk | 2018 | Canada | Pre-gestational diabetes | Type diabetes 2 | Positive |
| *Monasso | 2021 | Netherlands | Concentrations of plasma folate, serum vitamin B-12, plasma homocysteine | Metabolic syndrome | Negative |
| **Pregnancy and birth** | | | | | |
| **Birth anthropometrics** | | | | | |
| ***Birth weight continuous*** | | | | | |
| *Adair | 2003 | The Philippines | Birth weight | Hypertension | Negative |
| *Barros | 1999 | Brazil | Birth weight (≥ 2500 g) | Hypertension | None |
| *Barros | 1999 | Brazil | Birth weight/gestational age < 10th percentile | Hypertension | Positive |
| *Bowers | 2011 | China | Birth weight | Hypertension | Positive |
| *de Sousa | 2013 | Brazil | Birth weight | Hypertension | None |
| *Dong | 2017 | China | Birth weight | Hypertension | None |

| | | | | | |
|---|---|---|---|---|---|
| *Garcia | 2014 | Chile | Birth weight | Metabolic syndrome | None |
| *Halipchuk | 2018 | Canada | Birth weight | Type diabetes 2 | Positive |
| *Hemachandra | 2008 | USA | Birth weight | Hypertension | Positive |
| *Hirschler | 2008 | Argentina | Birth weight | Metabolic Syndrome | None |
| *Kuciene | 2022 | Lithuania | Birth weight | Hypertension | Positive |
| *Liang | 2020 | China | Birth weight | Hypertension | Negative |
| *Longo-Mbenza | 1999 | Democratic Republic of Congo | Birth weight | Hypertension | None |
| *Mardones | 2012 | Chile | Birth weight | Metabolic syndrome | Positive |
| *Mardones | 2014 | Chile | Birth weight | Metabolic syndrome | Positive |
| Pereira | 2010 | Brazil | Birth weight | Hypertension | Negative |
| *Shankaran | 2006 | USA | Birth weight | Hypertension | Positive |
| *Sun | 2018 | USA | Birth weight | Dyslipidemia; Glucose intolerance; Hypertension | Positive |
| *Vandyousefi | 2019 | USA | Birth weight | Glucose intolerance; Metabolic syndrome | None |
| *Yang | 2021 | China | Birth weight | Dyslipidemia | Positive |
| *Zarrati | 2013 | Iran | Birth weight | Hypertension | None |
| *Zhou | 2019 | China | Birth weight | Metabolic syndrome | Positive |
| ***Birth length continuous*** | | | | | |
| *Adair | 2003 | The Philippines | Birth lenght | Hypertension | Negative |
| *Bowers | 2011 | China | Birth lenght | Hypertension | Positive |

| | | | | | |
|---|---|---|---|---|---|
| *Kuciene | 2022 | Lithuania | Birth lenght | Hypertension | Positive |
| *Mardones | 2012 | Chile | Birth length | Metabolic syndrome | Positive |
| *Mardones | 2014 | Chile | Birth length | Metabolic syndrome | Positive |
| ***Low birth weight/small-for-gestational age*** | | | | | |
| *Barros | 1999 | Brazil | Low birth weight (< 2500 g) | Hypertension | None |
| *Bowers | 2011 | China | Low birth weight (< 2500 g) | Hypertension | Positive |
| *de Sousa | 2013 | Brazil | Low birth weight (< 2500 g) | Hypertension; Metabolic syndrome | None |
| *Hemachandra | 2008 | USA | Small for gestational age | Hypertension | None |
| *Hirschler | 2008 | Argentina | Low birth weight (≤ 2500 g) | Metabolic Syndrome | None |
| *Longo-Mbenza | 1999 | Democratic Republic of Congo | Low birth weight (< 2500 g) | Hypertension | Positive |
| *Liang | 2020 | China | Low birth weight (< 3000 g) | Hypertension | Positive |
| *Martens | 2016 | Canada | Small for gestational ag e | Type 2 diabetes | Positive |
| *Pettitt | 1998 | USA | Low birth weigth (≤ 2500 g) | Type 2 diabetes | Positive |
| *Shankaran | 2006 | USA | Intrauterine growth restriction | Hypertension | Positive |
| *Wei | 2003 | Taiwan | Low birth weight (< 2500 g) | Type 2 diabetes | Positive |
| *Wu | 2019 | China | Low birth weight (< 2500 g) | Glucose intolerance | Positive |
| *Young | 2002 | Canada | Low birth weight (< 2500 g) | Type 2 diabetes | None |
| *Zarrati | 2013 | Iran | Low birth weight (<2500 g) | Hypertension | Positive |
| *Zhou | 2019 | China | Low birth weight (<2500 g) | Metabolic syndrome | Positive |
| ***High birth weight/large-for-gestational age*** | | | | | |
| *Boney | 2005 | USA | Large for gestational age | Dyslipidemia; Glucose intolerance; | Positive |

| | | | | | |
|---|---|---|---|---|---|
| | | | | Hypertension; Metabolic syndrome | |
| *Bowers | 2011 | China | High birth weight (> 4000 g) | Hypertension | Positive |
| *de Sousa | 2013 | Brazil | High birth weight (> 4000 g) | Hypertension; Metabolic syndrome | Positive |
| *Dong | 2017 | China | High birth weight (> 4000 g) | Hypertension | None |
| *Halipchuk | 2018 | Canada | Large for gestational age | Type diabetes 2 | Positive |
| *Hemachandra | 2008 | USA | Large for gestational age | Hypertension | None |
| *Hirschler | 2008 | Argentina | High birth weight (≥ 4000g) | Metabolic Syndrome | Positive |
| *Kuciene | 2022 | Lithuania | High birth weight (> 4000 g) | Hypertension | Positive |
| *Kuciene | 2022 | Lithuania | Large for gestational age | Hypertension | Positive |
| *Pettitt | 1998 | USA | High birth weight (≥4500 g) | Type 2 diabetes | Positive |
| *Wei | 2003 | Taiwan | High birth weight (>4000 g) | Type 2 diabetes | Positive |
| *Wu | 2019 | China | High birth weight | Glucose intolerance | Positive |
| *Young | 2002 | Canada | High birth weight (>4000 g) | Type 2 diabetes | None |
| *Zarrati | 2013 | Iran | High birth weight (>4000 g) | Hypertension | None |
| *Zhou | 2019 | China | High birth weight (>4000 g) | Metabolic syndrome | Positive |
| ***Gestational age at birth continuous*** | | | | | |
| *Barros | 1999 | Brazil | Gestational age < 37 wks; > 37wks | Hypertension | None |
| *Garcia | 2014 | Chile | Gestational age at birth | Metabolic syndrome | Positive |
| *Kuciene | 2022 | Lithuania | Gestational age | Hypertension | Negative |
| *Mardones | 2012 | Chile | Gestational age (≤ 37 weeks) | Metabolic syndrome | Positive |
| *Mardones | 2014 | Chile | Gestational age (≤ 37 weeks) | Metabolic syndrome | Positive |
| *Vashishta | 2017 | USA | Gestational age | Hypertension | Positive |
| *Yang | 2021 | China | Gestational age at birth | Dyslipidemia | Positive |

| | | | | | |
|---|---|---|---|---|---|
| *Zhou | 2019 | China | Gestational age | Metabolic syndrome | Positive |
| ***Preterm birth*** | | | | | |
| *Alves | 2016 | Brazil | Preterm birth | Hypertension; Dyslipidemia | None |
| *Barros | 1999 | Brazil | Preterm | Hypertension | None |
| *Chen | 2019 | USA | Preterm | Hypertension | Positive |
| *Garcia | 2014 | Chile | Very preterm birth (< 32 weeks) | Metabolic syndrome | Positive |
| *Halipchuk | 2018 | Canada | Preterm | Type diabetes 2 | Positive |
| *Vashishta | 2017 | USA | Preterm (<30 weeks) | Hypertension | Positive |
| *Vohr | 2010 | USA | Preterm (> 600 g <1250g) | Hypertension | Positive |
| ***Mode of Delivery (CS)*** | | | | | |
| *Brunton | 2021 | UK | Mode of delivery (CS) | Hypertension | Positive |
| *Halipchuk | 2018 | Canada | Mode of delivery (CS) | Type diabetes 2 | Positive |
| *Zhou | 2019 | China | Mode of delivery (CS) | Metabolic syndrome | Positive |
| **Pregnancy complications** | | | | | |
| ***Gestational diabetes*** | | | | | |
| *Boney | 2005 | USA | Gestational diabetes | Dyslipidemia; Glucose intolerance; Hypertension; Metabolic syndrome | Positive |
| Chandler-Laney | 2012 | USA | Gestational diabetes | Glucose intolerance | Positive |
| *Chen | 2019 | USA | Gestational diabetes | Hypertension | Positive |
| *Folić | 2015 | Serbia | Gestational diabetes | Metabolic syndrome | Positive |
| *Halipchuk | 2018 | Canada | Gestational diabetes | Type diabetes 2 | Positive |
| Järvelin | 2009 | Finland | Gestational diabetes | Dyslipidemia; Glucose | Positive |

| | | | | | |
|---|---|---|---|---|---|
| | | | | intolerance; Hypertension; Metabolic Syndrome | |
| *Kuciene | 2022 | Lithuania | Gestational diabetes | Hypertension | Positive |
| *Leybovitz-Haleluya | 2018 | Israel | Gestational diabetes | Metabolic syndrome | Positive |
| Lowe | 2019 | USA | Gestational diabetes | Glucose intolerance | Positive |
| Lu | 2019 | China | Gestational diabetes | Hypertension | Positive |
| *Pettitt | 1998 | USA | Gestational diabetes | Type 2 diabetes | Positive |
| Silverman | 1995 | USA | Gestational diabetes | Glucose intolerance; Type 2 diabetes | Positive |
| *Tam | 2008 | China | Gestational diabetes | Glucose intolerance; Type 2 diabetes | Positive |
| *Tam | 2017 | China | Gestational diabetes | Glucose intolerance; Hypertension | Positive |
| *Vandyousefi | 2019 | USA | Gestational diabetes | Glucose intolerance; Metabolic syndrome | Positive |
| *Yang | 2021 | China | Gestational diabetes | Dyslipidemia | Positive |
| *Young | 2002 | Canada | Gestational diabetes | Type 2 diabetes | Positive |
| ***Pregnancy hypertension*** | | | | | |
| *Brunton | 2021 | UK | Pregnacy hypertension | Hypertension | Positive |
| *Kuciene | 2022 | Lithuania | Pregnancy hypertension | Hypertension | Positive |
| *Leybovitz-Haleluya | 2018 | Israel | Pregnancy hypertension | Metabolic syndrome | None |
| *Liang | 2020 | China | Pregnancy hypertension | Hypertension | Positive |
| *Yang | 2021 | China | Pregnancy hypertension | Dyslipidemia | Positive |

| | | | | | |
|---|---|---|---|---|---|
| ***Pre-eclampsia*** | | | | | |
| *Brunton | 2021 | UK | Pre-eclampsia | Hypertension | Positive |
| *Chen | 2019 | USA | Pre-eclampsia | Hypertension | Positive |
| Davidesko | 2021 | Israel | Pre-eclampsia | Dyslipidemia; Type 2 diabetes | None |
| *Halipchuk | 2018 | Canada | Pre-eclampsia | Type diabetes 2 | Positive |
| Hoodboy | 2021 | Pakistan | Pre-eclampsia | Hypertension | Positive |
| *Leybovitz-Haleluya | 2018 | Israel | Pre-eclampsia | Metabolic syndrome | None |
| *Vohr | 2010 | USA | Pre-eclampsia | Hypertension | Positive |
| ***Other*** | | | | | |
| Boerstra | 2022 | South Africa | Hyperglycemia | Hypertension | None |
| Pettit | 1991 | USA | Hyperglycemia | Glucose intolerance; Type 2 diabetes | Positive |
| *Pettitt | 1998 | USA | Hyperglycemia | Type 2 diabetes | Positive |
| Scholtens | 2019 | USA, Canada, Israel, UK, China | Hyperglycemia | Glucose intolerance | Positive |
| Tam | 2010 | China | Hyperinsulinemia | Metabolic syndrome | Positive |
| **Cord blood biomarkers** | | | | | |
| Martens | 2022 | Canada | Telemere length | Hypertension | Positive |
| Monasso | 2021 | Netherlands | Concentrations of plasma folate, serum vitamin B-12, plasma homocysteine | Metabolic syndrome | Negative |
| *Ni | 2021 | USA | Plasma folate concentrations | Hypertension | None |
| *Tam | 2008 | China | Umbilical insulin level | Glucose intolerance; Type 2 diabetes | Positive |
| **Sociodemographic** | | | | | |
| ***Maternal age*** | | | | | |
| *Chen | 2019 | USA | Maternal age | Hypertension | Positive |

| | | | | | |
|---|---|---|---|---|---|
| *Leybovitz-Haleluya | 2018 | Israel | Age | Metabolic syndrome | None |
| *Yang | 2021 | China | Age | Dyslipidemia | Positive |
| *Zhou | 2019 | China | Age | Metabolic syndrome | Positive |
| ***Maternal education*** | | | | | |
| *Barros | 1999 | Brazil | Education (Low) | Hypertension | Positive |
| *Bowers | 2011 | China | Education (High) | Hypertension | Positive |
| *Brunton | 2021 | UK | Education (Low) | Hypertension | Positive |
| *Chen | 2019 | USA | Education (Low) | Hypertension | Positive |
| *Iguacel | 2018 | Belgium, Cyprus, Estonia, Germany, Hungary, Italy, Spain and Sweden | Education (medium and low) | Metabolic syndrome | Positive |
| *Liang | 2020 | China | Education | Hypertension | Positive |
| *Shankaran | 2006 | USA | Education | Hypertension | Positive |
| *Yang | 2021 | China | Education | Dyslipidemia | None |
| *Zhou | 2019 | China | Education | Metabolic syndrome | Positive |
| ***Paternal education*** | | | | | |
| *Bowers | 2011 | China | Paternal education (High) | Hypertension | Positive |
| *Iguacel | 2018 | Belgium, Cyprus, Estonia, Germany, Hungary, Italy, Spain and Sweden | Education (medium-low) | Metabolic syndrome | Positive |
| *Liang | 2020 | China | Education | Hypertension | Positive |
| *Shankaran | 2006 | USA | Education | Hypertension | None |

| | | | | | |
|---|---|---|---|---|---|
| ***Income*** | | | | | |
| *Barros | 1999 | Brazil | Income (Low) | Hypertension | Positive |
| *Bowers | 2011 | China | Income (High) | Hypertension | Positive |
| *Halipchuk | 2018 | Canada | Income (Low) | Type diabetes 2 | Positive |
| *Iguacel | 2018 | Belgium, Cyprus, Estonia, Germany, Hungary, Italy, Spain and Sweden | Income (Low) | Metabolic syndrome | Positive |
| *Liang | 2020 | China | Income (low) | Hypertension | Positive |
| *Liang | 2020 | China | Income (High) | Hypertension | Negative |
| *Martens | 2016 | Canada | Income | Type 2 diabetes | None |
| ***Parental social economic status*** | | | | | |
| *Chen | 2019 | USA | Socio economic status (Low) | Hypertension | Positive |
| *Iguacel | 2018 | Belgium, Cyprus, Estonia, Germany, Hungary, Italy, Spain and Sweden | Socio economic status (low) | Metabolic syndrome | Positive |
| *Wei | 2003 | Taiwan | Socioeconomic status | Type 2 diabetes | None |
| ***Ethnicity*** | | | | | |
| *Barros | 1999 | Brazil | Ethnicity (Black race) | Hypertension | Positive |
| *Chen | 2019 | USA | Ethnicity (White race) | Hypertension | Negative |
| *Hemachandra | 2008 | USA | Ethnicity (White race) | Hypertension | Positive |
| *Ni | 2021 | USA | Ethnicity | Hypertension | None |
| *Shankaran | 2006 | USA | Ethnicity (Black Americans > White Americans) | Hypertension | None |
| *Sun | 2018 | USA | Ethnicity | Dyslipidemia; Glucose | Positive |

| | | | | | |
|---|---|---|---|---|---|
| | | | | intolerance; Hypertension | |
| *Vohr | 2010 | USA | Ethnicity | Hypertension | Positive |
| *Zhou | 2019 | China | Ethnicity | Metabolic syndrome | Positive |
| ***Parity*** | | | | | |
| *Martens | 2016 | Canada | Parity | Type 2 diabetes | None |

| | | | | | |
|---|---|---|---|---|---|
| ***Employment*** | | | | | |
| *Bowers | 2011 | China | Employment | Hypertension | Positive |
| *Iguacel | 2018 | Belgium, Cyprus, Estonia, Germany, Hungary, Italy, Spain and Sweden | Unemployed | Metabolic syndrome | |
| *Liang | 2020 | China | Employment | Hypertension | |
| ***Marital status*** | | | | | |
| *Chen | 2019 | USA | Marital status | Hypertension | Positive |
| ***Other*** | | | | | |
| *Iguacel | 2018 | Belgium, Cyprus, Estonia, Germany, Hungary, Italy, Spain and Sweden | Screen time | Metabolic syndrome | Positive |
| *Iguacel | 2018 | Belgium, Cyprus, Estonia, Germany, Hungary, Italy, Spain and Sweden | Social network parents | Metabolic syndrome | Positive |
| *Iguacel | 2018 | Belgium, Cyprus, Estonia, Germany, Hungary, Italy, | Non-traditional family | Metabolic syndrome | Positive |

| | | | | | |
|---|---|---|---|---|---|
| | | Spain and Sweden | | | |
| *Liang | 2020 | China | Living with grandparents | Hypertension | Positive |
| *Liang | 2020 | China | Medical insurance | Hypertension | Negative |
| **Lifestyle** | | | | | |
| ***Maternal smoking*** | | | | | |
| *Ni | 2021 | USA | Smoking | Hypertension | None |
| *Shankaran | 2006 | USA | Smoking | Hypertension | None |
| *Shankaran | 2010 | USA | Smoking | Hypertension | Positive |
| *Xu | 2020 | China | Smoking | Hypertension | Positive |
| *Yang | 2021 | China | Smoking | Dyslipidemia | Positive |
| ***Maternal alcohol*** | | | | | |
| *Shankaran | 2006 | USA | Alcohol | Hypertension | None |
| *Shankaran | 2010 | USA | Alcohol consumption | Hypertension | Positive |
| ***Maternal drugs*** | | | | | |
| *Chen | 2019 | USA | Drugs use: aspirin | Hypertension | Negative |
| *Shankaran | 2006 | USA | Drugs use: cocaine, marijuana, other | Hypertension | Positive |
| *Shankaran | 2010 | USA | Drugs use: cocaine, other | Hypertension | Positive |
| ***Diet*** | | | | | |
| *Iguacel | 2018 | Belgium, Cyprus, Estonia, Germany, Hungary, Italy, Spain and Sweden | Diet: lower fruit and vegetables intake | Metabolic syndrome | None |
| *Liang | 2020 | China | Diet: cereals and potatoes | Hypertension | Negative |
| *Liang | 2020 | China | Diet: Pickle intake | Hypertension | Positive |
| Mispireta | 2017 | Perú | Diet: maternal zinc supplementation | Glucose intolerance; Hypertension | None |
| *Ni | 2021 | USA | Diet | Hypertension | None |

| | | | | | |
|---|---|---|---|---|---|
| Stewart | 2009 | Nepal | Diet: micronutrient supplementation | Dyslipidemia; Glucose intolerance; Hypertension; Metabolic syndrome | Positive |
| *Zhou | 2019 | China | Diet | Metabolic syndrome | Positive |
| ***Physical activity*** | | | | | |
| *Iguacel | 2018 | Belgium, Cyprus, Estonia, Germany, Hungary, Italy, Spain and Sweden | Physical activity (Low) | Metabolic syndrome | None |
| *Liang | 2020 | China | Physical activity (Low) | Hypertension | Positive |
| *Liang | 2020 | China | Physical activity | Hypertension | Negative |
| **Physical** | | | | | |
| ***Anthropometrics*** | | | | | |
| *Shankaran | 2010 | USA | Maternal weight | Hypertension | Positive |
| ***Parental BMI or ovesrweight/obesity*** | | | | | |
| *Barros | 1999 | Brazil | Maternal BMI (High) | Hypertension | Positive |
| *Boney | 2005 | USA | Maternal obesity | Dyslipidemia; Glucose intolerance; Hypertension; Metabolic syndrome | Positive |
| *Chen | 2019 | USA | Maternal overweigth/Obesity | Hypertension | Positive |
| *Kuciene | 2022 | Lithuania | Maternal obesity | Hypertension | Positive |

| | | | | | |
|---|---|---|---|---|---|
| *Leybovitz-Haleluya | 2018 | Israel | Obesity | Metabolic syndrome | None |
| *Liang | 2020 | China | Maternal/paternal obesity | Hypertension | Positive |
| *Zhou | 2019 | China | Maternal BMI | Metabolic syndrome | Positive |
| ***Gestational weight gain continuous*** | | | | | |
| Gaillard | 2015 | Netherlands | Gestational weight gain | Metabolic syndrome | Positive |
| *Yang | 2021 | China | Gestational weight gain | Dyslipidemia | Positive |
| *Zhou | 2019 | China | Gestational weight gain | Metabolic syndrome | Positive |
| ***Other*** | | | | | |
| *Heikkinen | 2017 | Finland | Thyroid antibodies (TSH, fT4, TPO-Abs, thyroglobulin antibodies | Metabolic syndrome | Positive |
| **Environmental** | | | | | |
| *Bowers | 2011 | China | Rural area | Hypertension | Positive |
| *Dong | 2017 | China | Urban/rural area | Hypertension | None |
| *Liang | 2020 | China | Rural, urban or suburban area | Hypertension | Positive |
| *Liang | 2020 | China | City or suburb | Hypertension | Negative |
| *Martens | 2016 | Canada | Rural residence | Type 2 diabetes | None |
| *Valvi | 2016 | Spain | Endocrine-disrupting chemicals exposure (PFAS, phthalates, bisphenols, etc) | Hypertension | Negative |
| *Xu | 2020 | China | Pet ownership | Hypertension | Positive |
| **Early infancy** | | | | | |
| **Feeding patterns** | | | | | |
| ***Breastfeeding*** | | | | | |
| *Folić | 2015 | Serbia | Breastfeeding (during first six months of life) | Metabolic syndrome | Positive |

| | | | | | |
|---|---|---|---|---|---|
| *Folić | 2015 | Serbia | Lack of breastfeeding | Metabolic syndrome | Negative |
| *Halipchuk | 2018 | Canada | Breastfeeding initiation | Type diabetes 2 | Positive |
| *Halipchuk | 2018 | Canada | No breastfeeding | Type diabetes 2 | Negative |
| *Khuc | 2012 | Chile | Breastfeeding (≥ 90 days) | Metabolic syndrome | Positive |
| Izadi | 2013 | Iran | Breastfeeding | Dyslipidemia; Glucose intolerance; Hypertension | None |
| *Martens | 2016 | Canada | Breastfeeding | Type 2 diabetes | Positive |
| *Ni | 2021 | USA | Breastfeeding | Hypertension | None |
| *Pettitt | 1998 | USA | Breastfeeding | Type 2 diabetes | Positive |
| *Vandyousefi | 2019 | USA | Breastfeeding | Glucose intolerance; Metabolic syndrome | Positive |
| Yakubov | 2015 | Israel | Breastfeeding | Metabolic syndrome | None |
| *Zarrati | 2013 | Iran | Breastfeeding (> 12 months) | Hypertension | Positive |
| **Infant anthropometrics** | | | | | |
| ***BMI/weight change/height change*** | | | | | |
| *Adair | 2003 | The Philippines | Weight change | Hypertension | Negative |
| Aris | 2017 | Singapore | BMI change | Hypertension | None |
| Aris | 2018 | USA; Belarus | BMI change, Weight change | Dyslipidemia; Glucose intolerance; Hypertension | None |
| *Berentzen | 2016 | Netherlands | BMI change | Dyslipidemia; Hypertension | Positive |
| *Bowers | 2011 | China | Weight change | Hypertension | Positive |

| | | | | | |
|---|---|---|---|---|---|
| Ekelund | 2007 | Sweden | Weight change | Metabolic syndrome | Positive |
| *Fujita | 2013 | Japan | BMI change; Weight change | Dyslipidemia; Hypertension | Positive |
| *Hemachandra | 2008 | USA | Weight change | Hypertension | Positive |
| *Khuc | 2012 | Chile | Weight change | Metabolic syndrome | Positive |
| Marinkovic | 2017 | Netherlands | BMI change; Weight change | Metabolic syndrome | Positive |
| *Shankaran | 2010 | USA | Weight change | Hypertension | Positive |

*Study multiple times in table

**Figure S1.** Flow chart showing selection of eligible studies

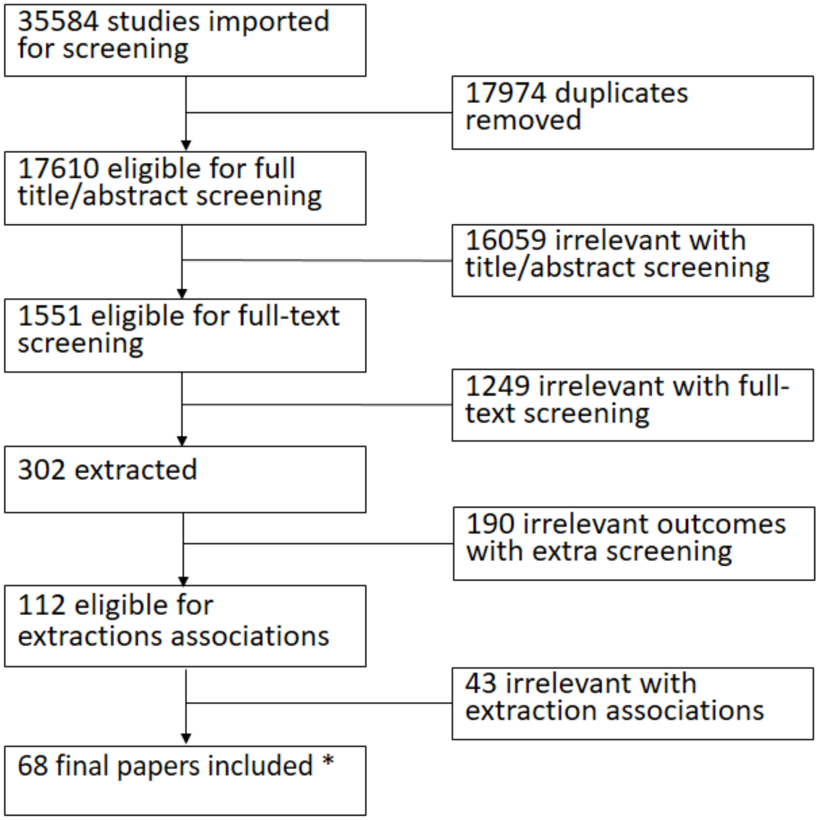

**Figure S2.** Number of positive, negative, and null associations for identified risk factors in A) preconception period, B) pregnancy and birth, and C) early infancy (<2 years).

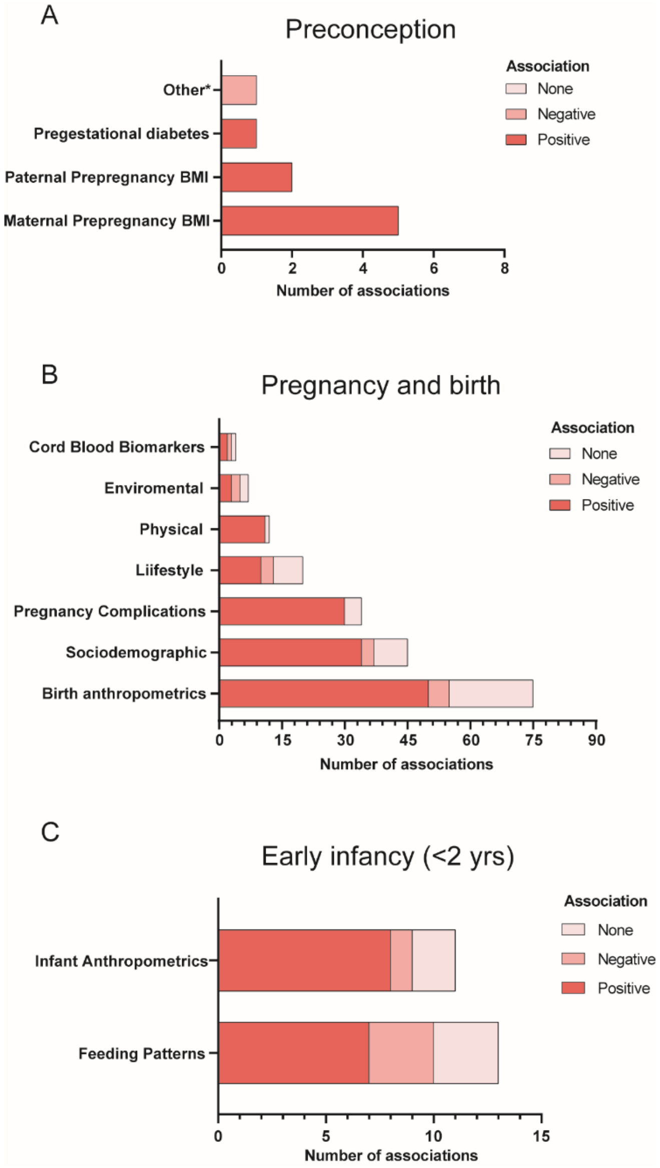

**Figure S3**. Number of positive, negative, and null associations for birth weight (A), LBW/SGA (B), HBW/LGA (C), ethnicity (D), maternal education (E), income (F), gestational diabetes (G), pregnancy hypertension (H), and preeclampsia (I) exposures with each cardiometabolic outcome. HBW: high-birth weight; LGA: large for gestational age; LBW: low-birth weight; SGA: small for gestational age.

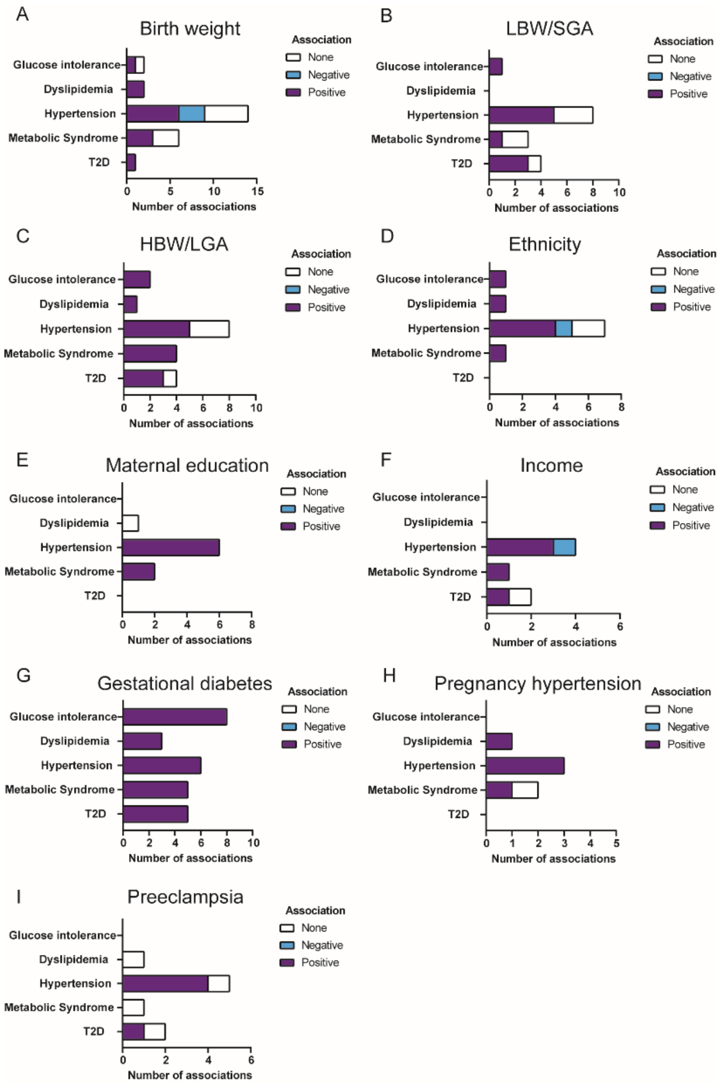

**Figure S4.** Risk of bias assessment of preconception risk factors in case-control studies.

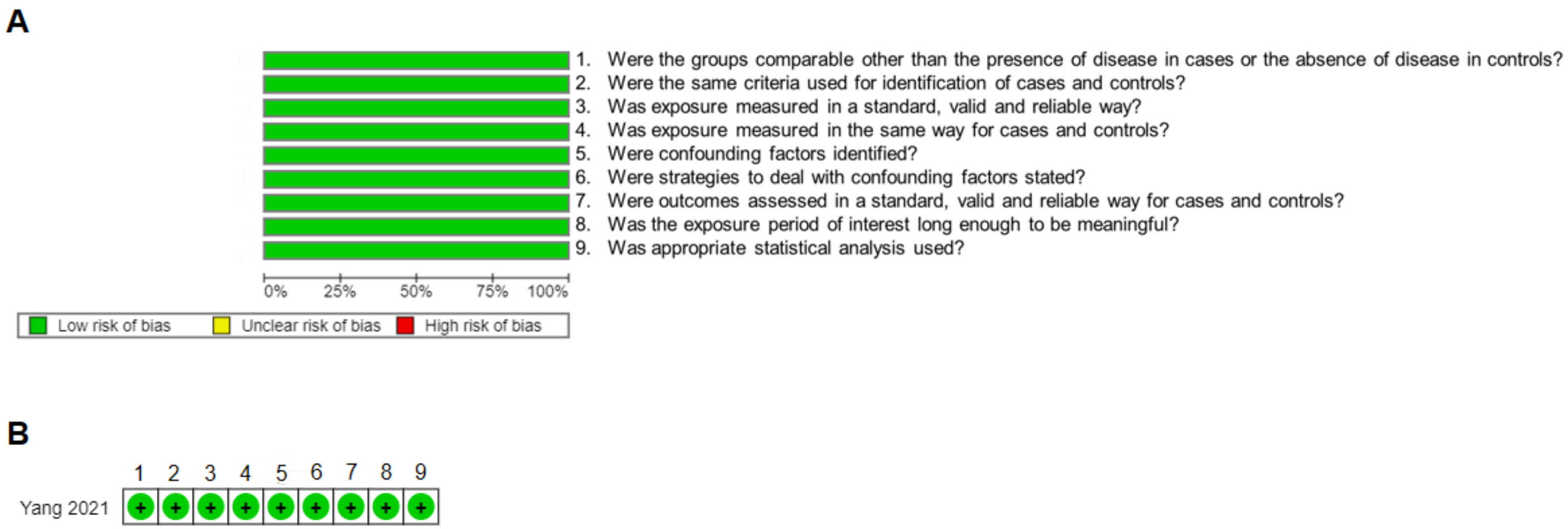

**Figure S5.** Risk of bias assessment of preconception risk factors in prospective studies.

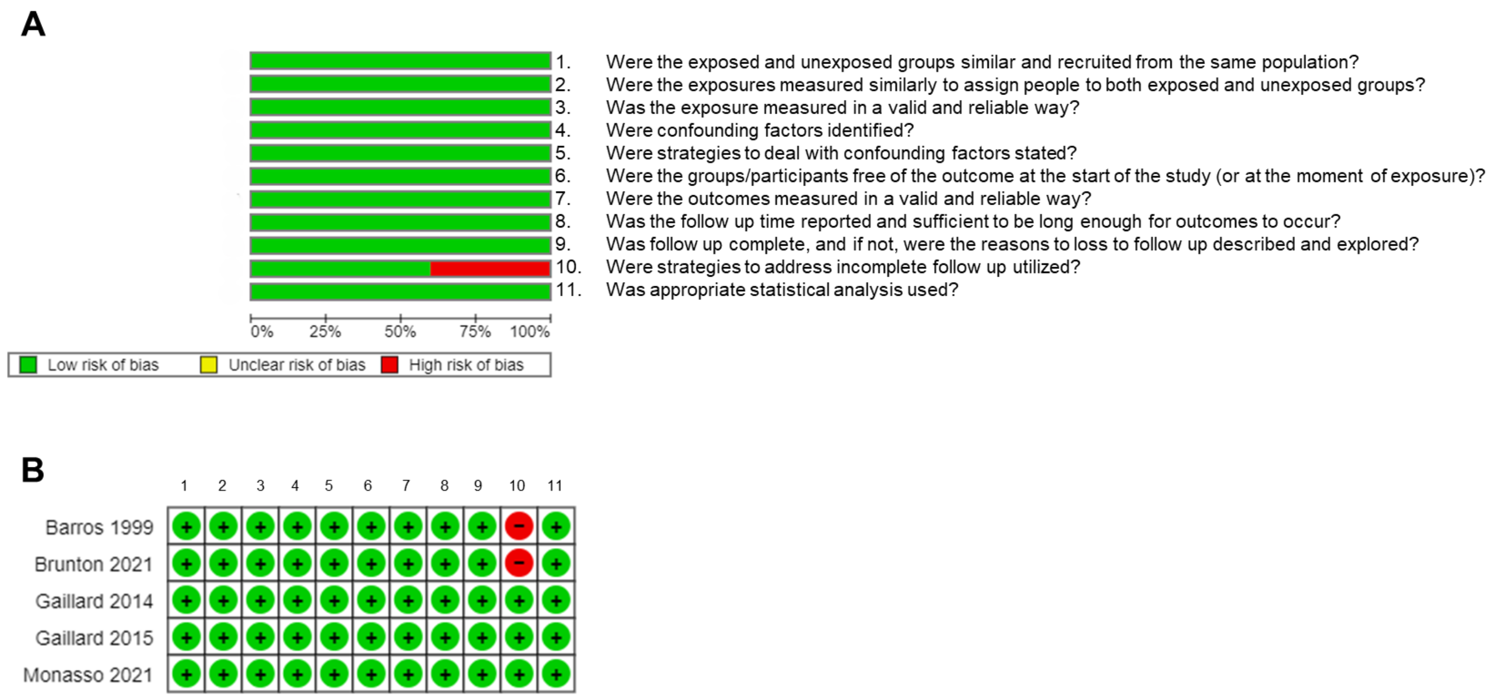

**Figure S6.** Risk of bias assessment of preconception risk factors in retrospective studies.

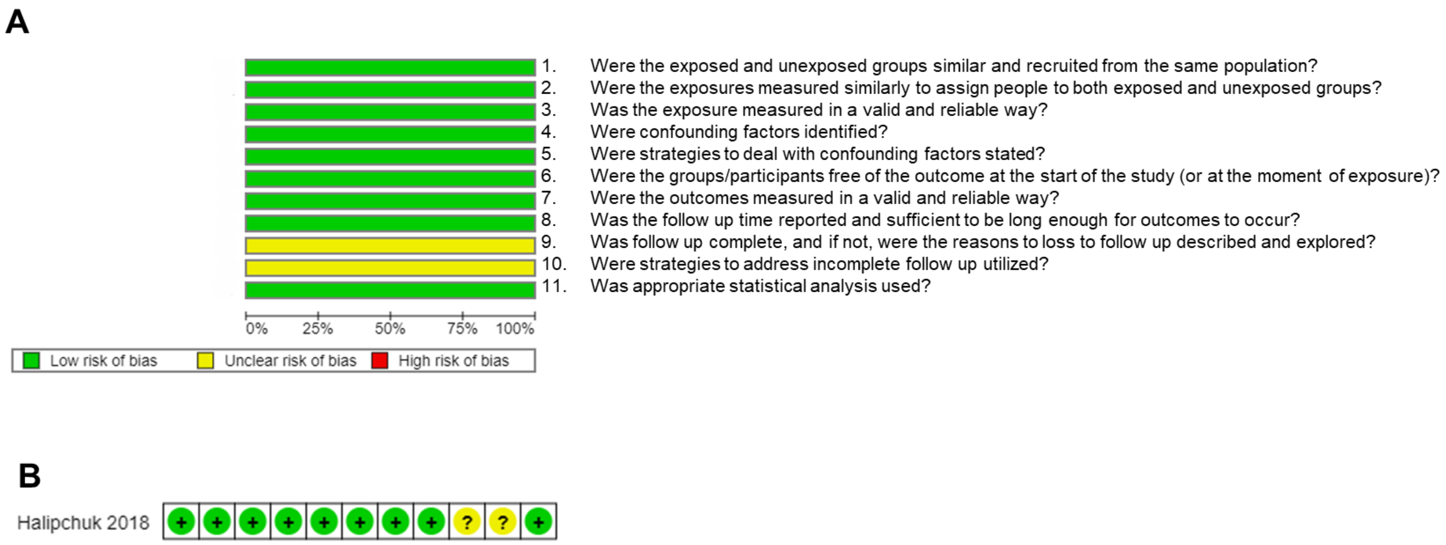

**Figure S7.** Risk of bias assessment of pregnancy and birth risk factors in case-control studies.

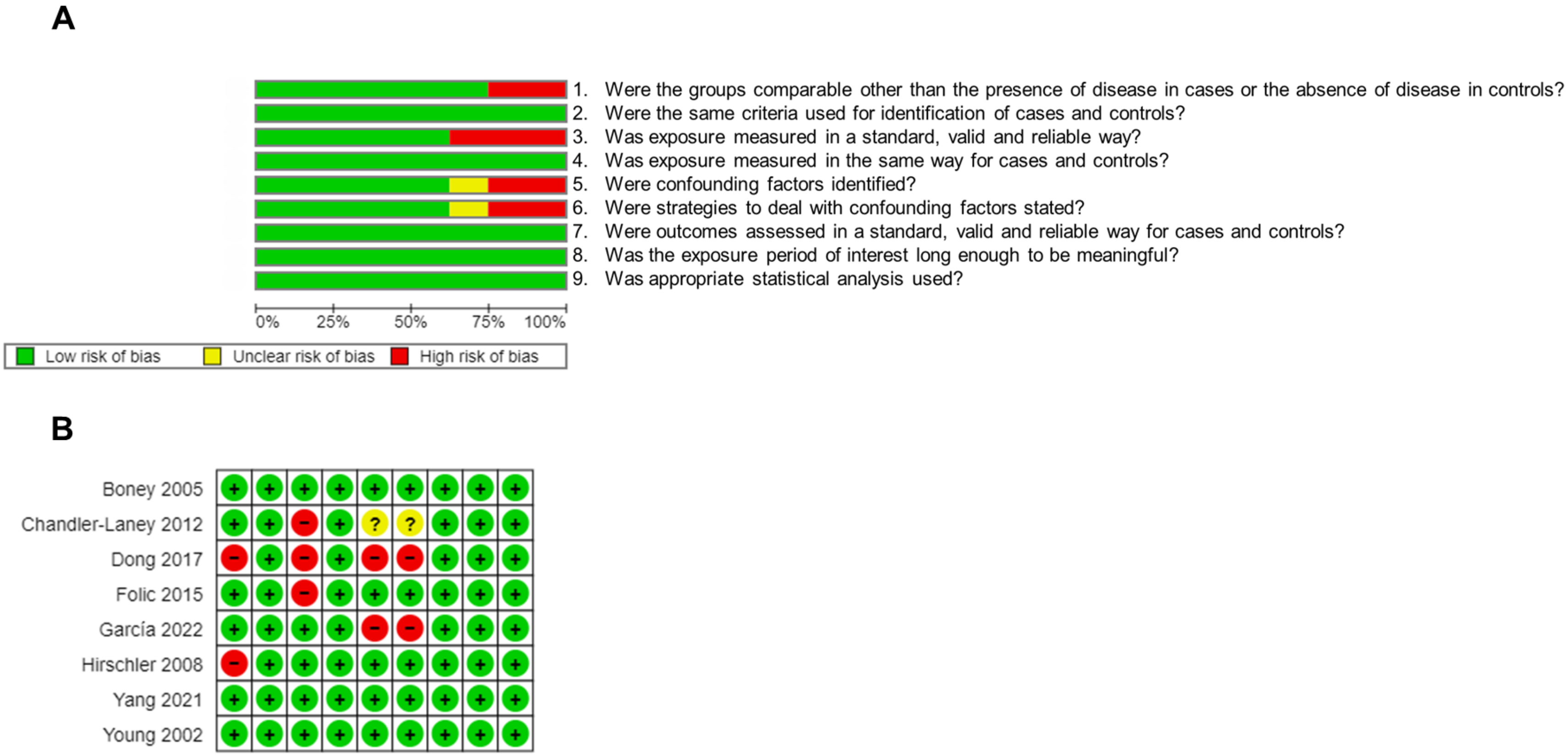

**Figure S8.** Risk of bias assessment of pregnancy and birth risk factors in randomized-controlled trials.

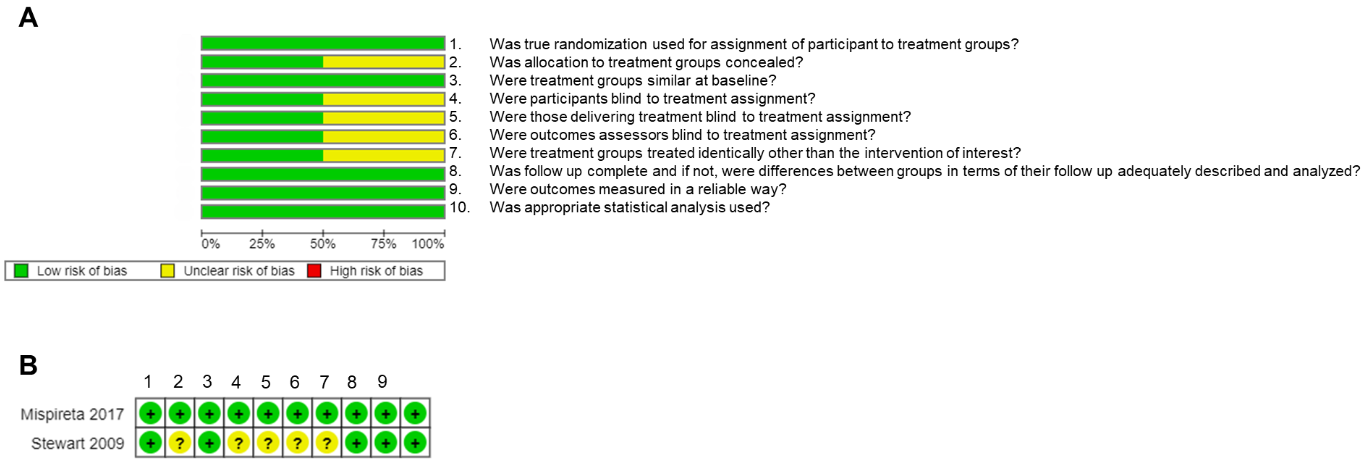

**Figure S9.** Risk of bias assessment of pregnancy and birth risk factors in prospective studies.

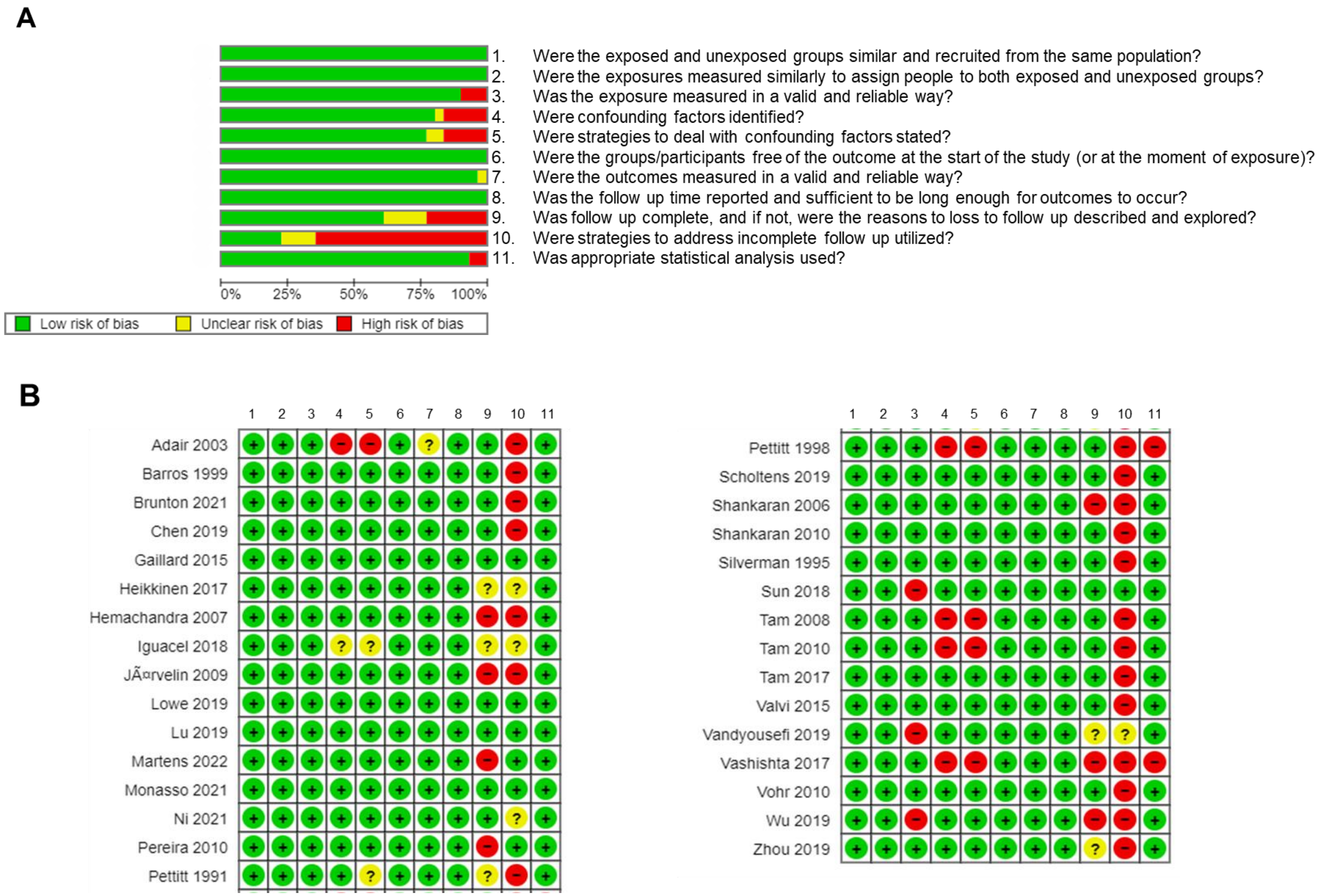

**Figure S10.** Risk of bias assessment of pregnancy and birth risk factors in retrospective studies.

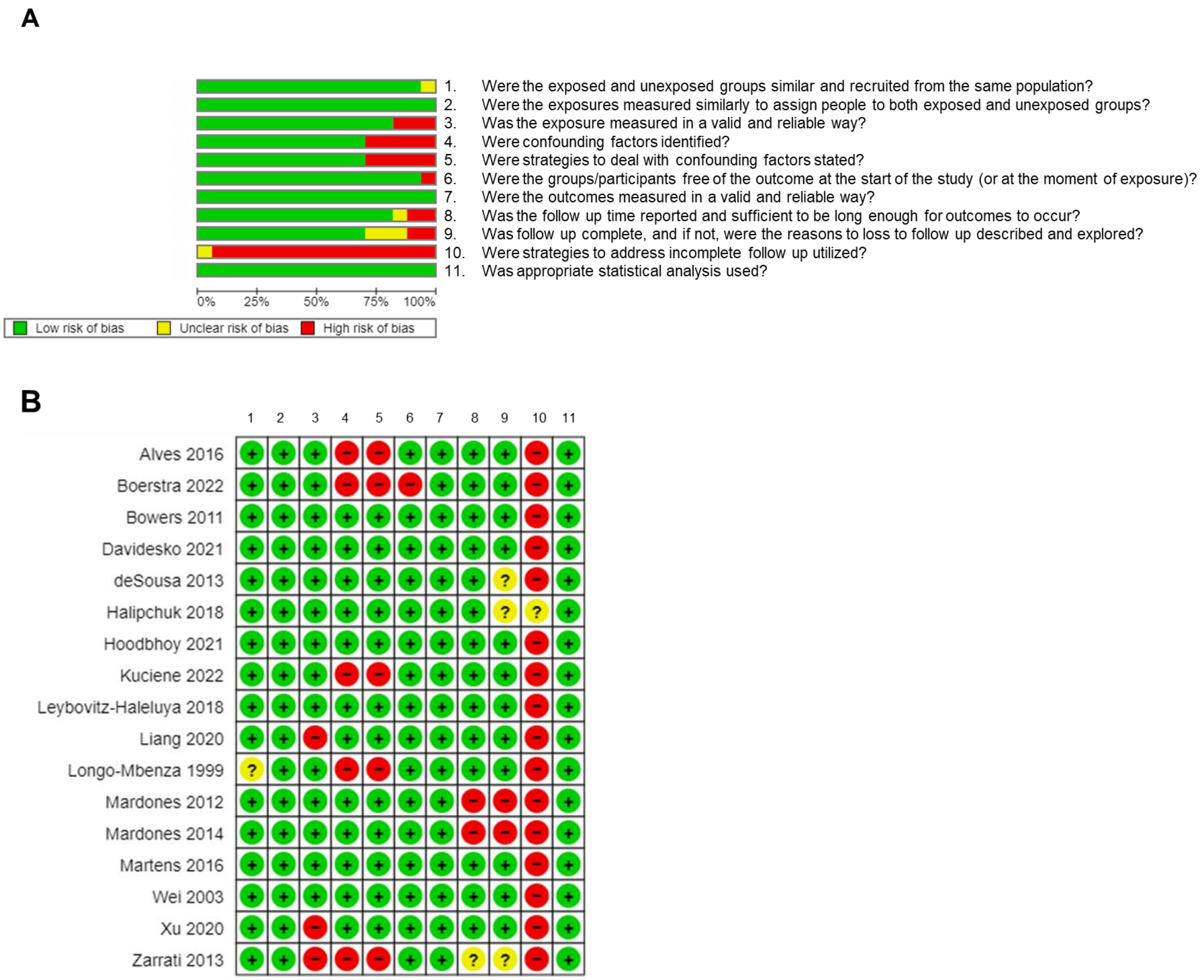

**Figure S11.** Risk of bias assessment of early infancy risk factors in case-control studies.

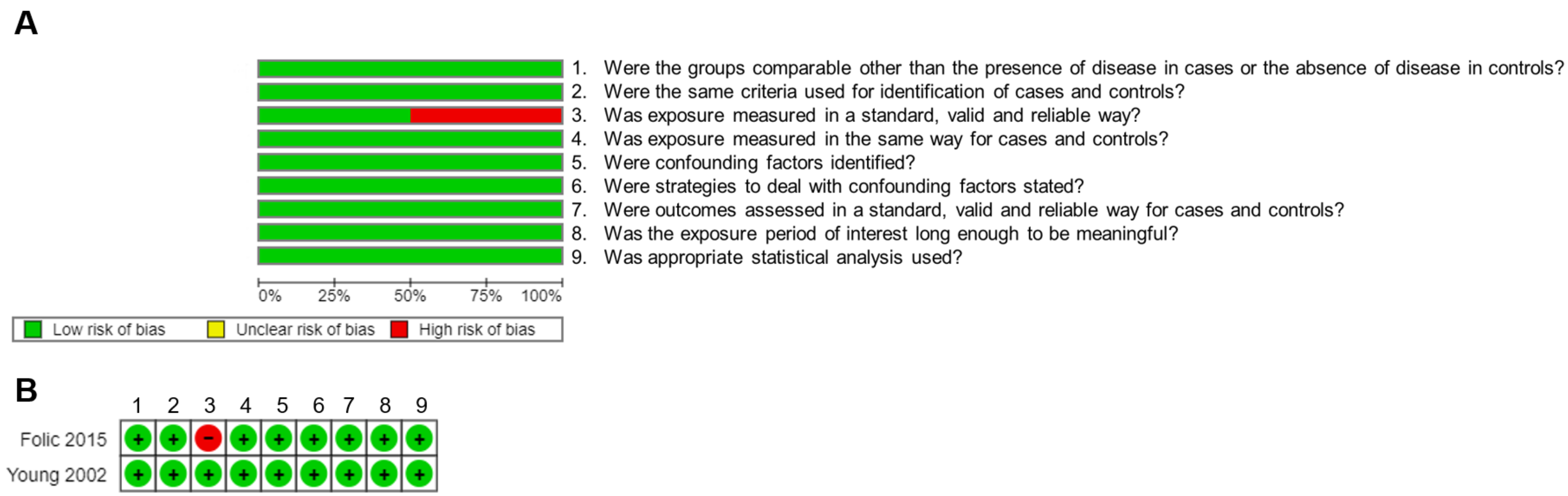

**Figure S12.** Risk of bias assessment of early infancy risk factors in prospective studies.

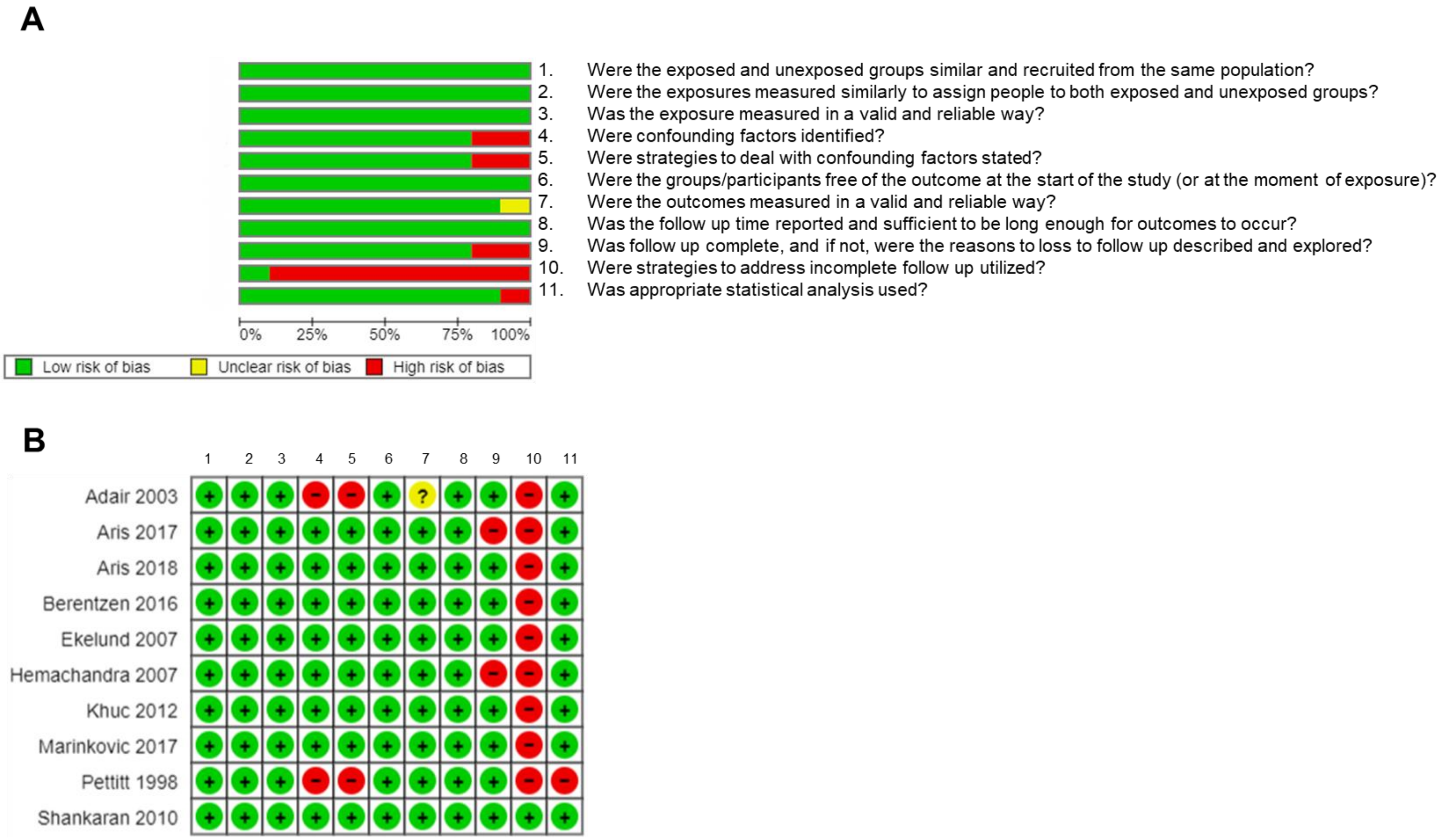

**Figure S13.** Risk of bias assessment of early infancy risk factors in retrospective studies.

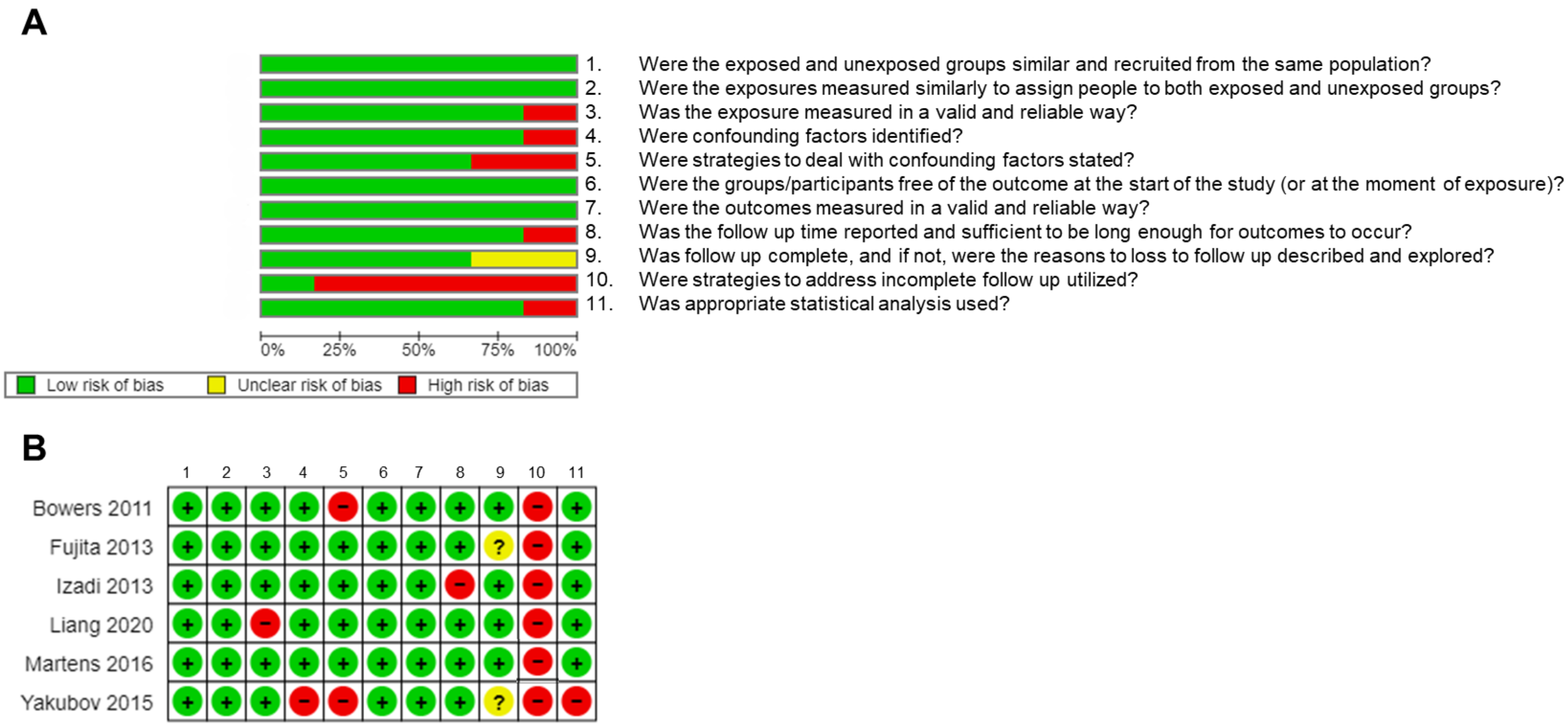

**Figure S14.** Assessment of bias in domains. Percentage of low risk, unclear and high risk of bias in 7 domains: population, measurement-outcome, measurement-exposure, confounding, duration of follow-up, lost to follow-up and statistical analysis for case-control studies (n=8) (**A**) and cohort studies (n=59) (**B**).

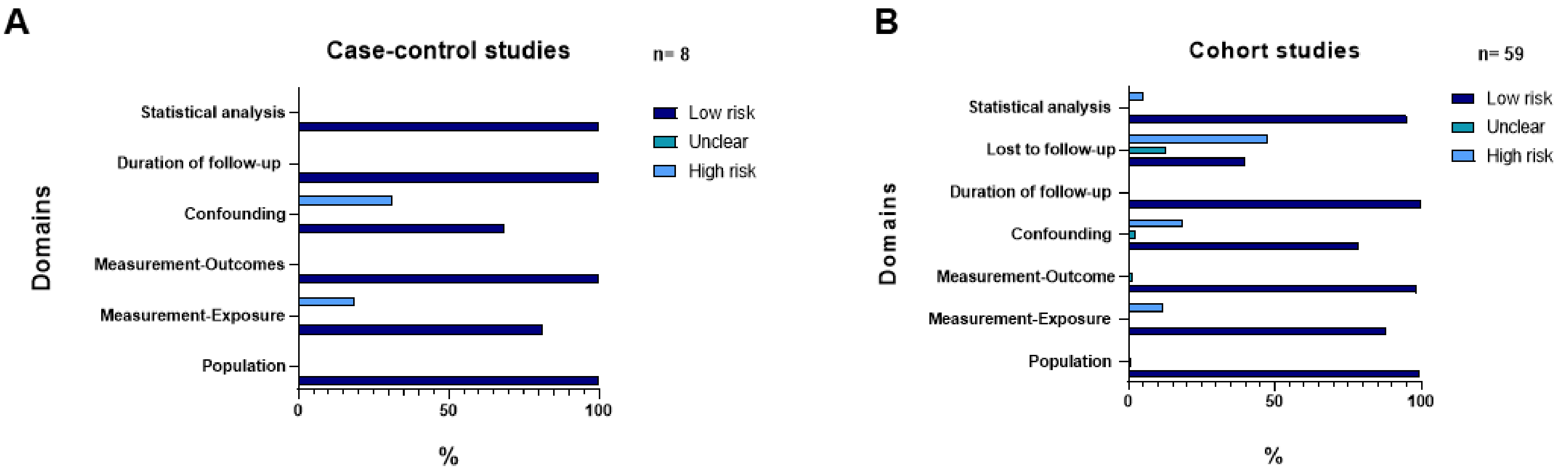
